# *In vivo* assessment of neurodegeneration in Spinocerebellar Ataxia type 7

**DOI:** 10.1101/2020.10.05.20207340

**Authors:** Jacob A. Parker, Shabbir H. Merchant, Sanaz Attaripour-Isfahani, Hyun Joo Cho, Patrick McGurrin, Brian P. Brooks, Albert R. La Spada, Mark Hallett, Laryssa A. Huryn, Silvina G. Horovitz

**Author notes:** Corresponding Author Silvina G. Horovitz, Human Motor Control Section, Medical Neurology Branch, National Institute of Neurological Disorders and Stroke, National Institutes of Health, 10 Center Drive, Bldg. 10, Rm. 7D37, Bethesda, MD 20892-1428.

## Abstract

Spinocerebellar Ataxia type 7 (SCA7) is a neurodegenerative disease characterized by progressive cerebellar ataxia and retinal degeneration. Increasing loss of visual function complicates the use of clinical scales to track the progression of motor symptoms, hampering our ability to develop accurate biomarkers of disease progression, and thus test the efficacy of potential treatments. In this cross-sectional study, we aimed to identify imaging measures of neurodegeneration, which may more accurately reflect SCA7 severity and progression. We analyzed diffusion tensor imaging (DTI) data collected from a cohort of 13 SCA7 patients and 14 healthy volunteers using two recent methodological advances: 1) a diffusion tensor-based image registration technique, and 2) a dual-compartment DTI model to control for the potential increase in extracellular CSF-like water due to neurodegeneration. These methodologies allowed us to assess both volumetric and microstructural abnormalities in both white and gray matter brain-wide in SCA7 patients for the first time. To measure tissue volume, we performed diffusion tensor-based morphometry (DTBM) using the tensor-based registration. To assess tissue microstructure, we computed the parenchymal mean diffusivity (pMD) and parenchymal fractional anisotropy (pFA) using the dual compartment model. This model also enabled us to estimate the parenchymal volume fraction (pVF), a measure of parenchymal tissue volume within a given voxel. While DTBM and pVF revealed tissue loss primarily in the brainstem, cerebellum, thalamus, and a subset of cerebral white matter tracts in patients, pMD and pFA detected microstructural abnormalities brain-wide (*p* < 0.05, FWE corrected; Hedge’s *g* > 1). This distinction was meaningful in terms of motor symptom severity, as we found that patient scores on the Scale for the Assessment and Rating of Ataxia correlated most strongly with cerebellar pVF (*r* = - 0.66, *p* = 0.015) and global white matter pFA (*r* = −0.64, *p* = 0.018). Since this contrast between focal tissue loss and global microstructural abnormality has previously been described in neuropathology, we believe the approach employed here is well suited for the *in-vivo* assessment of neurodegeneration. Moving forward, this approach could be applied to characterize the full spatiotemporal pattern of neurodegeneration in SCA7, and potentially develop an accurate imaging biomarker of disease progression.

**Highlights:**

- DTI study reveals brain-wide differences between SCA7 patients and controls.
- DTI dual-compartment model controls for increased CSF-like free water in patients.
- Tensor-based deformations show SCA7 tissue loss extends beyond cerebellum.
- Focal atrophy, but global microstructural abnormalities were observed in SCA7.
- Ataxia most correlated with cerebellar atrophy, global microstructural abnormality.

## 1. Introduction

Spinocerebellar Ataxia type 7 (SCA7) is an autosomal dominant neurodegenerative disease caused by a polyglutamine expansion in the *ATXN7* gene ^1-4^. The disease is characterized by progressive cerebellar ataxia and retinal degeneration, and patients progress toward significant loss of motor function and sometimes severe visual impairment ^1-4^. Potential routes of treatment have been identified ^5, 6^, but accurate biomarkers of progression are needed to assess efficacy. Clinical scales such as the Scale for the Assessment and Rating of Ataxia (SARA) ^7^ cannot capture the preclinical stages of the disease, and eventually become difficult to interpret as worsening visual impairment can confound the assessment of ataxia. Thus, a more rigorous measure of SCA7 deficits is warranted.

One potential solution to this problem may be to measure neurodegeneration *in vivo* in patients. Post-mortem neuropathology has revealed striking structural, cellular, and molecular abnormalities in patients ^1, 3, 8-11^. However, the evolution of this pathology and how it relates to disease progression remains unclear. Furthermore, the most appropriate way to probe these abnormalities *in vivo* is unclear.

Researchers have begun to address these challenges by using advanced magnetic resonance imaging (MRI) analyses such as voxel-based morphometry (VBM) and diffusion tensor imaging (DTI) to assess structural abnormalities *in vivo*. VBM, which measures differences in relative brain volume on a voxel-wise basis ^12^, has detected gray matter (GM) atrophy not only in the cerebellum, but also in many cortical areas including the sensorimotor cortices, motor association areas, cuneus, precuneus, insula, inferior frontal gyrus, medial frontal gyrus, inferior parietal lobule, and temporal regions. ^13-16^. DTI, which is typically used to assess WM microstructural integrity via measures of water diffusivity ^17, 18^, has detected microstructural abnormalities in the cerebellar WM, cerebellar peduncles, brainstem, cerebral peduncles, and several cerebral WM tracts including the internal and external capsules, corona radiata, optical radiation, corpus callosum, and miscellaneous temporal, occipital, parietal, and frontal WM in patients ^13, 19^. These abnormalities were indicated by both an increased mean diffusivity (MD), a measure of the magnitude of diffusion in all directions, and decreased fractional anisotropy (FA), a measure of how preferentially water diffuses in the principal direction of diffusion as compared to the other directions ^20^. These same changes are observed in many neurodegenerative WM pathologies ^21^, making these diffusion metrics a reliable marker of microstructural injury.

Despite the useful insights these methodologies have provided, they are limited in their ability to assess structural abnormalities across all anatomical structures and tissue types. VBM can only accurately measure volumetric differences in GM, while conventional DTI analyses are constrained to only the center of large, easily distinguishable WM tracts. We know from neuropathology that drastic volume loss in SCA7 has been noted in both the WM and GM of the cerebellum and brainstem, and that cellular and molecular abnormalities are found in virtually all tissues of the brain ^1, 3, 8-10, 22^. Another drawback of conventional DTI is that diffusion metrics derived from the standard single-compartment DTI model can be biased by an increase in cerebrospinal fluid (CSF)-like free water in and around the tissues, as can happen during pathologies such as neurodegeneration and edema ^23, 24^. When this occurs, DTI does not accurately reflect tissue microstructure. Therefore, the severity of atrophy associated with SCA7 presents a significant confound to conventional DTI analysis. Ideally, *in vivo* imaging metrics would be able to measure both gross volumetric and subtle microstructural changes across all tissue types and anatomical structures while remaining robust to substantial anatomical changes.

We address these challenges using two recently developed advancements in DTI analysis. First, we employed an image registration technique that uses information from the full diffusion tensor to accurately align anatomical structures across all tissue types ^25^. This allowed us to assess microstructural abnormalities in both WM and GM. It also allowed us to quantify the brain-wide gross volumetric abnormalities through diffusion tensor-based morphometry (DTBM), which uses the warps from subject to template space to determine which areas were larger or smaller relative to the template ^26^.

Second, we used a multi-shell diffusion weighted imaging (DWI) acquisition to estimate a dual-compartment, biexponential DTI model ^27^ to control for increases in CSF-like free water. This model assumes that the water diffusing in a given voxel can belong to one of two components: 1) a highly diffusing compartment of water in the interstitium (CSF-like free water), and 2) a relatively constrained compartment of water diffusing within the parenchyma. Diffusion metrics (FA and MD) can then be estimated from the parenchymal compartment alone, excluding the influence of diffusion in the CSF-like free water compartment. These metrics (parenchymal FA and parenchymal MD) have been shown to be more robust to the biases caused by neuropathology ^23, 27^ and have a higher test-retest reliability compared to metrics derived from the standard single-compartment DTI model ^28^.

Here we aimed to use these methodologies to more fully and accurately characterize the volumetric and microstructural differences between a cohort of SCA7 patients and a group of healthy volunteers. Furthermore, we sought to connect these structural abnormalities to ataxia severity.

## 2. Materials and Methods

### 2.1. Participants

Fourteen SCA7 patients were recruited from the Ophthalmic Genetics clinic at the National Institutes of Health between September 2014 and June 2018 from either the Genetics of Inherited Eye Disease Protocol (NCT02471287) or the Natural History of Spinocerebellar Ataxia type 7 study (NCT02741440). Fourteen healthy volunteers (HV) were recruited from the Physiological Investigations of Movement Disorders protocol (NCT01019343) between September 2014 and July 2016. SCA7 patients were required to have a CAG repeat expansion over 35 in the *ATXN7* gene, and were excluded if another condition not related to SCA7 was present that could complicate interpretation of the study data. HVs were excluded if any of the following were present: abnormal finding on neurological exam, illicit drug use in preceding 6 months, consumption of more than 7 or 14 alcoholic drinks per week for females and males respectively, psychiatric disorder, brain tumor, head trauma, history of head injury with loss of consciousness. All study participants were evaluated in the Human Motor Control Clinic by a movement disorder specialist. Ataxia severity was quantified in patients using the SARA. Patients with no measurable ataxia (a SARA score of 0) were seen under the protocol but excluded from this analysis since we wished to assess structural abnormalities in the context of tangible, quantifiable deficit. A history and physical, and a neurological exam were performed on HVs to screen for the presence of exclusionary conditions. All protocols were approved by the appropriate Institutional Review Board and informed consent was obtained from all participants prior to participation.

### 2.2. Image Acquisition

The following imaging sequences were collected on a 3-T MR750 GE scanner using a 32-channel head coil: 3-plane localizer, ASSET calibration, T1 weighted MPRAGE (3D inversion recovery, TR: 7664 ms, TE: 3.42 ms, TI: 425 ms, slice thickness: 1mm, 1×1 mm in-plane resolution, percent phase FoV: 100 mm^2^, flip angle: 7, matrix size: 256×256), DWI ^29^ (Fat suppression, 62 slices, TR 6751.68 ms, TE 79 ms, voxel size 2.5mm isotropic, acceleration factor (ASSET) 2, b-values (volumes): b=0 (10), intermediate b = 300 (10) and b = 1100 (60) s/mm^2^ and phase-encode direction anterior-posterior), and T2 weighted (T2W) fast spin-echo volume (62 slices, TR: 7500 ms, TE: 100.74 ms, slice thickness: 2.5 mm, 0.9375 mm × 0.9375 mm in-plane resolution, FoV: 240×180 mm^2^, percent phase FoV: 80, flip angle: 90, matrix size 256×192).

### 2.3. Image Preprocessing

Each participant’s T2W image was reconstructed and axialized by rigid alignment to an AC-PC aligned MNI template using FATCAT ^30^. Each raw DWI volume was visually inspected and those containing significant motion artifact were removed. Using TORTOISE (V3.1.2) ^31, 32^, DWIs were then corrected for Gibbs ringing, subject motion, and eddy-current distortion by appropriate rotation of the b-matrix for each volume.

### 2.4. Dual Compartment Model

The diffusion tensor was also fit using a dual-compartment model introduced by Pierpaoli and Jones 2004 ^27^ and implemented in TORTOISE. This model assumes that the water diffusing in a given voxel can belong to one of two components: 1) a highly diffusing compartment of water in the interstitium (CSF-like free water), and 2) a relatively constrained compartment of water diffusing within the parenchyma. In this model, the observed signal intensities *S* for a set of diffusion weightings *(b*-matrix) is a biexponential decay function given by the following equation:

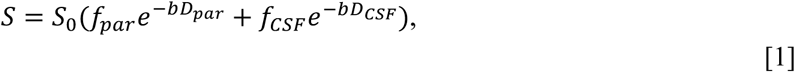

where *S*_0_ is the signal intensity when there is no diffusion weighting (b=0); *b* is the *b*-matrix; *D*_par_, *D*_CSF_ are the diffusion tensors of the CSF-like free water and parenchymal compartments respectively; and *f*_par_, *f*_CSF_ are the volume fractions of the CSF-like free water and parenchymal compartments respectively. *f*_par_ and *f*_CSF_ represent volume fractions because each term indicates the proportion of water in a given voxel belonging to that compartment. Assuming a relatively uniform distribution of water, and assuming negligible transfer between each compartment, the proportion of water in a given compartment is equal to the proportion of volume that compartment takes up within a voxel.

The model assumes that the CSF-like free water and parenchymal compartments together comprise the entire volume of a given voxel, thus the sum of *f*_par_ and *f*_CSF_ is set to 1. Furthermore, previous results ^27, 33^ indicate that *D*_CSF_ is well approximated by an isotropic tensor with a fixed diffusivity of 3×10^−3^ mm^2^/s, which is the diffusivity of free water at 37°C. Thus, this biexponential, dual-compartment model simplifies to the following:

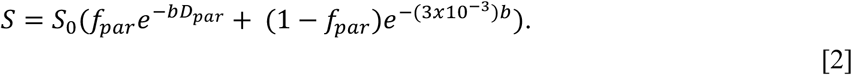

Finally, the diffusivity of the parenchymal diffusion tensor, *D*_par_, is constrained to < 3×10^−3^ mm^2^/s since diffusion in the parenchymal compartment should be more restricted than in the CSF-like free water compartment. No constraint is placed on the isotropy of *D*_par_, thus it takes the form of the standard diffusion tensor.

Model parameters *f*_par_ and *D*_par_ as given in Eq 2 were fit to the processed DWI data for each subject using nonlinear least squares. The use of a multi-shell acquisition with two distinct nonzero b-values (10 volumes at b=300 s/mm^2^ and 60 volumes at b=1100 s/mm^2^) enabled accurate estimation of exponential signal decay attributable to the isotropic CSF-like free water component, and thus accurate estimation of *f*_par_ and *D*_par_ ^34^.

The FA and MD of the parenchymal compartment were computed with non-linear fitting from *D*_par_ to assess tissue microstructure without the influence of the CSF-like free water compartment. Henceforth, we will refer to these measures as the parenchymal FA (pFA) and the parenchymal MD (pMD).

While we place emphasis on the pFA and pMD, the parenchymal volume fraction (pVF), denoted so far as *f*_par_, is itself a useful metric of tissue composition and integrity. As described above, the pVF indicates the proportion of volume in a given voxel comprised of parenchymal tissue. The remaining volume fraction is associated with the CSF-like free water compartment, which could belong to a number of extracellular spaces including CSF, circulation, perivascular, and lacunae in the tissue more than a few tens of microns in diameter. Thus, a decrease in the pVF necessarily indicates an increase in this extracellular volume fraction, which can indicate the presence of pathological processes.

### 2.5. Single Compartment Model

The diffusion tensor was also non-linearly fit using the conventional monoexponential, single-compartment model ^17^. As described below, the single-compartment diffusion tensor images were used to create a study-specific template and to perform spatial normalization. Furthermore, the diffusivity metrics of FA and MD were computed for comparison with the pFA and the pMD.

### 2.6. Diffusion Tensor Template and Spatial Normalization

To perform brain-wide group comparisons, we created a study-specific template and normalized each subject’s images to that template using DR-TAMAS ^25^. DR-TAMAS uses the full diffusion tensor to compute the transformation from subject space to standard space, achieving an accurate alignment across all tissue types and anatomical structures.

We created a template reflective of the entire study population using the single-compartment diffusion tensor images of an equal number (n=13) of HVs and patients. Compared to an HV-only template, this population template lessened the chance that one group’s spatial transformations were significantly larger than the other, which could introduce bias to the group comparison. Each subject’s single compartment diffusion tensor image was registered to this template to create a transformation. The scalar maps of the metrics described above (pFA, pMD, pVF, FA, MD) were computed in subject space, then subsequently brought to standard space by application of this transformation.

All statistical testing was performed in the standard space defined by the population template. Given the high level of atrophy present in the patient population, an HV only template was created for the purpose of visualizing the results of the statistical analysis. The population template was registered to the HV template and the resulting transformation was applied to the statistical maps produced during the statistical analysis.

### 2.7. Diffusion Tensor-Based Morphometry

The transformation from each subject’s space to the standard space was also used to perform DTBM, which allows for quantification of morphological differences between two groups on a voxel-wise basis ^26^. Since these transformations were computed using information from the full diffusion tensor, they contain accurate information about how all tissue types must be deformed to match the population template. We used each subject’s transformation to compute a natural log of the Jacobian determinant (logJ) map, which quantifies a subject’s brain volume relative to the template at each voxel.

### 2.8. Voxel-Based Morphometry

To assess how our morphometric analysis compared to more conventional methods, we also performed VBM using FSL-VBM ^12, 35, 36^. First, brain extraction followed by GM segmentation was performed on each subject’s T1W image. Each subject’s GM image was then nonlinearly registered to the Montreal Neurological Institute (MNI)152 standard space template. Next, a study-specific population GM template was created by averaging an equal number of SCA7 and HV GM images (13 each) and flipping along the x-axis to make it left-right symmetric. In a similar manner to the study-specific population diffusion tensor template described above, the use of a study-specific population template here minimizes the possibility that one group could require significantly larger deformations to be registered to the template and thus bias the group comparison ^37^. All subject GM images in subject space were nonlinearly registered to this template and corrected for local expansion and contraction (modulations). Finally, the corrected GM images were smoothed using a Gaussian kernel with a sigma of 2. Henceforth, we will refer to these corrected and smoothed GM images as GM maps.

### 2.9. Total Intracranial Volume Estimation and Segmentation

We processed each subject’s T1 weighted MPRAGE using the default cortical reconstruction process of FreeSurfer ^38^ in order to segment the brain and estimate the Total Intracranial Volume (TICV) ^39^. The TICV was used as a covariate for both the logJ group comparison and the GM map group comparison since the TICV is correlated with the volume of individual brain structures ^40^.

### 2.10. Statistical Testing

We performed voxel-wise comparisons evaluating the effect of group (SCA7 vs HV) on the pFA, pMD, pVF, FA, MD, logJ, and GM maps. We used the FSL randomise function ^35, 41^ to perform permutation-based nonparametric tests using the threshold-free cluster enhancement method with a family-wise error rate of 5%. The comparison of the logJ and GM maps both used a design with group as the covariate of interest and age and TICV as additional covariates. The comparison of the other metrics (pFA, pMD, pVF, FA, MD) all used a design with group as the covariate of interest and age as an additional covariate.

Considering the number of comparisons and that many methods exist for performing voxel-wise comparisons, we additionally computed Hedge’s *g* effect size maps for each group comparison. These maps were used to assess the statistical power of the comparisons performed by FSL’s randomise function (see Supplement 1 section S2).

In patients, we additionally tested whether the SARA score was significantly correlated with each DTI metric (pMD, pFA, pVF, logJ) independently. Due to the relatively low number of patients in our cohort, we tested correlations between the average value of each metric in the brainstem, cerebellum, and cerebrum with the SARA score.

Each correlation was tested in R using the *lm* function and corrected for multiple comparisons using the Bonferroni procedure. We also assessed the same set of correlations including the asymptomatic patient to see if the observed correlations remained consistent.

### 2.11. Masking for Group Comparisons and Correlation Analyses

The group comparisons of the logJ, pMD, and pVF were conducted across the entire brain. Thus, we used a simple whole brain mask computed from the population diffusion tensor template using the 3dAutomask function of AFNI ^42^. Since FA is not an informative measure in the GM, we analyzed the pFA and FA only in the WM. We defined WM to be any region in the population diffusion tensor template with an FA > 0.2 and created a mask from this criterium for the pFA and FA group comparisons. Finally, the group comparison of the GM maps was performed only within the GM mask created by FSL-VBM.

To create masks corresponding to the brainstem, cerebellum, and cerebrum for the correlation analysis, we used the FreeSurfer segmentation of the subject that was used as the initial basis of the population diffusion tensor template. The segmentation was normalized to the population template using the transformation from that subject to the template computed by DT-TAMAS. Masks of each region were formed from the segmentation and used to extract the average value of each metric in each region for each subject. Similar to the group comparison, we only extracted the average pFA from the WM portion of each region, which we defined to be where the FA of the template exceeded 0.2.

### 2.12. WM and GM Atlas Tables

The results of all voxel-wise group comparisons were tabularized using standard atlases of WM and GM regions in the brain. For the WM, we utilized the ICBM-DTI-81 atlas of 48 WM regions ^43, 44^ distributed with FSL. We will refer to this atlas as the ICBM-DTI-81 WM atlas. For the GM, we used a FreeSurfer GM atlas created from the combination of subcortical GM ROIs derived from whole brain segmentation ^45^ and cortical GM ROIs derived from the Desikan-Killiany atlas ^46^. Altogether, this GM atlas contained 87 ROIs. We will refer to this atlas as the FreeSurfer GM atlas. A full listing of all regions in both the ICBM-DTI-81 WM atlas and the FreeSurfer GM atlas can be found in the tables of Supplement 2.

We determined the peak *t*-statistic, peak Hedge’s *g*, and the proportion of statistically significant voxels (Qvoxels) within each atlas region for every voxel-wise group comparison performed. Qvoxels was calculated by dividing the number of statistically significant voxels by the number of total voxels within a given ROI. Since the pFA and FA group comparisons were only performed in voxels where the FA of the population diffusion tensor template exceeded 0.2, we masked the atlases by the WM skeleton formed by this criterion when tabularizing these metrics. Furthermore, we did not include the cortical regions of the FreeSurfer GM atlas when tabularizing the pFA and FA group comparisons because the FA is not an informative measure in cortical GM. We similarly did not tabularize the results of the VBM group comparison using the ICBM-DTI-81 WM atlas.

Due to the number of regions in each atlas, we only list the regions for which Qvoxels was at least 0.1 in the tables presented in the main text. The corresponding tables for the VBM, MD, and FA group comparisons can be found in section S3 of Supplement 1. Tables including every region from each atlas for every group comparison can be found in Supplement 2 section S6. Tables including every metric next to each other for easy comparison can be found in Supplement 2 section S5.

## 3. Results

### 3.1. Participants and Preprocessing

One patient was excluded for having a SARA score of 0, thus thirteen patients were used for data analysis (mean age: 36.4 years; range: 16-62; 9 female, 4 male; CAG repeat range 40-66; mean SARA: 13.7; range: 2.5-22). All 14 HVs were included in the analyses (mean age: 44.5; range: 22-64; 9 female, 5 male). Age was not significantly different between groups (Wilcoxon Rank Sum Test; *p*=0.16, U=152.5). Examination of all included participants revealed the absence of all of the following: abnormal finding on neurological exam (not related to SCA7 in patients), illicit drug use in preceding 6 months, consumption of more than 7 or 14 alcoholic drinks per week for females and males respectively, psychiatric disorder, brain tumor, head trauma, history of head injury with loss of consciousness.

During visual quality control inspection of the raw DWIs, 10 out of 1040 volumes were removed across all SCA7 patients and 0 out of 1120 volumes were removed across all HVs. The largest number of volumes removed in a single patient was 3 out of the 80 DWI volumes, which meant each participant had sufficient data for diffusion tensor modeling.

### 3.2. DTBM reveals focal gross volumetric loss in SCA7

SCA7 patients had a significantly smaller logJ, and thus significantly less tissue volume, in the cerebellar peduncles, corticospinal tracts, bilateral medial lemniscus, cerebral peduncles, internal capsules, superior corona radiata, posterior corona radiata, bilateral fornix/stria terminalis, bilateral superior fronto-occipital fasciculus, cerebellar WM and GM, brainstem, thalamus, bilateral hippocampus, and ventral diencephalon (Figure 1, Tables 1 and 2, and Tables S5.1, S5.2, S6.1.1, S6.1.2 in Supplement 2). A significantly higher logJ was observed in the CSF spaces around the cerebellum, the 4^th^ ventricle, and in the longitudinal fissure of the occipital lobe near the lingual gyri. A large (>1 Hedge’s *g*) effect size was observed in all of the same regions listed above (Figure S2.1).

**Table 1:**
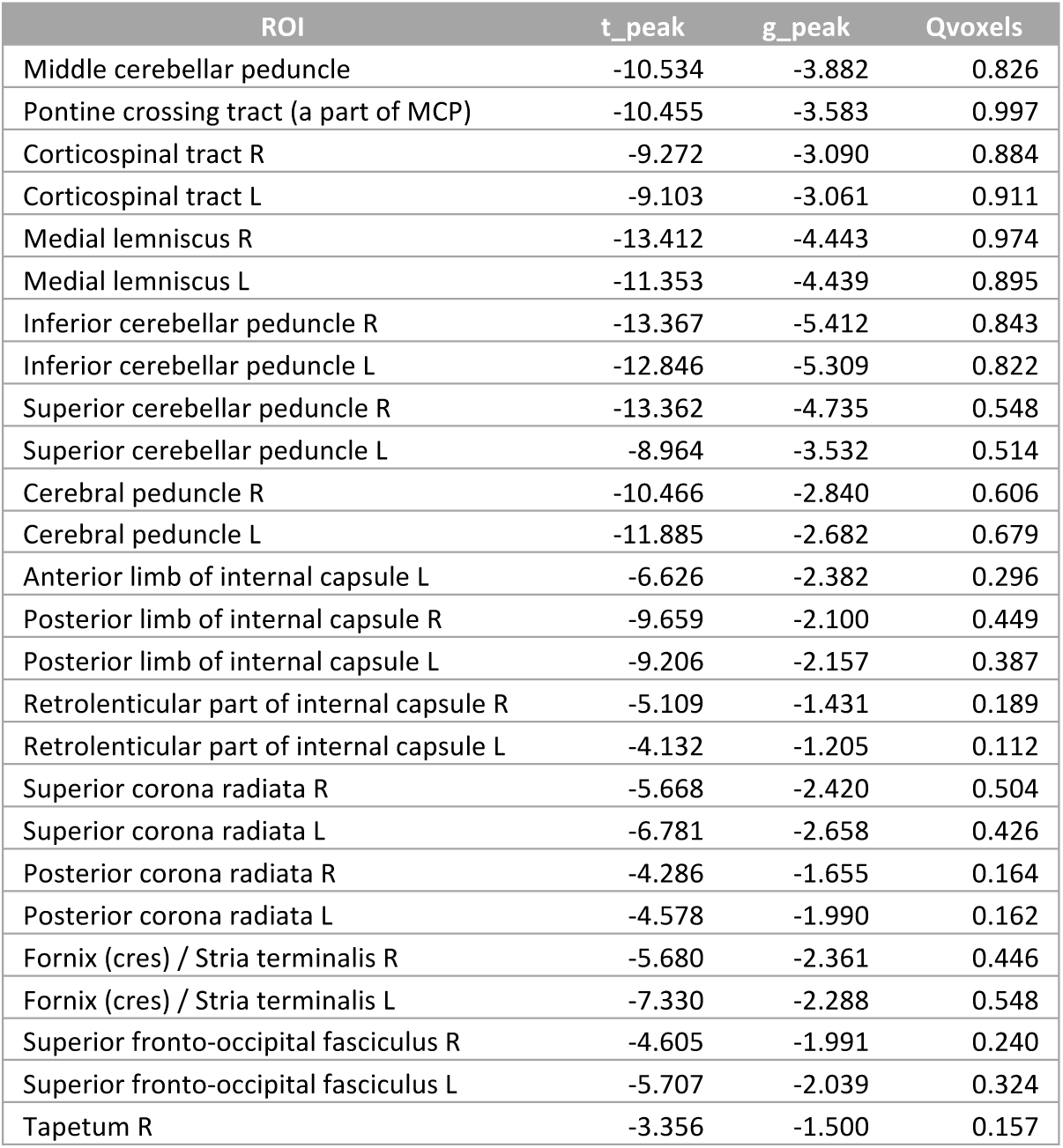
Peak *t*-statistic value, peak Hedge’s *g* value, and the fraction of ROI voxels significantly different between HVs and SCA7 patients (Qvoxels) for the DTBM group comparison within ROIs of the ICBM-DTI-81 WM atlas. Only regions with Qvoxels ≥ 0.10 are shown.

**Table 2:**
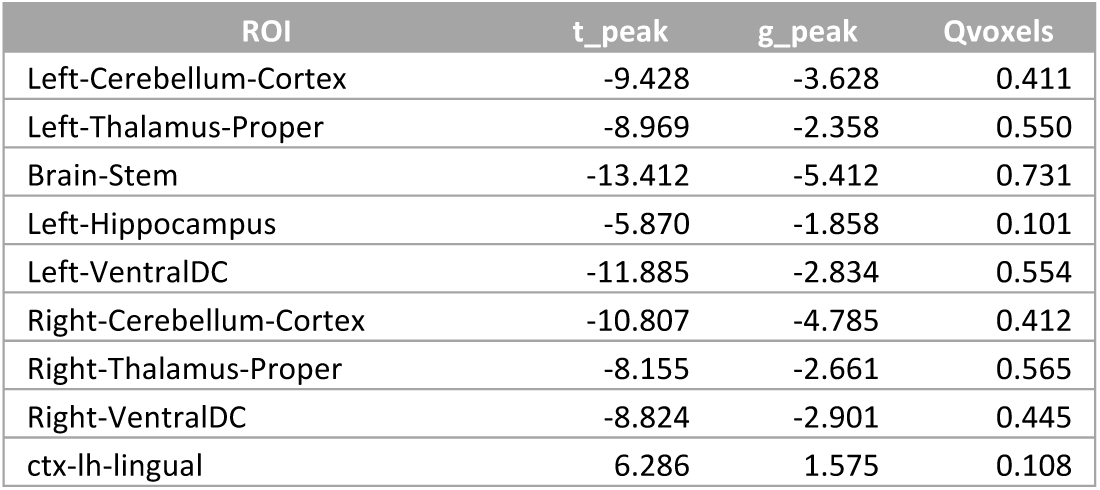
Peak *t*-statistic value, peak Hedge’s *g* value, and the fraction of ROI voxels significantly different between HVs and SCA7 patients (Qvoxels) for the DTBM group comparison within ROIs of the FreeSurfer GM atlas. Only regions with Qvoxels ≥ 0.10 are shown.

| ROI | t_peak | g_peak | Qvoxels |
| --- | --- | --- | --- |
| Left-Cerebellum-Cortex | -9.428 | -3.628 | 0.411 |
| Left-Thalamus-Proper | -8.969 | -2.358 | 0.550 |
| Brain-Stem | -13.412 | -5.412 | 0.731 |
| Left-Hippocampus | -5.870 | -1.858 | 0.101 |
| Left-VentralDC | -11.885 | -2.834 | 0.554 |
| Right-Cerebellum-Cortex | -10.807 | -4.785 | 0.412 |
| Right-Thalamus-Proper | -8.155 | -2.661 | 0.565 |
| Right-VentralDC | -8.824 | -2.901 | 0.445 |
| ctx-lh-lingual | 6.286 | 1.575 | 0.108 |

**Figure 1:**
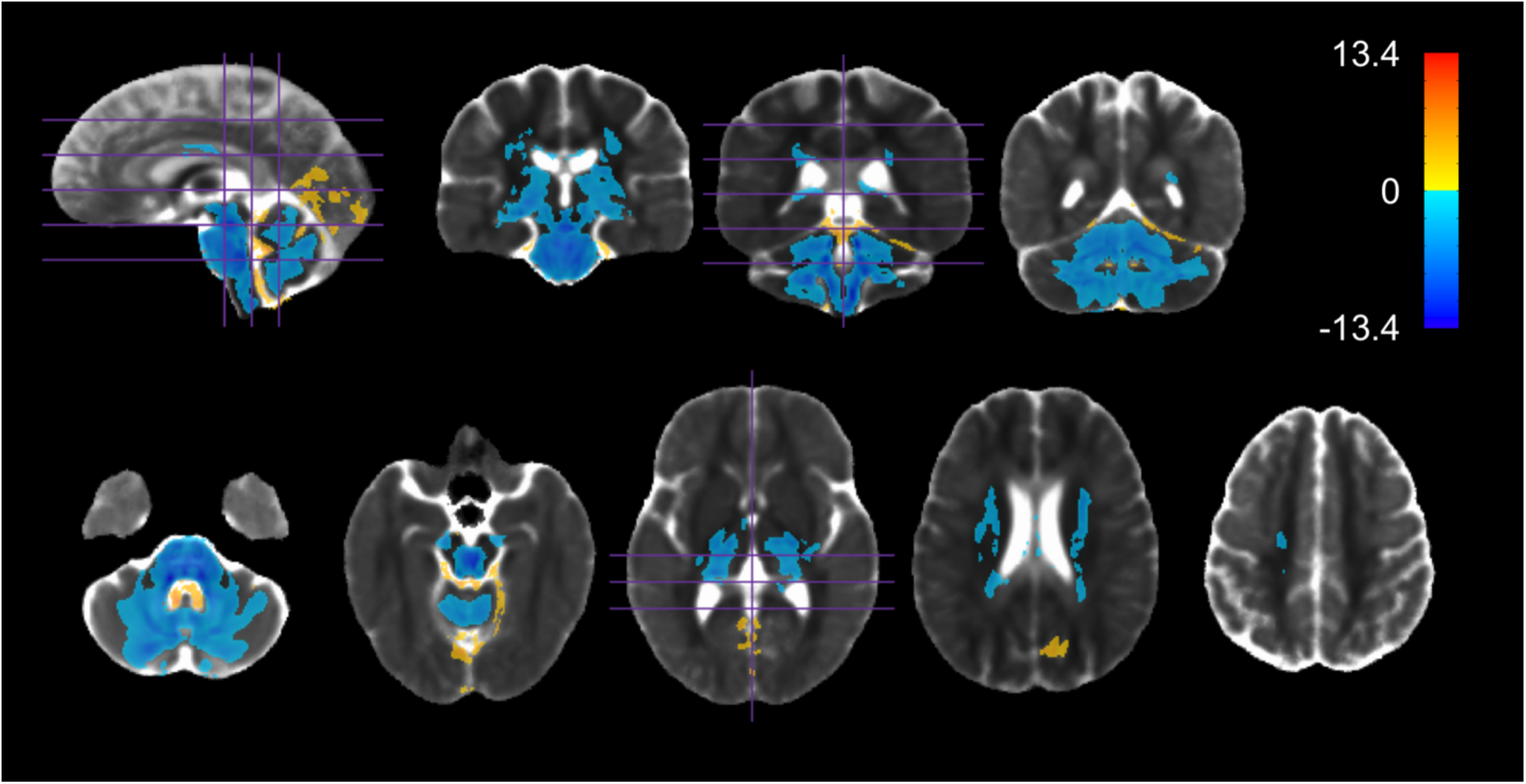
Regions in which logJ was significantly different between HVs and SCA7 *(p<*0.05, familywise error corrected). Values shown are the t-statistic. Blue indicates lower values (lower logJ/pVF) in SCA7 patients versus HVs. Orange indicates higher values in SCA7 patients versus HVs. Results are overlaid on the HV diffusion tensor template.

The group comparison of the GM maps from VBM revealed significant volume loss in all of the same GM regions in patients that DTBM detected volume loss in. Notably, this comparison also detected GM volume loss in the bilateral sensorimotor cortices, which DTBM did not detect (Figures S1.1 and S2.5, Table S3.1.1 in Supplement 1, and Tables S5.2, S6.5.1 in Supplement 2).

### 3.3. pVF reveals significant parenchyma loss in brainstem and cerebellum

SCA7 patients had a significantly lower pVF around the cerebellar peduncles, bilateral medial lemniscus, cerebral peduncles, bilateral fornix/stria terminalis, cerebellar cortices, brainstem, bilateral hippocampus, and ventral diencephalon (Figure 2, Tables 3 and 4, and Tables S5.1, S5.2, S6.2.1 and S6.2.2 in Supplement 2). Notably, no decrease in pVF was found in any tissue not directly adjacent to CSF (internal brainstem, internal cerebellar WM, almost all of the cerebrum). A large (>1 Hedge’s *g*) effect size was observed in the same regions (Figure S2.2).

**Table 3:**
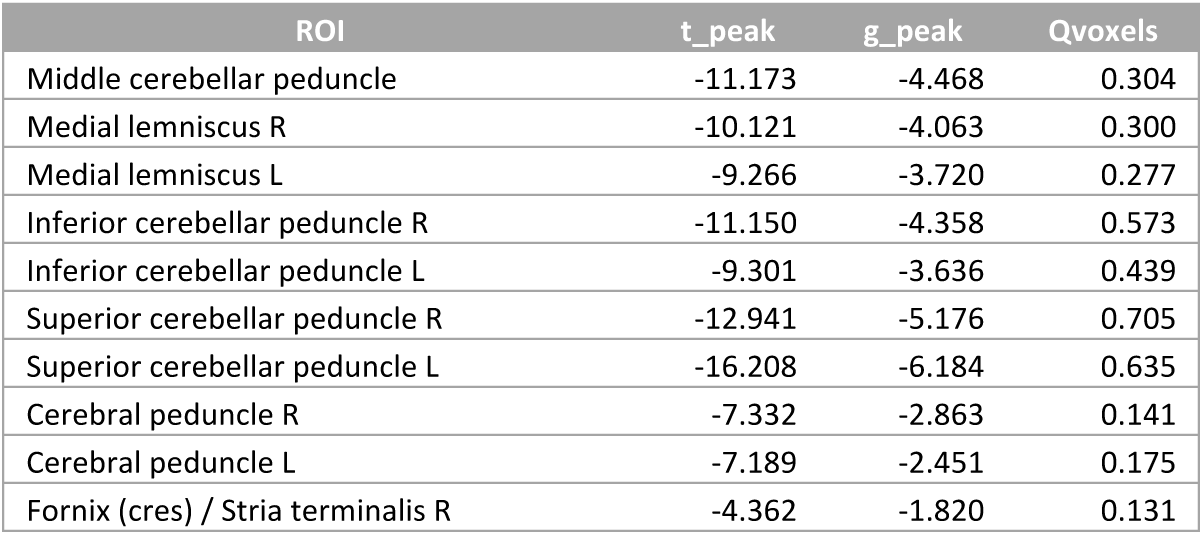
Peak *t*-statistic value, peak Hedge’s *g* value, and the fraction of ROI voxels significantly different between HVs and SCA7 patients (Qvoxels) for the pVF group comparison within ROIs of the ICBM-DTI-81 WM atlas. Only regions with Qvoxels ≥ 0.10 are shown.

**Table 4:**
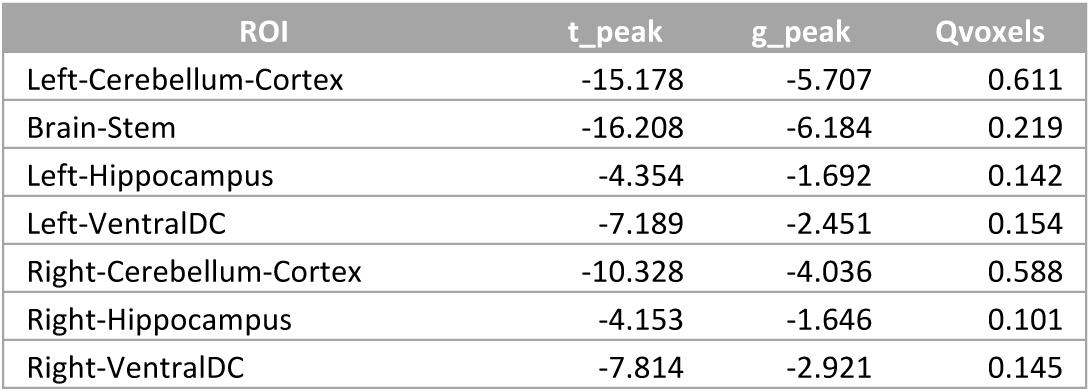
Peak *t*-statistic value, peak Hedge’s *g* value, and the fraction of ROI voxels significantly different between HVs and SCA7 patients (Qvoxels) for the pVF group comparison within ROIs of the FreeSurfer GM atlas. Only regions with Qvoxels ≥ 0.10 are shown.

| ROI | $t_{\text{peak}}$ | $g_{\text{peak}}$ | Qvoxels |
| --- | --- | --- | --- |
| Left-Cerebellum-Cortex | -15.178 | -5.707 | 0.611 |
| Brain-Stem | -16.208 | -6.184 | 0.219 |
| Left-Hippocampus | -4.354 | -1.692 | 0.142 |
| Left-VentralDC | -7.189 | -2.451 | 0.154 |
| Right-Cerebellum-Cortex | -10.328 | -4.036 | 0.588 |
| Right-Hippocampus | -4.153 | -1.646 | 0.101 |
| Right-VentralDC | -7.814 | -2.921 | 0.145 |

**Figure 2:**
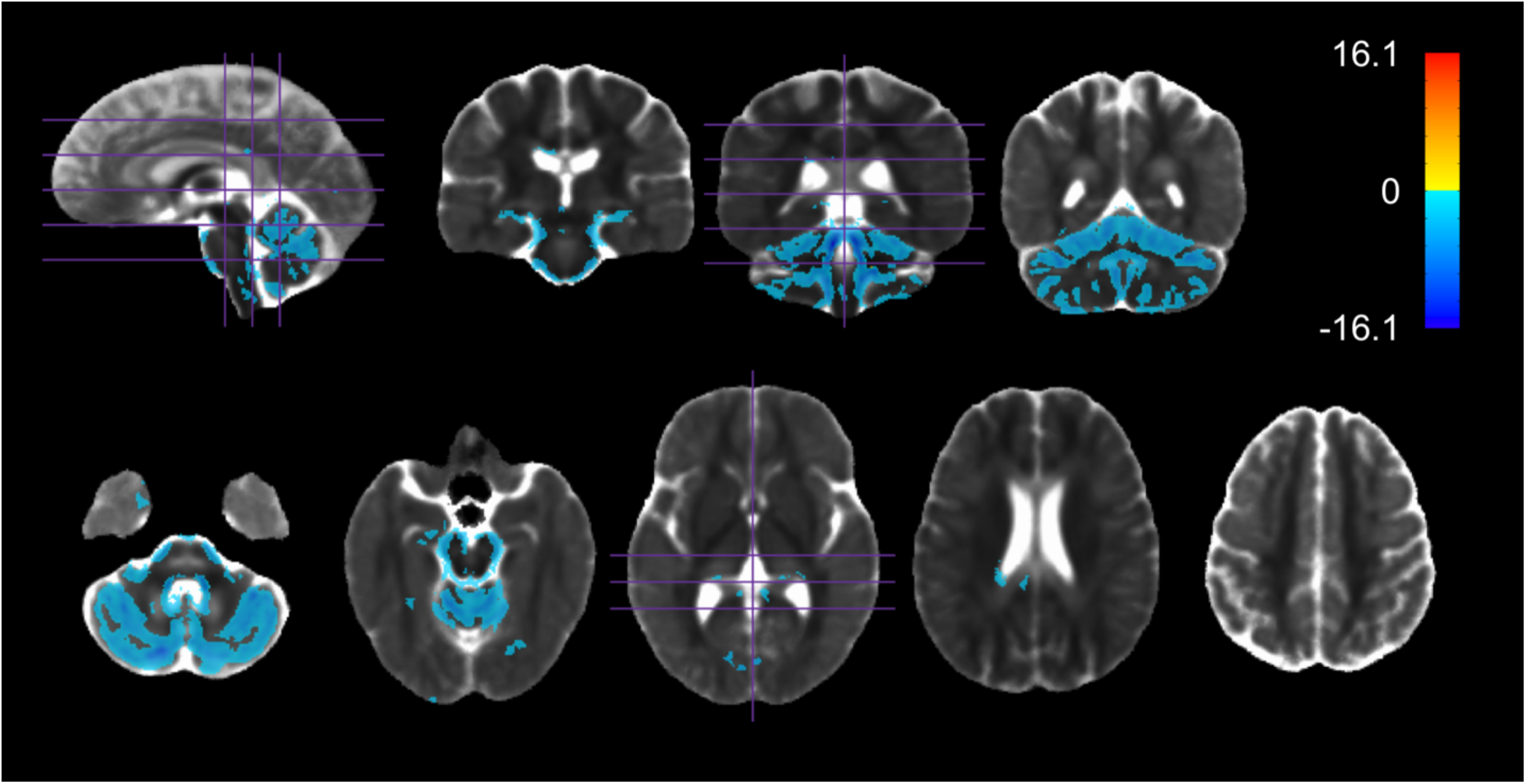
Regions in which pVF was significantly different between HVs and SCA7 *(p<*0.05, familywise error corrected). Values shown are the t-statistic. Blue indicates lower values (lower logJ/pVF) in SCA7 patients versus HVs. Orange indicates higher values in SCA7 patients versus HVs. Results are overlaid on the HV diffusion tensor template.

### 3.4. pMD reveals brain-wide microstructural abnormalities

SCA7 patients had a significantly higher pMD throughout the whole brain including the cerebellar peduncles, bilateral medial lemniscus, cerebral peduncles, corticospinal tracts, corpus callosum, internal capsules, external capsules, bilateral fornix/stria terminalis, bilateral posterior thalamic radiation, bilateral sagittal stratum, superior corona radiata, anterior corona radiata, posterior corona radiata, bilateral superior longitudinal fasciculus, bilateral superior fronto-occipital fasciculus, left uncinate fasciculus, cerebellar WM and GM, brainstem, thalamus, bilateral pallidum, bilateral amygdala, ventral diencephalon, and many cortical GM areas in all four lobes (Figure 3, Tables 5 and 6, and Tables S5.1, S5.2, S6.3.1 and S6.3.2 in Supplement 2). A large (>1 Hedge’s *g*) effect size was observed in the same regions (Figure S2.3 in Supplement 1).

**Table 5:**
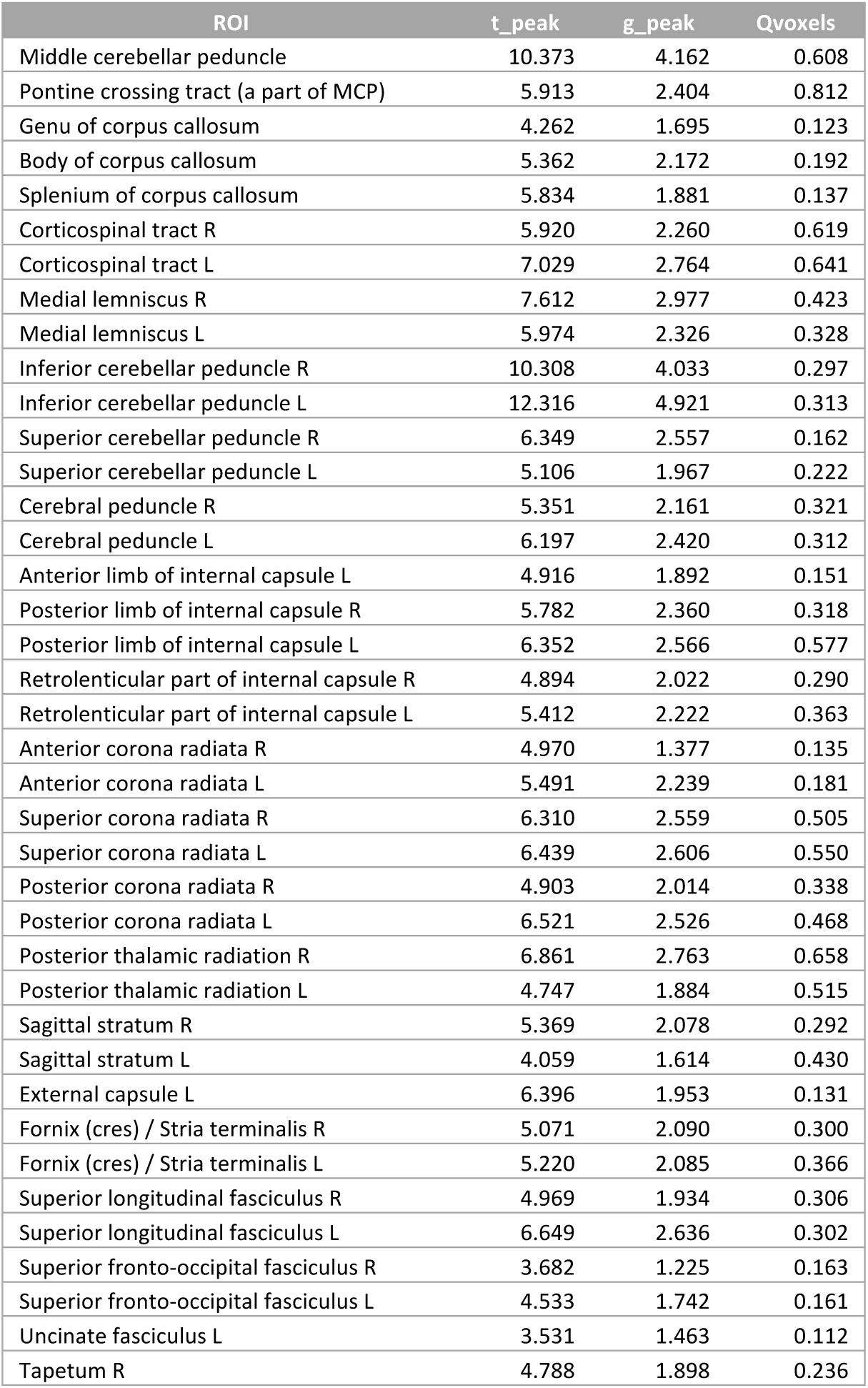
Peak *t*-statistic value, peak Hedge’s *g* value, and the fraction of ROI voxels significantly different between HVs and SCA7 patients (Qvoxels) for the pMD group comparison within ROIs of the ICBM-DTI-81 WM atlas. Only regions with Qvoxels ≥ 0.10 are shown.

**Table 6:**
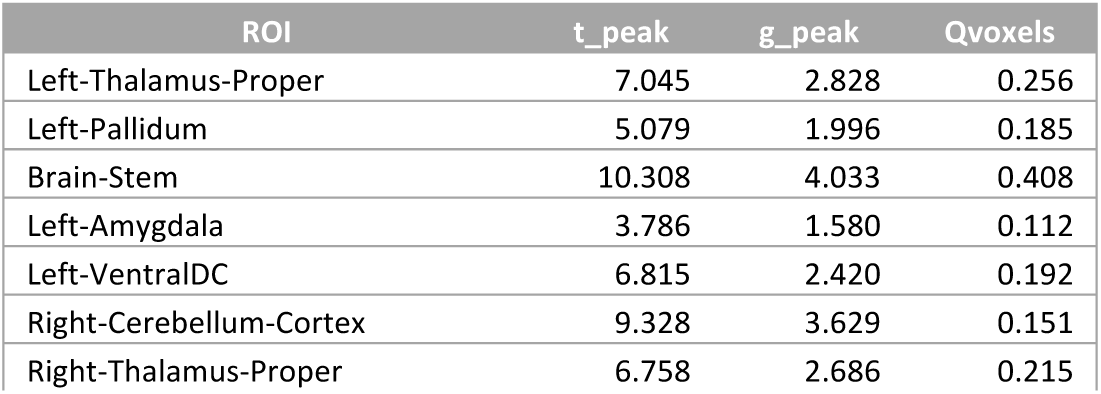
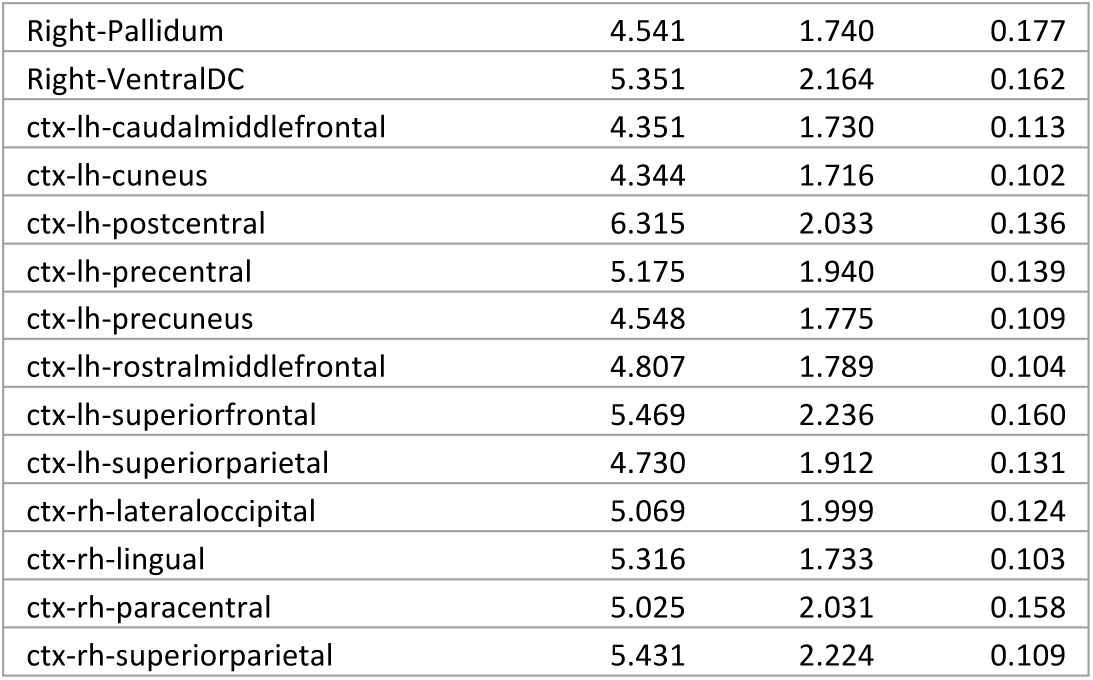
Peak *t*-statistic value, peak Hedge’s *g* value, and the fraction of ROI voxels significantly different between HVs and SCA7 patients (Qvoxels) for the pMD group comparison within ROIs of the FreeSurfer GM atlas. Only regions with Qvoxels ≥ 0.10 are shown.

| ROI | t_peak | g_peak | Qvoxels |
| --- | --- | --- | --- |
| Left-Thalamus-Proper | 7.045 | 2.828 | 0.256 |
| Left-Pallidum | 5.079 | 1.996 | 0.185 |
| Brain-Stem | 10.308 | 4.033 | 0.408 |
| Left-Amygdala | 3.786 | 1.580 | 0.112 |
| Left-VentralDC | 6.815 | 2.420 | 0.192 |
| Right-Cerebellum-Cortex | 9.328 | 3.629 | 0.151 |
| Right-Thalamus-Proper | 6.758 | 2.686 | 0.215 |
| Right-Pallidum | 4.541 | 1.740 | 0.177 |
| Right-VentralDC | 5.351 | 2.164 | 0.162 |
| ctx-lh-caudalmiddlefrontal | 4.351 | 1.730 | 0.113 |
| ctx-lh-cuneus | 4.344 | 1.716 | 0.102 |
| ctx-lh-postcentral | 6.315 | 2.033 | 0.136 |
| ctx-lh-precentral | 5.175 | 1.940 | 0.139 |
| ctx-lh-precuneus | 4.548 | 1.775 | 0.109 |
| ctx-lh-rostralmiddlefrontal | 4.807 | 1.789 | 0.104 |
| ctx-lh-superiorfrontal | 5.469 | 2.236 | 0.160 |
| ctx-lh-superiorparietal | 4.730 | 1.912 | 0.131 |
| ctx-rh-lateraloccipital | 5.069 | 1.999 | 0.124 |
| ctx-rh-lingual | 5.316 | 1.733 | 0.103 |
| ctx-rh-paracentral | 5.025 | 2.031 | 0.158 |
| ctx-rh-superiorparietal | 5.431 | 2.224 | 0.109 |

**Figure 3:**
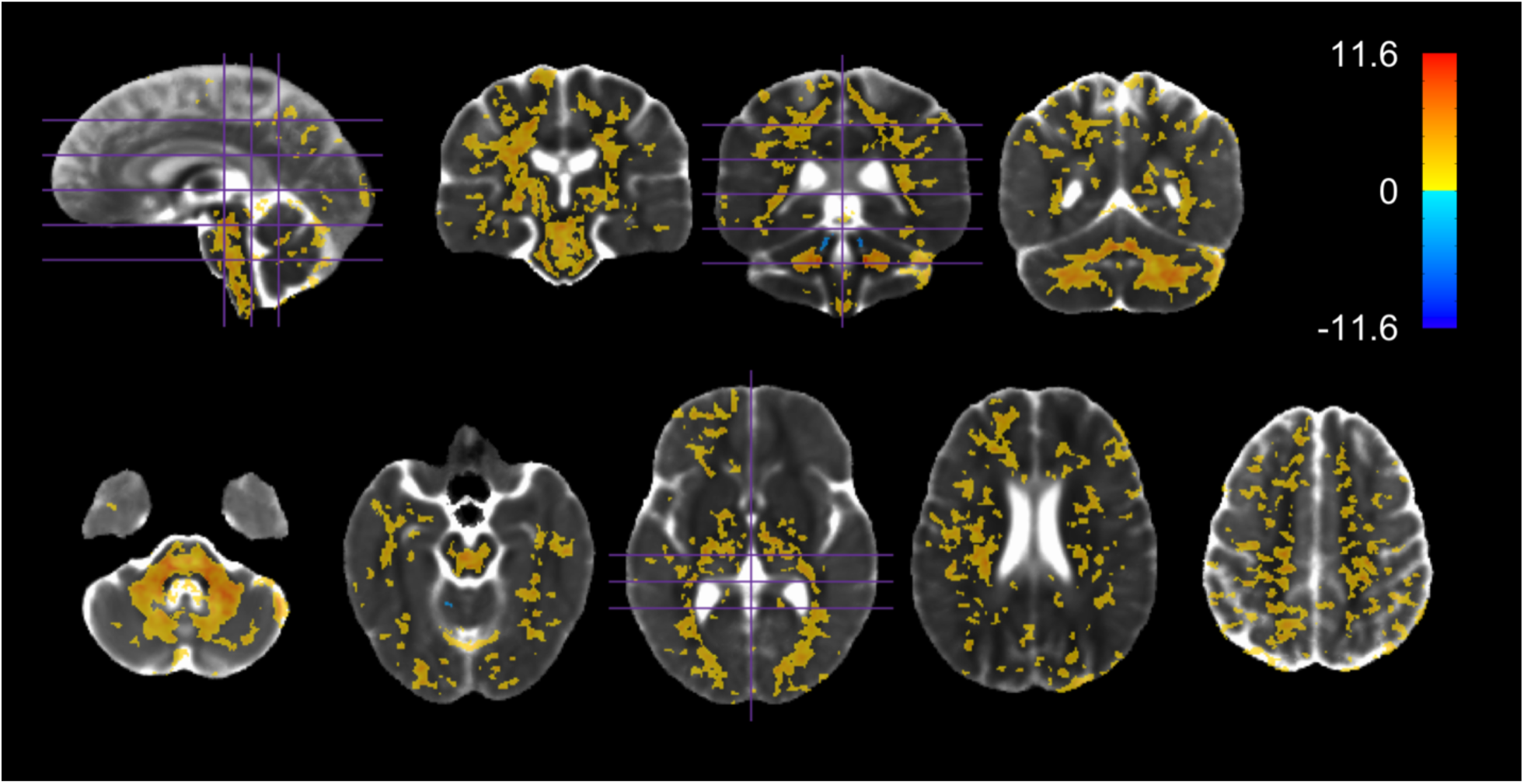
Regions in which pMD was significantly different between HVs and SCA7 *(p<*0.05, familywise error corrected). Values shown are the t-statistic. Blue indicates lower values (lower pMD/pFA) in SCA7 patients versus HVs. Orange indicates higher values in SCA7 patients versus HVs. Results are overlaid on the HV diffusion tensor template.

The MD was found to be significantly higher in SCA7 patients in most of the same regions that pMD was elevated (Figure S1.2, S2.6 and Tables S3.2.1, S3.2.2 in Supplement 1, and Tables S5.1, S5.2, S6.6.1, S6.6.2 in Supplement 2), but a some key differences were found between the two metrics. In contrast to the pMD, the MD was significantly greater in patients in almost every voxel of the cerebellar cortices and peduncles. Additionally, pMD, unlike the single-compartment MD, remained robust to decreases in pVF (Figure S4.2 in Supplement 1).

### 3.5. pFA reveals widespread WM microstructural abnormalities

SCA7 patients had a significantly lower pFA in almost the exact same WM and subcortical GM regions where pMD was found to be higher in patients (cerebellar peduncles, bilateral medial lemniscus, cerebral peduncles, corticospinal tracts, bilateral fornix/stria terminalis, internal capsules, external capsules, corpus callosum, bilateral posterior thalamic radiation, superior corona radiata, anterior corona radiata, posterior corona radiata, bilateral cingulum, bilateral superior longitudinal fasciculus, bilateral fronto-occipital fasciculus, bilateral sagittal stratum, cerebellar WM and GM, brainstem, thalamus, bilateral putamen, bilateral pallidum, and bilateral ventral diencephalon) (Figure 4, Tables 7 and 8, and Tables S5.1, S5.2, S6.4.1, S6.4.2 in Supplement 2). A large (>1 Hedge’s *g*) effect size was observed in the same regions (Figure S2.4 in Supplement 1).

**Table 7:**
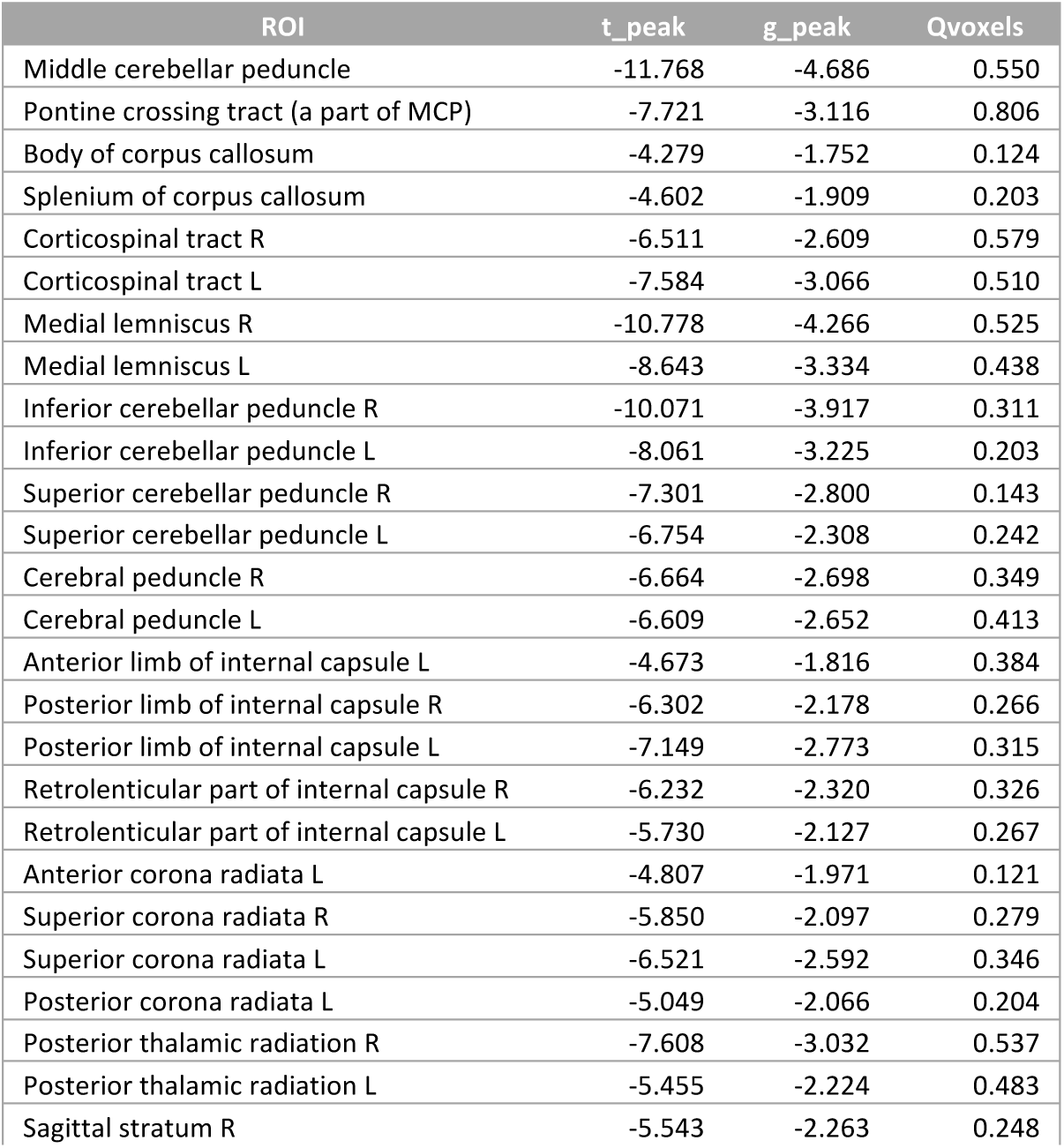
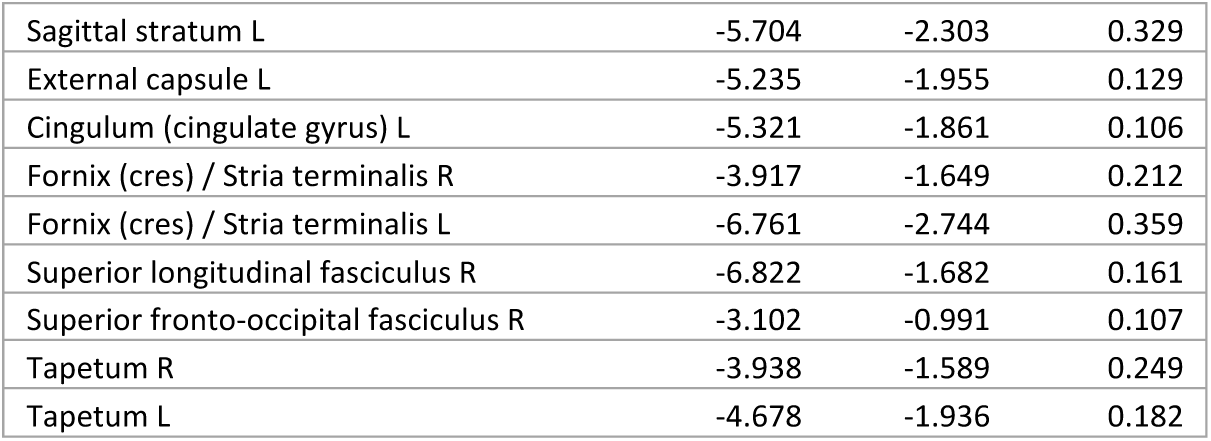
Peak *t*-statistic value, peak Hedge’s *g* value, and the fraction of ROI voxels significantly different between HVs and SCA7 patients (Qvoxels) for the pFA group comparison within ROIs of the ICBM-DTI-81 WM atlas. Only regions with Qvoxels ≥ 0.10 are shown.

**Table 8:**
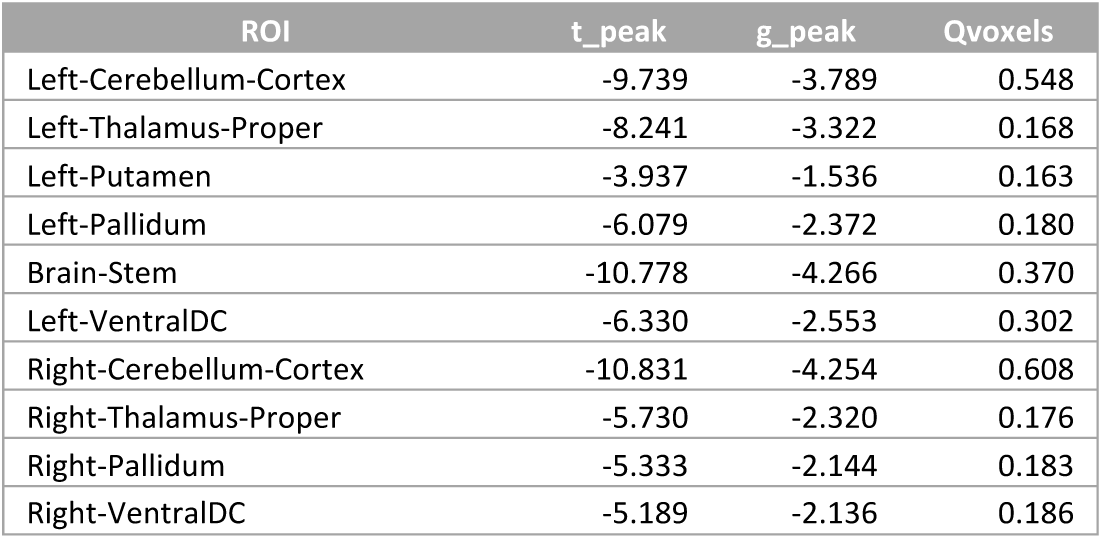
Peak *t*-statistic value, peak Hedge’s *g* value, and the fraction of ROI voxels significantly different between HVs and SCA7 patients (Qvoxels) for the pFA group comparison within ROIs of the FreeSurfer GM atlas. Only regions with Qvoxels ≥ 0.10 are shown.

| ROI | $t_{\text{peak}}$ | $g_{\text{peak}}$ | Qvoxels |
| --- | --- | --- | --- |
| Left-Cerebellum-Cortex | -9.739 | -3.789 | 0.548 |
| Left-Thalamus-Proper | -8.241 | -3.322 | 0.168 |
| Left-Putamen | -3.937 | -1.536 | 0.163 |
| Left-Pallidum | -6.079 | -2.372 | 0.180 |
| Brain-Stem | -10.778 | -4.266 | 0.370 |
| Left-VentralDC | -6.330 | -2.553 | 0.302 |
| Right-Cerebellum-Cortex | -10.831 | -4.254 | 0.608 |
| Right-Thalamus-Proper | -5.730 | -2.320 | 0.176 |
| Right-Pallidum | -5.333 | -2.144 | 0.183 |
| Right-VentralDC | -5.189 | -2.136 | 0.186 |

**Figure 4:**
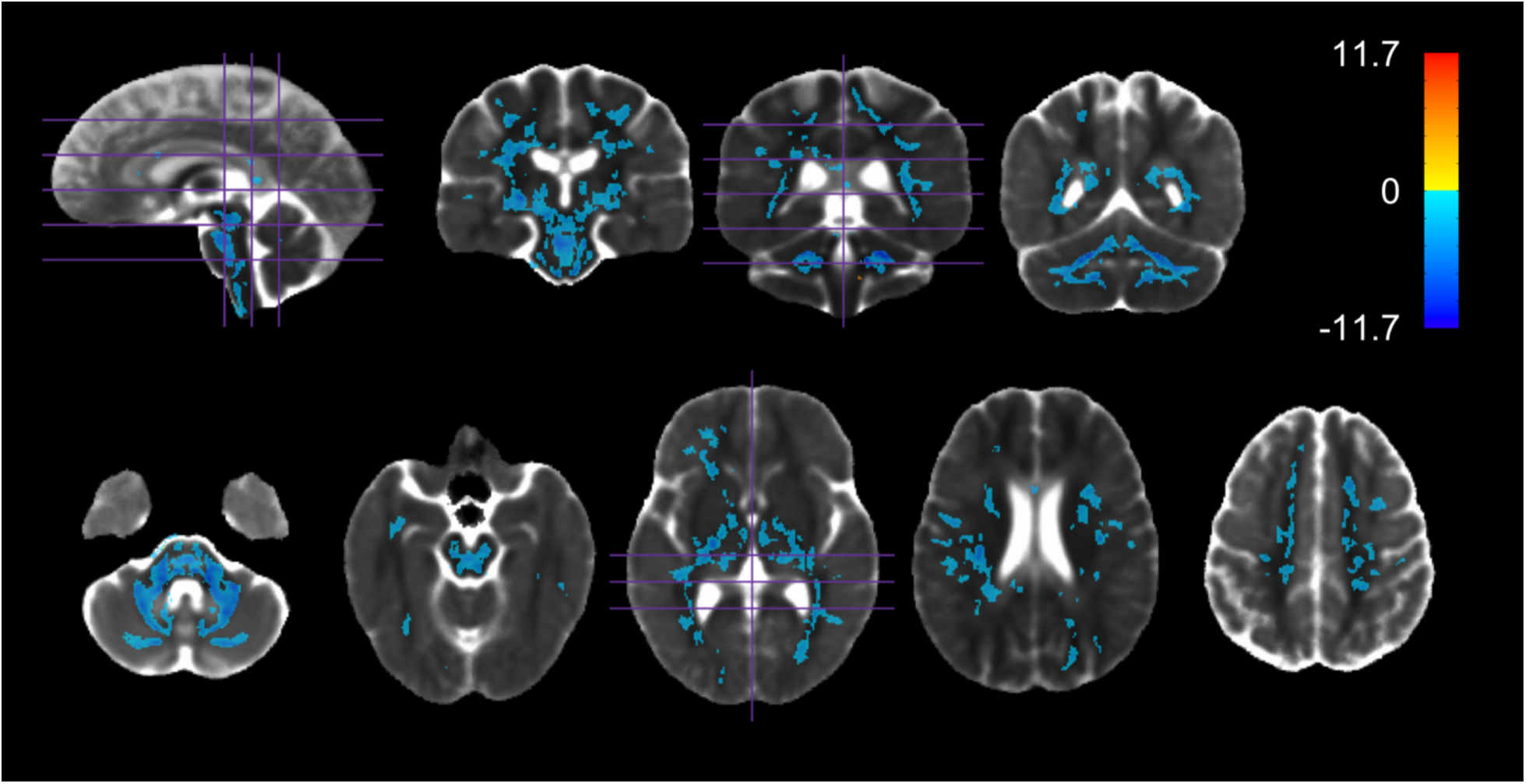
Regions in which pFA was significantly different between HVs and SCA7 *(p<*0.05, familywise error corrected). Values shown are the t-statistic. Blue indicates lower values (lower pMD/pFA) in SCA7 patients versus HVs. Orange indicates higher values in SCA7 patients versus HVs. Results are overlaid on the HV diffusion tensor template.

The FA was found to be significantly lower in most of the same regions (Figures S1.3 and S2.7, Table S3.3.1 and S3.3.2 in Supplement 1, Tables S5.1, S5.2, S6.7.1, S6.7.2 in Supplement 2). However, in contrast to the pFA, the FA was significantly lower in patients in almost every voxel of the cerebellar cortices and peduncles. Like the pMD, the pFA remained robust to decreases in pVF compared to the single-compartment FA (Figure S4.2 in Supplement 1).

### 3.6. Cerebellar pVF, Whole Brain pFA most strongly correlated with ataxia severity

We found three correlations that achieved the significance threshold of *p* < 0.05 (uncorrected): cerebellar pVF with SARA (*r* = −0.66, *p* = 0.014), cerebral pFA with SARA (*r* = −0.62, *p* = 0.024), and brainstem pFA with SARA (*r* = −0.57, *p* = 0.040). While none of these correlations survived corrections for multiple comparisons, we present them due to their high effect size and the fact that the threshold to achieve significance was stringent. Considering 12 correlations were assessed (4 metrics X 3 regions) with a sample size of 13 patients, any single correlation needed to have a Pearson’s *r* greater than 0.73 to be considered statistically significant (*p* < 0.05/12).

In contrast to the other metrics, the pFA was relatively well correlated with the SARA score in all three regions (brainstem – *r* = −0.57, *p* = 0.040; cerebellum – *r* = −0.54, *p* = 0.055; cerebrum – *r* = −0.62, *p* = 0.024). Thus, we decided to look at how the average pFA of the whole brain correlated with the SARA score. We found that whole brain pFA correlated more strongly with the SARA score (*r* = −0.64, *p* = 0.018) than any of the three subregions individually. Although this correlation also does not survive corrections for multiple corrections, we again present it due to the high effect size.

The correlations between whole brain pFA and cerebellar pVF with the SARA score can be found in figure 5. Scatter plots of all correlations assessed can be found in Figure S4.1 in Supplement 1. Inclusion of the patient without measurable ataxia had little effect on observed correlations (Figures S4.3 and S4.4 in Supplement 1).

**Figure 5:**
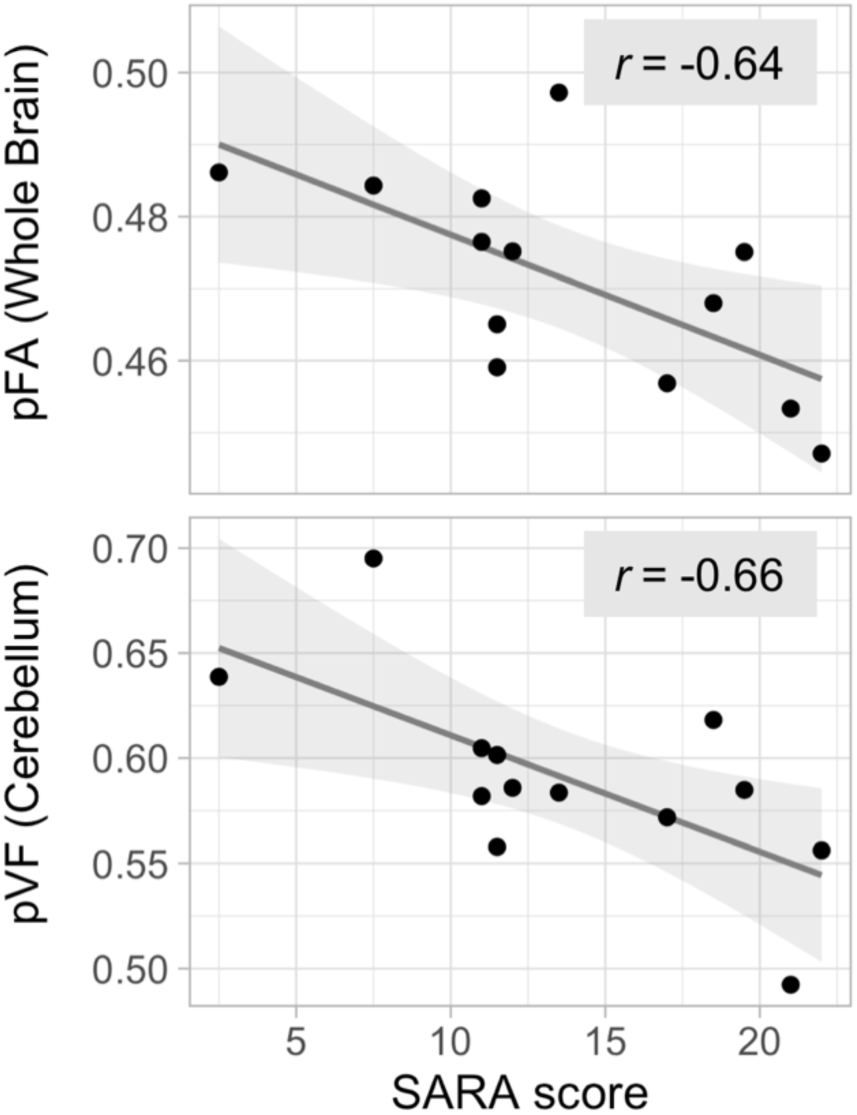
Scatter plots of whole brain pFA versus the SARA score (*r* = −0.64, *p* = 0.018, uncorrected) and cerebellar pVF versus the SARA score (*r* = −0.66, *p* = 0.014, uncorrected). Line of best fit and 95% confidence interval shown.

## 4. Discussion

In this study, we analyzed the brain-wide volumetric and microstructural abnormalities present in a cohort of SCA7 patients *in vivo* and evaluated their relationship to ataxia severity as measured by SARA. We used a recently developed diffusion tensor image registration technique ^25^ to perform voxel-wise analyses across all anatomical structures and tissue types. At the same time, we used a multi-shell DWI acquisition and dual-compartment DTI model ^27^ to account for potential increases in CSF-like free water due to neurodegeneration.

Using this approach, we found a distinction between the volumetric and microstructural abnormalities present in SCA7 patients. While volume loss was found primarily in the brainstem, cerebellum, thalamus, and a subset of cerebral WM tracts, microstructural abnormalities were detected brain-wide in WM and GM alike. This distinction was also meaningful in terms of ataxia severity. Among our imaging metrics, cerebellar pVF and global WM pFA were the most strongly associated with the SARA score. This suggests that cerebellar tissue loss and/or global WM microstructural abnormality may not only be involved in motor symptomology, but may potentially serve as biomarkers of SCA7 progression as well.

Since we were able to assess volume loss and microstructural abnormalities in both the GM and WM while controlling for the effects of significant tissue atrophy, we conclude that this is one of the most comprehensive *in vivo* assessments of neurodegeneration in SCA7 to date. In fact, the distinction we found between volumetric and microstructural abnormalities closely matches the general findings of post-mortem neuropathology – neuronal and axonal loss in primarily the brainstem and cerebellum, but molecular and cellular abnormalities brain-wide ^1, 3, 8-10^. Moreover, we were able to connect specific structural abnormalities to ataxia severity. Thus, we believe this approach holds promise for further study of SCA7 and other neurodegenerative diseases more generally.

### 4.1. Possible neurobiological correlates of imaging findings

logJ, which quantifies the degree of expansion and compression necessary to warp a subject’s brain to a template ^26^, is an excellent measure of relative volumetric differences between the two groups. Therefore, the significantly lower logJ in certain brain regions in patients is a good indicator of volume loss in those regions. This atrophy likely reflects the neuronal and axonal loss reported in post-mortem neuropathology ^1, 9, 10^.

pVF is a measure of the proportion of water in a given voxel belonging to the parenchymal compartment. As described in the Materials and Methods section, this proportion is assumed to be equal to the proportion of volume within the voxel comprised of parenchymal tissue. Thus, a decrease in this value indicates a decrease in the amount of parenchymal tissue coupled with a commensurate increase in the amount of extracellular space filled with CSF-like free water. Given that we primarily observe a decrease in the pVF in patients only around the CSF-tissue borders of structures that are atrophying, such as the cerebellum, we believe this metric reflects volume loss in areas where the tissue is retreating and being replaced by CSF space. Therefore, we interpret the observed decreases in pVF in patients as volumetric findings complementary to those revealed by DTBM, though only where tissue is being directly replaced by CSF space.

MD and FA differences can indicate the presence of various microstructural abnormalities. A simultaneous decrease in FA and increase in MD in the WM has been associated with many pathological changes including, but not limited to, axonal injury, demyelination, dysmyelination, edema, and inflammation (see ^21^ for a review). Neuropathology has revealed axonal loss in the WM of the brainstem and cerebellum in SCA7 patients ^1, 3, 9, 10^, so the observed increase in pMD and decrease in pFA in these areas may be a result of this ongoing degeneration of the WM. Astrogliosis has also been reported in almost all brain structures and tissues of SCA7 patients ^1, 9, 10^. Astrogliosis is characterized by a proliferation of astrocytes and morphological changes to them, including cellular and cytoskeletal hypertrophy ^47^. While one study found that reactive gliosis was associated with increases in FA in the cortex adjacent to lesion sites ^48^, the effect of widespread gliosis on FA and MD is unknown. Given the subtle cellular changes associated with astrogliosis, however, one would expect altered diffusivity in the affected tissues. Thus, the widespread increases in pMD and decreases in pFA observed in patients may be associated with astrogliosis, though further study is warranted to test such a connection.

### 4.2. Connection to clinical manifestations

Qualitatively, our results appear to be consistent with the clinical manifestation of the disease. We found severe atrophy of the brainstem and cerebellum, a pathology that is classically associated with many ataxic conditions ^11^. Given that we found a strong correlation between ataxia severity and tissue loss in the cerebellum specifically, we believe this pathology is heavily involved in motor symptoms. We also found volume loss in the thalamus and corticospinal tract, both of which are essential to motor function. Whether this volume loss is a direct result of the disease process or secondary to the drastic loss of innervation from the cerebellum and brainstem structures is unknown. Also unknown is whether this volume loss in the thalamus and corticospinal tract directly contributes to motor difficulties in patients.

Additionally, we noted microstructural abnormalities in many structures involved in motor and visual function including the cerebellum, brainstem, thalamus, corticospinal tract, sensorimotor cortices, optic radiation, and occipital cortices. However, the relevance of these findings to the symptomology of SCA7 is unclear. Microstructural abnormalities were also found in many non-motor/non-visual regions, namely miscellaneous frontal, parietal, and temporal GM and WM. Furthermore, global WM microstructural abnormality, as measured by pFA, correlated best with ataxia severity as compared to microstructural abnormality in the cerebrum, brainstem, or cerebellum alone. If the abnormality we are measuring directly led to the development of symptoms, we would reasonably expect to see more non-motor/non-visual symptoms. Given that pMD and pFA are nonspecific measures of tissue microstructure, it is certainly possible we are measuring changes resulting from multiple factors. Specifically, they might reflect tissue damage or loss in atrophied regions (cerebellum, brainstem, thalamus, corticospinal tract), but other more subtle changes in tissue properties in other regions. It is also possible that we are detecting an abnormality that is known to be present almost everywhere in the brain, such as astrogliosis ^1, 8-10^. Such an abnormality would be unlikely to drive the clinical deficits observed in SCA7 due to its ubiquity, but it may still be valuable as a measure of disease progression.

### 4.3. DTBM versus VBM

To evaluate DTBM as a methodology for assessing volume in comparison to more conventional methods, we performed a group comparison of the GM maps. We found GM volume loss in all of the same regions that DTBM detected GM volume loss in (Figure S1.1 and Table S3.1.1 in Supplement 1, Tables S5.2 and S6.5.1 in Supplement 2). Unlike DTBM, however, VBM also found volume loss in the sensorimotor cortices. One possible explanation for this difference might be that DTBM relies on tensor-based registration to standard space while VBM uses scalar-based registration. Given that the information in these transformations is used to compute relative tissue volume for each method, differences in the transformations would lead to slightly different results. Another possible explanation is the fact that the magnitude of multiple comparison correction was smaller for the group comparison of the GM maps than for the group comparison of logJ, since GM maps were only compared in the GM while logJ was compared brain-wide. This gave VBM greater statistical power than DTBM, and thus may explain the additional finding in the sensorimotor cortices.

Other studies employing VBM to study SCA7 have reported volume loss in even more cortical regions ^13, 15, 16^. However, it is worth noting that two of these studies had a much greater sample size of patients (24+) ^15, 16^.

Although DTBM failed to detect volume loss in the sensorimotor cortices like our VBM analysis, it enabled us to accurately assess WM volume loss in vivo for the first time. This enabled us to detect volume loss in the cerebellar WM, cerebellar peduncles, brainstem, corticospinal tract, and other cerebral WM tracts. While volume loss in these WM tracts have already been reported in post-mortem studies, measuring it *in vivo* opens the door to better understanding how this pathology evolves during disease progression. Considering this, and the fact that DTBM detected volume loss in most of the GM regions that VBM did, we believe DTBM is the preferable method for assessing volume on the voxel-wise level.

### 4.4. Dual compartment diffusivity metrics versus single compartment diffusivity metrics

To evaluate pMD and pFA as measures of tissue microstructural abnormality as compared to the standard single-compartment DTI diffusivity metrics, we performed group comparisons of the MD and FA. We found that the MD and FA were abnormal in most of the same regions that the pMD and pFA were abnormal in (Figures S1.2 and S1.3, Tables S3.2.1, S3.2.2, S3.3.1, and S3.3.2 in Supplement 1, Tables S5.1, S5.2, S6.6.1, S6.6.2, S6.7.1, and S6.7.2 in Supplement 2). However, the MD differed from its parenchymal counterpart by being larger in patients in almost every voxel of the cerebellar cortices and peduncles. Similarly, the FA differed from the pFA by also being smaller in patients in almost every voxel of the cerebellar peduncles. Other studies using DTI to study SCA7 corroborated this, finding that the MD and FA were abnormal in the same regions we found the single-compartment MD and FA to be abnormal in ^13, 19^.

However, using the dual-compartment model, we know that the pVF was found to be lower in the cerebellar GM and the cerebellar peduncles. Thus, we believe that the observed abnormality in the MD and FA in these regions largely reflects an increase in CSF-like free water rather than a microstructural abnormality in the tissues themselves. Given this ability to separate out the effect of CSF-like free water, we believe the dual-compartment approach we employed is more appropriate for measuring and interpreting the structural abnormalities present in patients.

### 4.5. Limitations

Although there was no statistically significant difference in the age of the SCA7 patients versus HVs, the mean difference might still be considered large (36.4 years for SCA7 vs 44.5 years for HV). We addressed this concern by including age as a covariate in all group comparisons performed. Some of the effects that we observe in patients (decreased GM volume, increased WM MD, decreased WM FA) are actually associated with increasing age ^49, 50^, meaning we would expect the differences between our SCA7 and HV cohorts to be greater if the ages were better matched. Regardless, we would expect the results to be more accurate with better age matching.

Another limitation of our analysis was our small sample size (14 HVs, 13 SCA7). While limiting the power of our statistical testing, we verified that the reported results have a large effect size, as indicated by a Hedge’s *g* > 1. Nevertheless, this likely explains why we generally found structural abnormalities in fewer brain regions compared to other imaging studies. Furthermore, this small sample size of patients led us to test correlations between our imaging metrics and the SARA score across macroscopic regions rather than on the voxel-wise level.

One limitation that is not just specific to our study is the inability to identify the exact structural change underlying abnormalities in the DTI diffusion metrics (FA, MD). While the general consensus is that abnormalities in these metrics indicates microstructural abnormalities, they are associated with many different pathologies including, but not limited to, axonal injury, demyelination, dysmyelination, edema, and inflammation ^21^. Thus, as previously mentioned, we do not know for sure why we observe an increased parenchymal MD and decreased parenchymal FA in patients throughout much of the brain. Until the specific neurobiological correlate is established, it will remain challenging to understand the relevance of this finding to the pathophysiology of SCA7.

### 4.6. Extensions

Given that we were able to recapitulate the general findings of neuropathology *in vivo* and relate specific structural abnormalities to ataxia severity, we believe the approach employed here holds considerable promise to further study of SCA7. In particular, this approach could be applied to a longitudinal analysis of a cohort of SCA7 patients. Such an analysis has the potential to characterize the full spatiotemporal pattern of structural changes occurring during SCA7 progression. Specific structural changes could potentially be linked to the progression of specific clinical deficits, thereby increasing our understanding of SCA7 pathophysiology. Ultimately, the insights gained from such an analysis may enable the development of highly specific biomarkers of SCA7 progression that can be used in future treatment studies.

Pertinent to this last point, we identified two measures, cerebellar tissue volume and global WM microstructural abnormality, that correlated well with ataxia severity in this cohort. These measures would serve as a good starting point in the search for imaging biomarkers of SCA7 progression.

## 5. Conclusion

We used two recently developed DTI methodologies to assess both volumetric and microstructural abnormalities brain-wide in a cohort of SCA7 patients. These methods revealed features comparable to those found by neuropathology, namely severe volume loss restricted to the cerebellum, brainstem, thalamus, and a subset of cerebral WM tracts, but microstructural abnormalities in the GM and WM brain wide. This distinction was also meaningful in terms of ataxia severity, as we found that the SARA score was most strongly correlated with cerebellar tissue loss and global WM microstructural abnormality. Given these findings, we believe the approach employed here could be utilized in a longitudinal analysis to identify imaging biomarkers of SCA7 progression.

## Data Availability

The data that support the findings of this study are available from the corresponding author upon reasonable request.

## Abbreviations

DTBM: diffusion tensor-based morphometry
DTI: diffusion tensor imaging
DWI: diffusion weighted imaging
FA: fractional anisotropy
GM: gray matter
HV: healthy volunteer
logJ: natural log of the Jacobian determinant
MD: mean diffusivity
MNI: Montreal Neurological Institute
MRI: magnetic resonance imaging
pFA: parenchymal fractional anisotropy
pMD: parenchymal mean diffusivity
pVF: parenchymal volume fraction
Qvoxels: fraction of voxels
SARA: Scale for the Assessment and Rating of Ataxia
SCA7: Spinocerebellar Ataxia type 7
TICV: total intracranial volume
VBM: voxel-based morphometry
WM: white matter

## Appendices

Supplementary Information 1 of 2

Supplementary Information 2 of 2

