## Supplementary Material 1 of 2 for "*In vivo* assessment of neurodegeneration in Spinocerebellar Ataxia type 7"

### Supplementary Materials 1 of 2

#### Contents

**S1: VBM, MD, and FA group comparisons**
**Figure S1.1:** VBM group comparison
**Figure S1.2:** MD group comparison
**Figure S1.3:** FA group comparison

**S2: Group comparison effect size maps**
**Figure S2.1:** DTBM (logJ) group comparison effect size map
**Figure S2.2:** pVF group comparison effect size map
**Figure S2.3:** pMD group comparison effect size map
**Figure S2.4:** pFA group comparison effect size map
**Figure S2.5:** VBM group comparison effect size map
**Figure S2.6:** MD group comparison effect size map
**Figure S2.7:** FA group comparison effect size map

**S3: VBM, MD, and FA group comparison tables of most affected regions**
**S3.1:** VBM table
**Table S3.1.1:** VBM FreeSurfer GM atlas
**S3.2:** MD tables
**Table S3.2.1:** MD FreeSurfer GM atlas
**Table S3.2.2:** MD ICBM-DTI-81 WM atlas
**S3.3:** FA tables
**Table S3.3.1:** FA FreeSurfer GM atlas
**Table S3.3.2:** FA ICBM-DTI-81 WM atlas

**S4: Correlation analysis supplementary material**
**Figure S4.1:** Correlations between imaging metrics and SARA score
**Figure S4.2:** Correlations of single compartment diffusivity metrics (MD and FA) and dual compartment diffusivity metrics (pMD and pFA) with the parenchymal VF
**Figure S4.3:** Correlations of whole brain pFA and cerebellar pVF with the SARA score with asymptomatic patient included
**Figure S4.4:** Correlations between imaging metrics and SARA score with asymptomatic patient included

#### S1 VBM, MD, and FA group comparisons

**Figure S1.1:** VBM group comparison

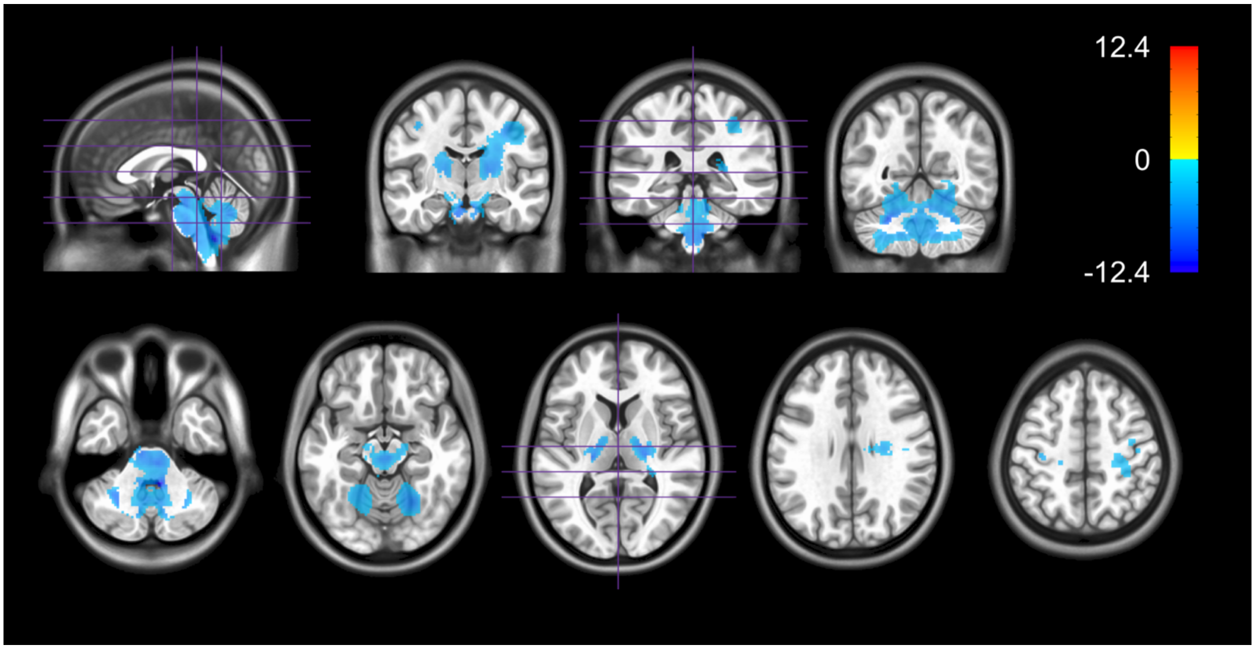


Figure S1.1: Regions in which GM volume was significantly different between HVs and SCA7 (*p*<0.05, familywise error corrected). Values shown are the *t*-statistic. Blue indicates a lower GM volume in SCA7 patients versus HVs. Orange indicates a higher GM volume. Results are overlaid on the MNI ICBM 2009c Nonlinear Asymmetric template.

**Figure S1.2:** MD group comparison

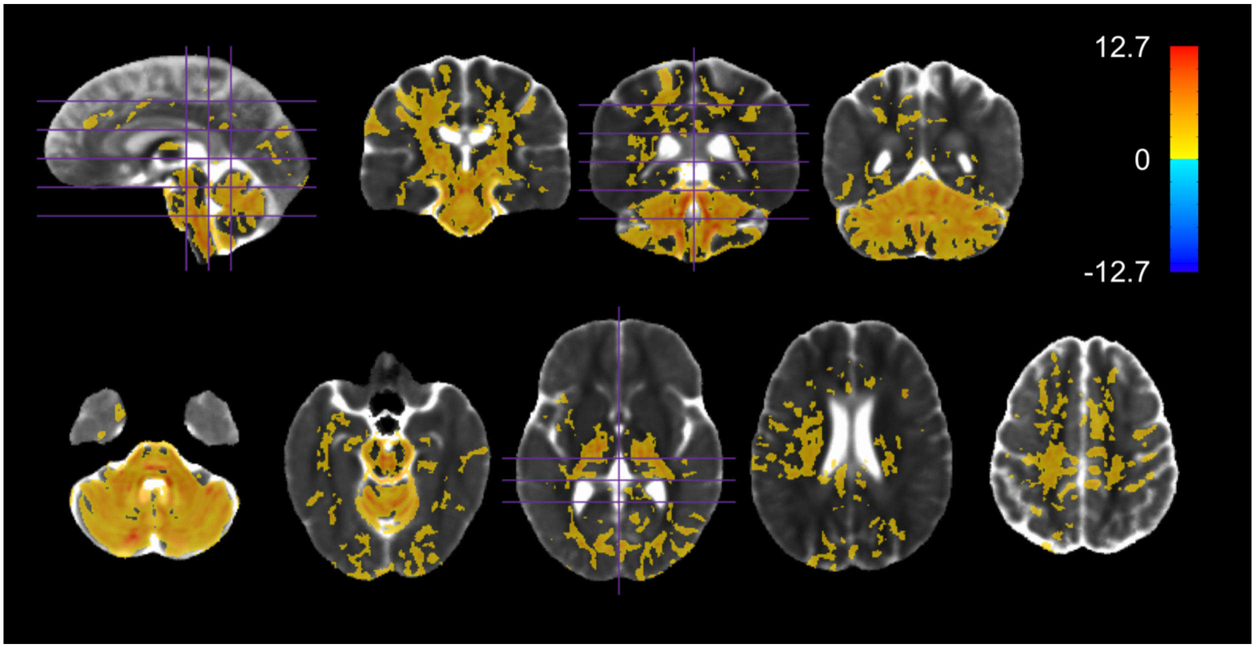


Figure S1.2: Regions in which MD was significantly different between HVs and SCA7 ($p$<0.05, familywise error corrected). Values shown are the $t$-statistic. Orange indicates a higher MD in SCA7 patients versus HVs. Blue indicates a lower MD. Results are overlaid on the HV diffusion tensor template.

**Figure S1.3:** FA group comparison

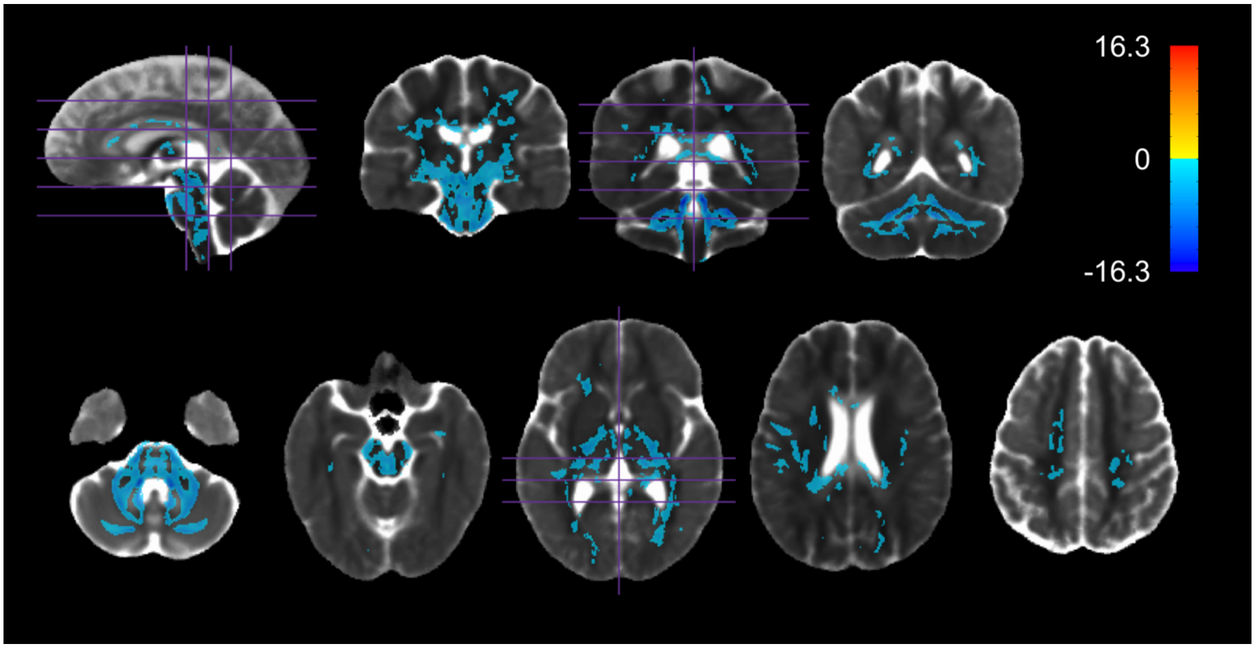


Figure S1.3: Regions in FA was significantly different between HVs and SCA7 ($p$<0.05, familywise error corrected). Values shown are the $t$-statistic. Blue indicates a lower FA in SCA7

patients versus HVs. Orange indicates a higher FA. Results are overlaid on the HV diffusion tensor template.

#### S2 Group comparison effect size maps

##### Figure S2.1 DTBM (logJ) group comparison effect size map

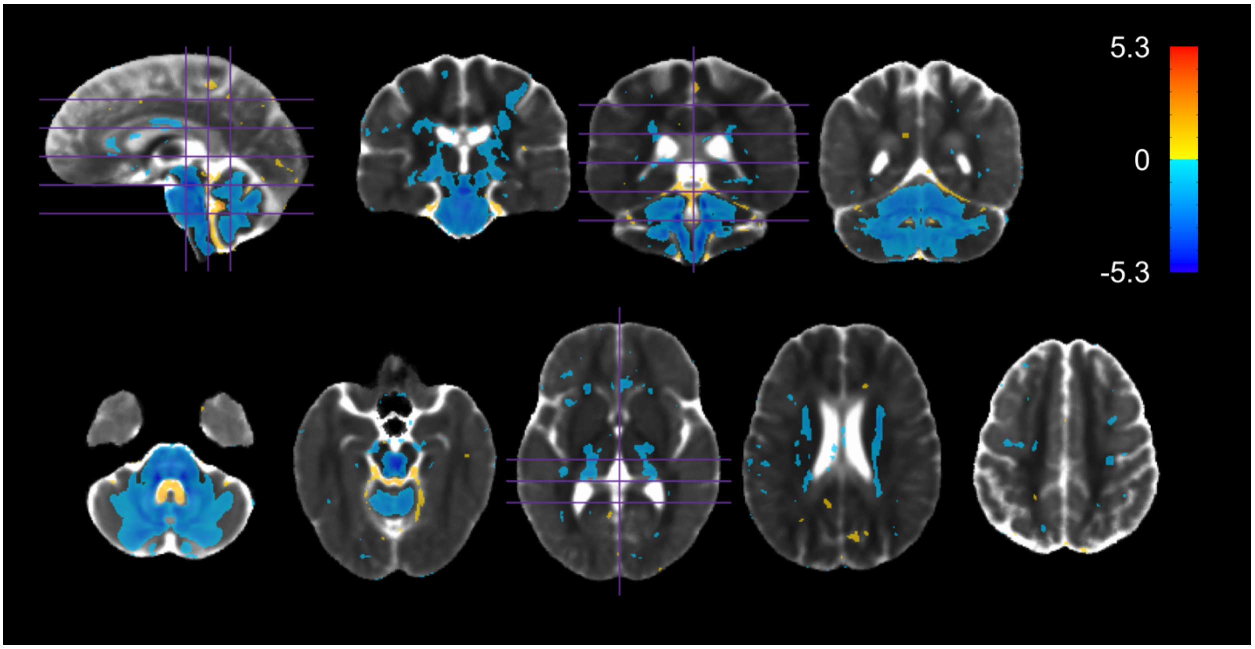


Figure S2.1: Regions in which the absolute value of Hedge's *g* for the logJ HV versus SCA7 comparison exceeded 1. Values shown are Hedge's *g*. Blue indicates a lower volume (lower logJ) in SCA7 patients versus HVs. Orange indicates a higher volume in SCA7 patients versus HVs. Results are overlaid on the HV diffusion tensor template.

##### Figure S2.2 pVF group comparison effect size map

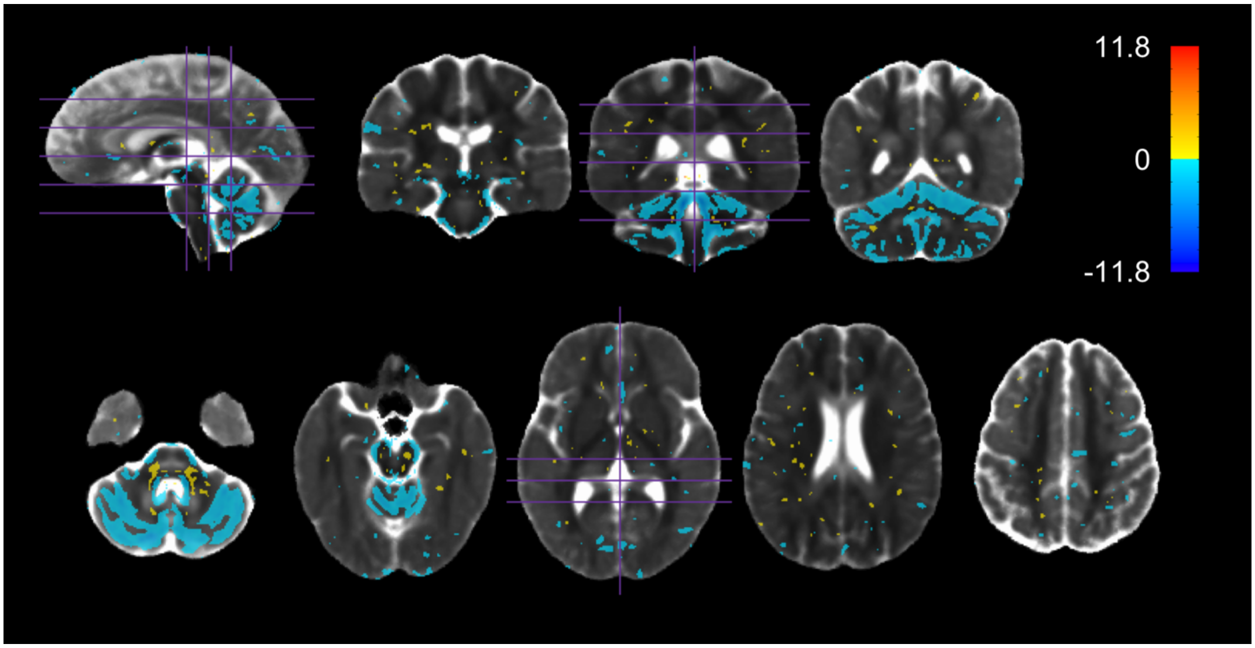


Figure S2.2: Regions in which the absolute value of Hedge's *g* for the pVF HV versus SCA7 comparison exceeded 1. Values shown are Hedge's *g*. Blue indicates a lower pVF in SCA7 patients versus HVs. Orange indicates a higher pVF in SCA7 patients versus HVs. Results are overlaid on the HV diffusion tensor template.

**Figure S2.3 pMD group comparison effect size map**

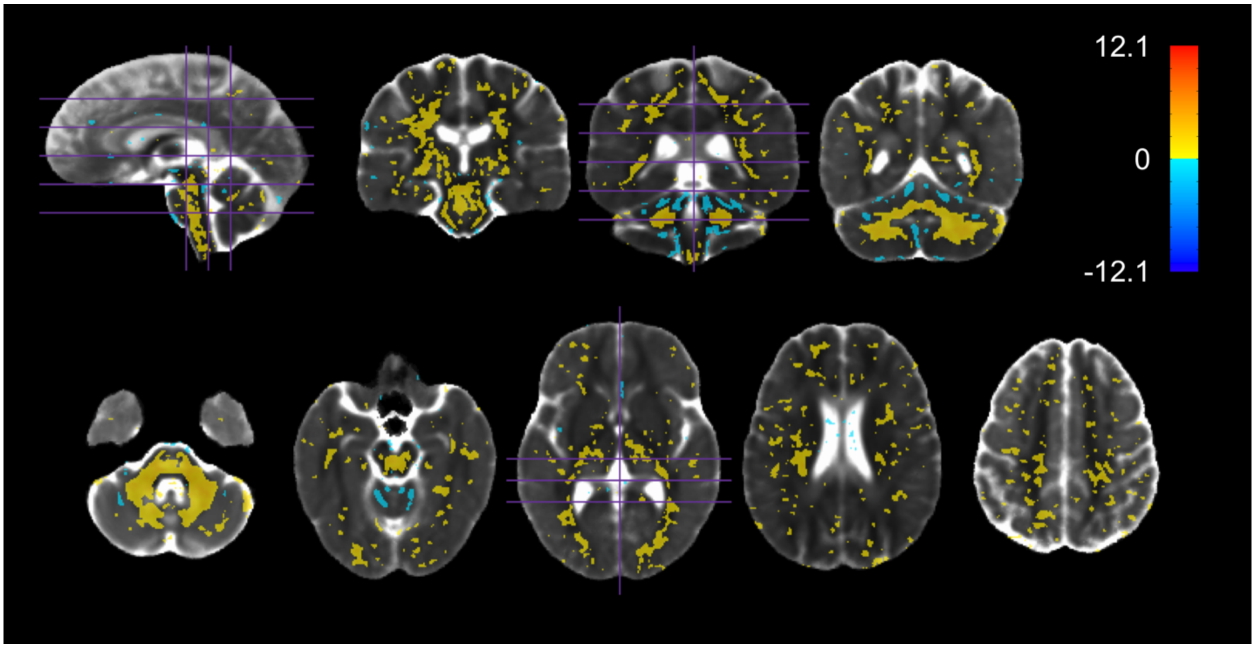


Figure S2.3: Regions in which the absolute value of Hedge's *g* for the pMD HV versus SCA7 comparison exceeded 1. Values shown are Hedge's *g*. Orange indicates a higher pMD in SCA7

patients versus HVs. Blue indicates a lower pMD in SCA7 patients versus HVs. Results are overlaid on the HV diffusion tensor template.

**Figure S2.4 pFA group comparison effect size map**

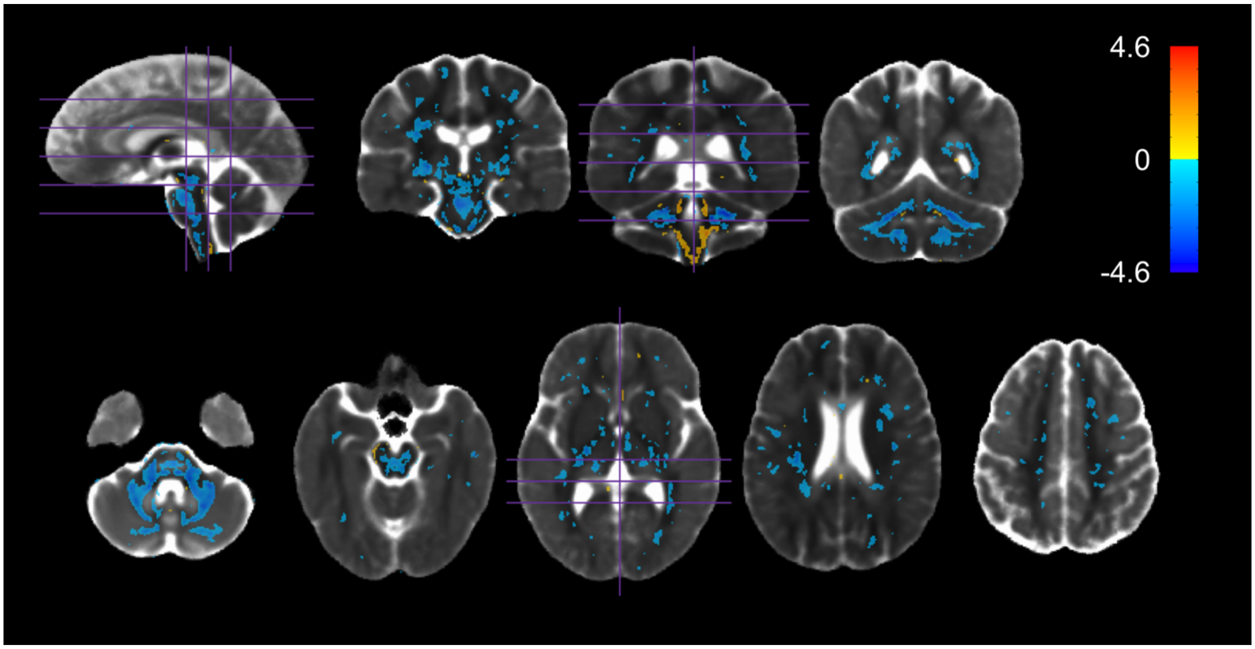


Figure S2.4: Regions in which the absolute value of Hedge's *g* for the pFA HV versus SCA7 comparison exceeded 1. Values shown are Hedge's *g*. Blue indicates a lower pFA in SCA7 patients versus HVs. Orange indicates a higher pFA in SCA7 patients versus HVs. Results are overlaid on the HV diffusion tensor template.

**Figure S2.5:** VBM group comparison effect size map

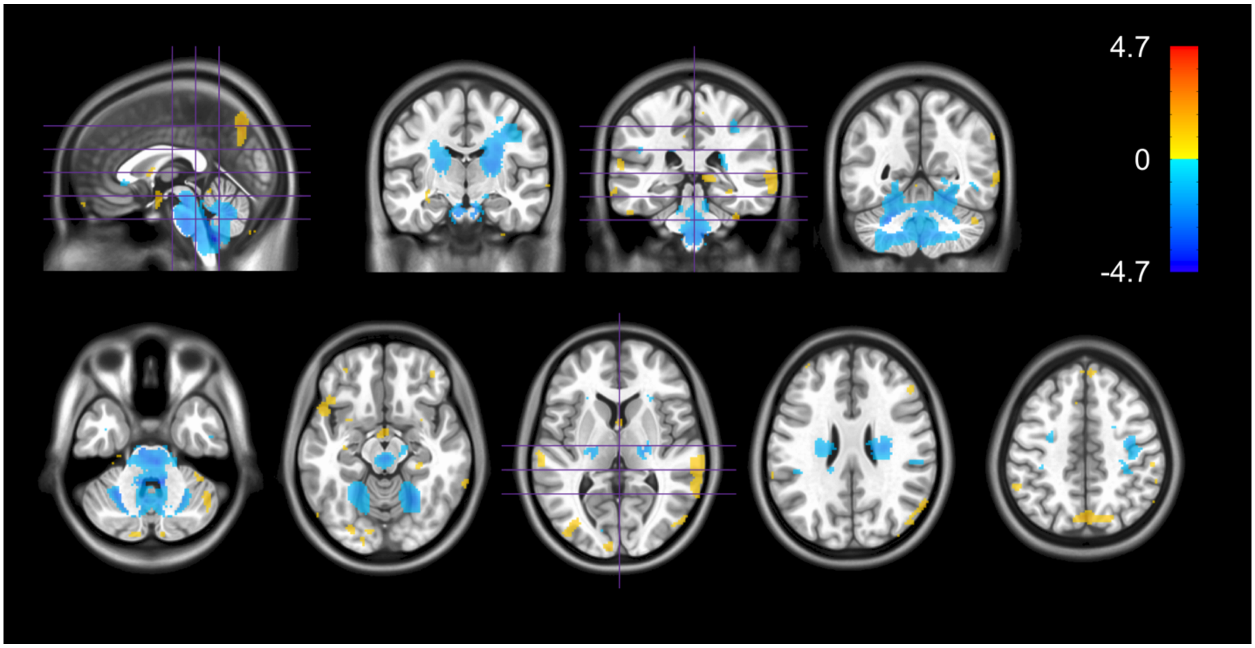


Figure S2.5: Regions in which the absolute value of Hedge's *g* for the VBM HV versus SCA7 comparison exceeded 1. Values shown are Hedge's *g*. Blue indicates a lower GM volume in SCA7 patients versus HVs. Orange indicates a higher GM volume. Results are overlaid on the MNI ICBM 2009c Nonlinear Asymmetric template.

**Figure S2.6:** MD group comparison effect size map

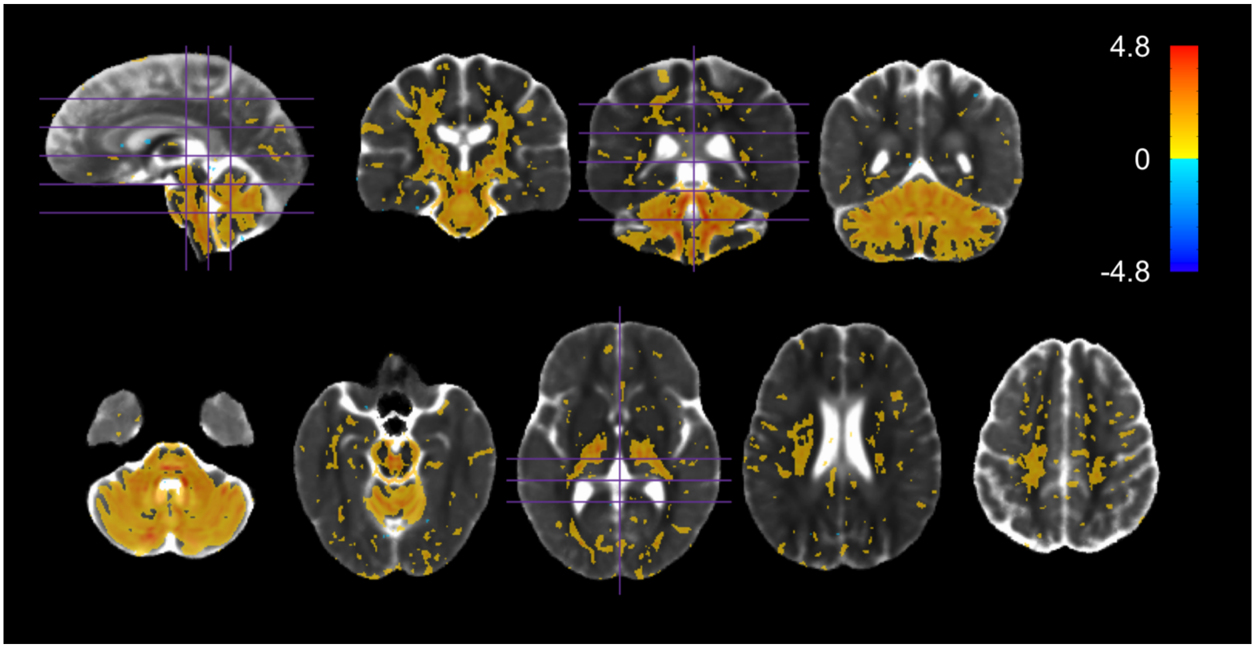


Figure S2.6: Regions in which the absolute value of Hedge's *g* for the MD HV versus SCA7 comparison exceeded 1. Values shown are Hedge's *g*. Orange indicates a higher MD in SCA7

patients versus HVs. Blue indicates a lower MD. Results are overlaid on the HV diffusion tensor template.

**Figure S2.7:** FA group comparison effect size map

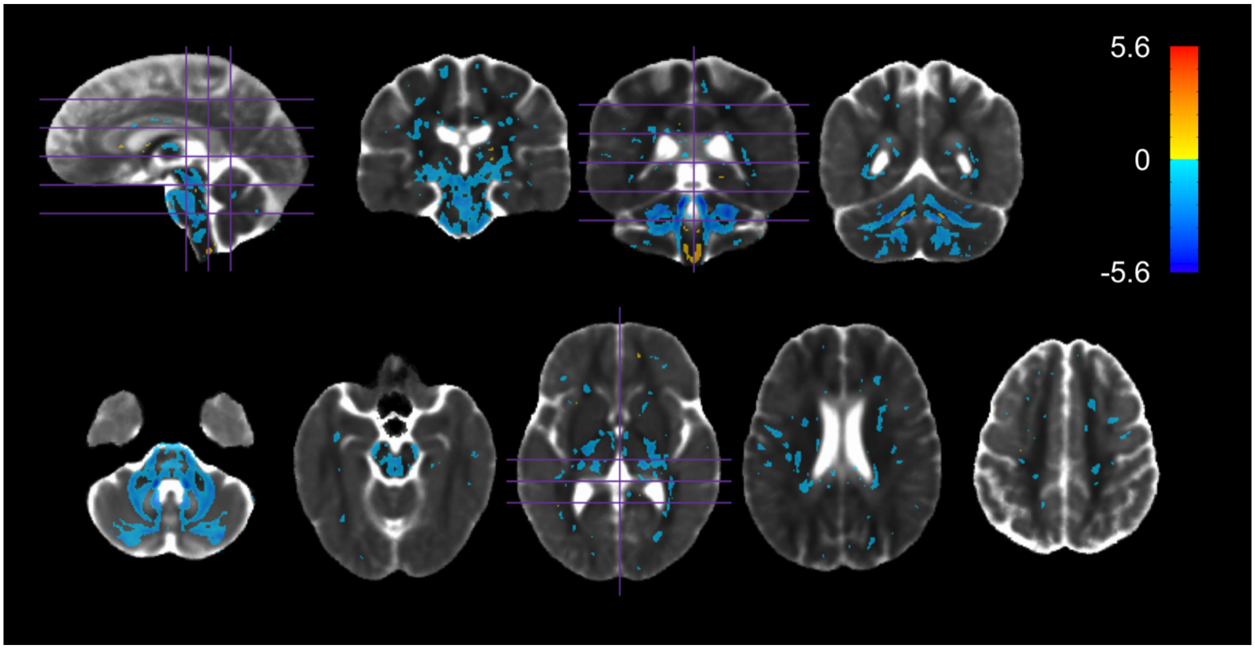


Figure S2.7: Regions in which the absolute value of Hedge's *g* for the FA HV versus SCA7 comparison exceeded 1. Values shown are Hedge's *g*. Blue indicates a lower FA in SCA7 patients versus HVs. Orange indicates a higher FA. Results are overlaid on the HV diffusion tensor template.

### S3: VBM, MD, and FA group comparison tables of most affected regions

#### S3.1: VBM tables

##### Table S3.1.1: VBM FreeSurfer GM atlas

| ROI | t_peak | g_peak | Qvoxels |
|---|---|---|---|
| Left-Cerebellum-Cortex | -11.242 | -4.199 | 0.282 |
| Left-Thalamus-Proper | -8.238 | -2.122 | 0.199 |
| Brain-Stem | -12.066 | -4.570 | 0.794 |
| Left-VentralDC | -6.790 | -2.338 | 0.177 |
| Right-Cerebellum-Cortex | -10.527 | -2.847 | 0.243 |
| Right-Thalamus-Proper | -7.765 | -3.067 | 0.259 |
| Right-VentralDC | -6.782 | -2.270 | 0.234 |
| ctx-rh-precentral | -6.153 | -1.760 | 0.107 |

Table S3.1.1: Peak *t*-statistic value, peak Hedge's *g* value, and the fraction of ROI voxels significantly different between HVs and SCA7 patients (Qvoxels) for the VBM group comparison within ROIs of the FreeSurfer GM atlas. Only regions with Qvoxels $\geq$ 0.10 are shown.

#### S3.2: MD tables

##### Table S3.2.1: MD ICBM-DTI-81 WM atlas

| ROI | t_peak | g_peak | Qvoxels |
|---|---|---|---|
| Middle cerebellar peduncle | 11.879 | 4.706 | 0.922 |
| Pontine crossing tract (a part of MCP) | 11.588 | 4.443 | 0.960 |
| Body of corpus callosum | 4.942 | 1.703 | 0.196 |
| Corticospinal tract R | 7.170 | 2.889 | 0.786 |
| Corticospinal tract L | 7.453 | 2.918 | 0.812 |
| Medial lemniscus R | 11.524 | 4.396 | 0.773 |
| Medial lemniscus L | 10.868 | 3.923 | 0.781 |
| Inferior cerebellar peduncle R | 11.249 | 4.508 | 0.903 |
| Inferior cerebellar peduncle L | 10.490 | 4.042 | 0.813 |
| Superior cerebellar peduncle R | 14.568 | 5.362 | 0.896 |
| Superior cerebellar peduncle L | 11.237 | 4.472 | 0.838 |
| Cerebral peduncle R | 7.082 | 2.862 | 0.538 |
| Cerebral peduncle L | 8.495 | 2.989 | 0.538 |
| Anterior limb of internal capsule L | 6.849 | 2.725 | 0.342 |
| Posterior limb of internal capsule R | 6.621 | 2.688 | 0.647 |
| Posterior limb of internal capsule L | 8.300 | 3.254 | 0.772 |
| Retrolenticular part of internal capsule R | 5.996 | 2.429 | 0.369 |
| Retrolenticular part of internal capsule L | 6.082 | 2.457 | 0.330 |
| Anterior corona radiata L | 4.343 | 1.607 | 0.110 |
| Superior corona radiata R | 6.012 | 2.440 | 0.356 |
| Superior corona radiata L | 6.240 | 2.372 | 0.735 |
| Posterior corona radiata R | 4.441 | 1.833 | 0.280 |
| Posterior corona radiata L | 4.918 | 1.982 | 0.424 |
| Posterior thalamic radiation R | 4.221 | 1.759 | 0.211 |
| Posterior thalamic radiation L | 4.808 | 1.854 | 0.290 |
| Sagittal stratum R | 3.957 | 1.587 | 0.239 |
| Sagittal stratum L | 4.260 | 1.773 | 0.449 |

| | | | |
|---|---|---|---|
| External capsule L | 5.381 | 2.086 | 0.181 |
| Cingulum (cingulate gyrus) R | 4.087 | 1.627 | 0.148 |
| Cingulum (cingulate gyrus) L | 5.324 | 1.946 | 0.299 |
| Cingulum (hippocampus) R | 4.182 | 1.697 | 0.199 |
| Fornix (cres) / Stria terminalis R | 5.883 | 2.396 | 0.696 |
| Fornix (cres) / Stria terminalis L | 7.055 | 2.816 | 0.669 |
| Superior longitudinal fasciculus R | 3.995 | 1.620 | 0.115 |
| Superior longitudinal fasciculus L | 4.862 | 1.990 | 0.331 |
| Superior fronto-occipital fasciculus R | 3.528 | 1.497 | 0.110 |
| Superior fronto-occipital fasciculus L | 3.707 | 1.509 | 0.268 |
| Uncinate fasciculus L | 5.660 | 2.171 | 0.378 |

Table S3.2.1: Peak *t*-statistic value, peak Hedge's *g* value, and the fraction of ROI voxels significantly different between HVs and SCA7 patients (Qvoxels) for the MD group comparison within ROIs of the ICBM-DTI-81 WM atlas. Only regions with Qvoxels ≥ 0.10 are shown.

**Table S3.2.2:** MD FreeSurfer GM atlas

| ROI | t_peak | g_peak | Qvoxels |
|---|---|---|---|
| Left-Cerebellum-Cortex | 11.504 | 4.558 | 0.807 |
| Left-Thalamus-Proper | 8.300 | 3.254 | 0.454 |
| Left-Putamen | 5.534 | 2.100 | 0.127 |
| Left-Pallidum | 4.688 | 1.939 | 0.103 |
| Brain-Stem | 14.568 | 5.362 | 0.742 |
| Left-Hippocampus | 5.023 | 1.876 | 0.192 |
| Left-Amygdala | 4.554 | 1.641 | 0.115 |
| Left-VentralDC | 7.482 | 2.965 | 0.442 |
| Right-Cerebellum-Cortex | 11.628 | 4.418 | 0.815 |
| Right-Thalamus-Proper | 9.102 | 3.659 | 0.463 |
| Right-Pallidum | 4.457 | 1.778 | 0.123 |
| Right-Hippocampus | 4.651 | 1.847 | 0.154 |
| Right-VentralDC | 7.944 | 3.033 | 0.417 |
| ctx-lh-caudalanteriorcingulate | 4.777 | 1.654 | 0.262 |
| ctx-lh-cuneus | 5.405 | 2.072 | 0.161 |
| ctx-lh-entorhinal | 4.981 | 1.967 | 0.210 |
| ctx-lh-fusiform | 5.619 | 2.188 | 0.129 |
| ctx-lh-isthmuscingulate | 4.514 | 1.517 | 0.134 |
| ctx-lh-lateraloccipital | 5.871 | 2.003 | 0.139 |
| ctx-lh-lingual | 5.089 | 2.079 | 0.159 |
| ctx-lh-paracentral | 4.604 | 1.594 | 0.117 |
| ctx-lh-pericalcarine | 4.846 | 1.801 | 0.358 |
| ctx-lh-postcentral | 5.091 | 1.735 | 0.119 |
| ctx-lh-posteriorcingulate | 4.942 | 1.611 | 0.194 |
| ctx-lh-precuneus | 6.319 | 1.701 | 0.123 |
| ctx-lh-insula | 5.865 | 2.171 | 0.147 |
| ctx-rh-caudalanteriorcingulate | 4.283 | 1.631 | 0.182 |
| ctx-rh-cuneus | 5.312 | 2.094 | 0.197 |
| ctx-rh-fusiform | 7.262 | 2.575 | 0.163 |
| ctx-rh-isthmuscingulate | 4.400 | 1.595 | 0.226 |
| ctx-rh-lateraloccipital | 5.761 | 2.188 | 0.233 |
| ctx-rh-lingual | 6.828 | 1.905 | 0.137 |
| ctx-rh-parahippocampal | 5.229 | 2.113 | 0.185 |
| ctx-rh-paracentral | 7.162 | 2.837 | 0.217 |
| ctx-rh-pericalcarine | 6.305 | 2.230 | 0.347 |

| | | | |
|---|---|---|---|
| ctx-rh-posteriorcingulate | 5.867 | 1.645 | 0.228 |

Table S3.2.2: Peak *t*-statistic value, peak Hedge's *g* value, and the fraction of ROI voxels significantly different between HVs and SCA7 patients (Qvoxels) for the MD group comparison within ROIs of the FreeSurfer GM atlas. Only regions with Qvoxels ≥ 0.10 are shown.

**S3.3:** FA tables

**Table S3.3.1:** FA ICBM-DTI-81 WM atlas

| ROI | t_peak | g_peak | Qvoxels |
|---|---|---|---|
| Middle cerebellar peduncle | -14.007 | -4.536 | 0.740 |
| Pontine crossing tract (a part of MCP) | -7.921 | -2.995 | 0.910 |
| Body of corpus callosum | -4.918 | -1.702 | 0.145 |
| Splenium of corpus callosum | -4.652 | -1.872 | 0.163 |
| Corticospinal tract R | -6.023 | -2.445 | 0.675 |
| Corticospinal tract L | -7.951 | -2.969 | 0.626 |
| Medial lemniscus R | -10.257 | -3.596 | 0.884 |
| Medial lemniscus L | -10.361 | -3.718 | 0.819 |
| Inferior cerebellar peduncle R | -13.507 | -4.956 | 0.671 |
| Inferior cerebellar peduncle L | -12.553 | -5.002 | 0.515 |
| Superior cerebellar peduncle R | -16.047 | -5.455 | 0.929 |
| Superior cerebellar peduncle L | -16.335 | -5.591 | 0.892 |
| Cerebral peduncle R | -7.620 | -2.961 | 0.456 |
| Cerebral peduncle L | -8.831 | -3.276 | 0.603 |
| Anterior limb of internal capsule L | -6.877 | -2.755 | 0.443 |
| Posterior limb of internal capsule R | -6.265 | -2.436 | 0.268 |
| Posterior limb of internal capsule L | -6.649 | -2.342 | 0.297 |
| Retrolenticular part of internal capsule R | -5.508 | -1.831 | 0.411 |
| Retrolenticular part of internal capsule L | -5.227 | -1.914 | 0.196 |
| Anterior corona radiata L | -5.115 | -1.628 | 0.134 |
| Superior corona radiata R | -4.913 | -1.915 | 0.103 |
| Superior corona radiata L | -5.299 | -2.115 | 0.282 |
| Posterior thalamic radiation R | -4.702 | -1.945 | 0.432 |
| Posterior thalamic radiation L | -5.519 | -2.250 | 0.378 |
| Sagittal stratum R | -4.356 | -1.790 | 0.193 |
| Sagittal stratum L | -4.704 | -1.945 | 0.276 |
| Fornix (cres) / Stria terminalis R | -7.278 | -2.946 | 0.553 |
| Fornix (cres) / Stria terminalis L | -7.380 | -2.972 | 0.574 |
| Tapetum R | -3.923 | -1.621 | 0.288 |
| Tapetum L | -4.273 | -1.713 | 0.282 |

Table S3.3.1: Peak *t*-statistic value, peak Hedge's *g* value, and the fraction of ROI voxels significantly different between HVs and SCA7 patients (Qvoxels) for the FA group comparison within ROIs of the ICBM-DTI-81 WM atlas. Only regions with Qvoxels ≥ 0.10 are shown.

**Table S3.3.2:** FA FreeSurfer GM atlas

| ROI | t_peak | g_peak | Qvoxels |
|---|---|---|---|
| Left-Cerebellum-Cortex | -15.155 | -5.471 | 0.829 |
| Left-Thalamus-Proper | -6.382 | -2.560 | 0.276 |

| | | | |
|---|---|---|---|
| Left-Caudate | -4.602 | -1.833 | 0.136 |
| Left-Putamen | -4.722 | -1.881 | 0.127 |
| Left-Pallidum | -5.069 | -1.991 | 0.184 |
| Brain-Stem | -16.335 | -5.591 | 0.639 |
| Left-Hippocampus | -7.866 | -3.114 | 0.350 |
| Left-Amygdala | -6.477 | -2.105 | 0.118 |
| Left-VentralDC | -8.732 | -3.276 | 0.524 |
| Right-Cerebellum-Cortex | -15.866 | -5.266 | 0.824 |
| Right-Thalamus-Proper | -6.969 | -2.816 | 0.261 |
| Right-Pallidum | -6.381 | -2.464 | 0.180 |
| Right-Hippocampus | -5.964 | -2.356 | 0.287 |
| Right-VentralDC | -7.614 | -2.925 | 0.435 |

Table S3.3.2: Peak *t*-statistic value, peak Hedge's *g* value, and the fraction of ROI voxels significantly different between HVs and SCA7 patients (Qvoxels) for the FA group comparison within ROIs of the FreeSurfer GM atlas. Only regions with Qvoxels ≥ 0.10 are shown.

#### S4: Correlation analysis supplementary material

##### Figure S4.1: Correlations between imaging metrics and SARA score

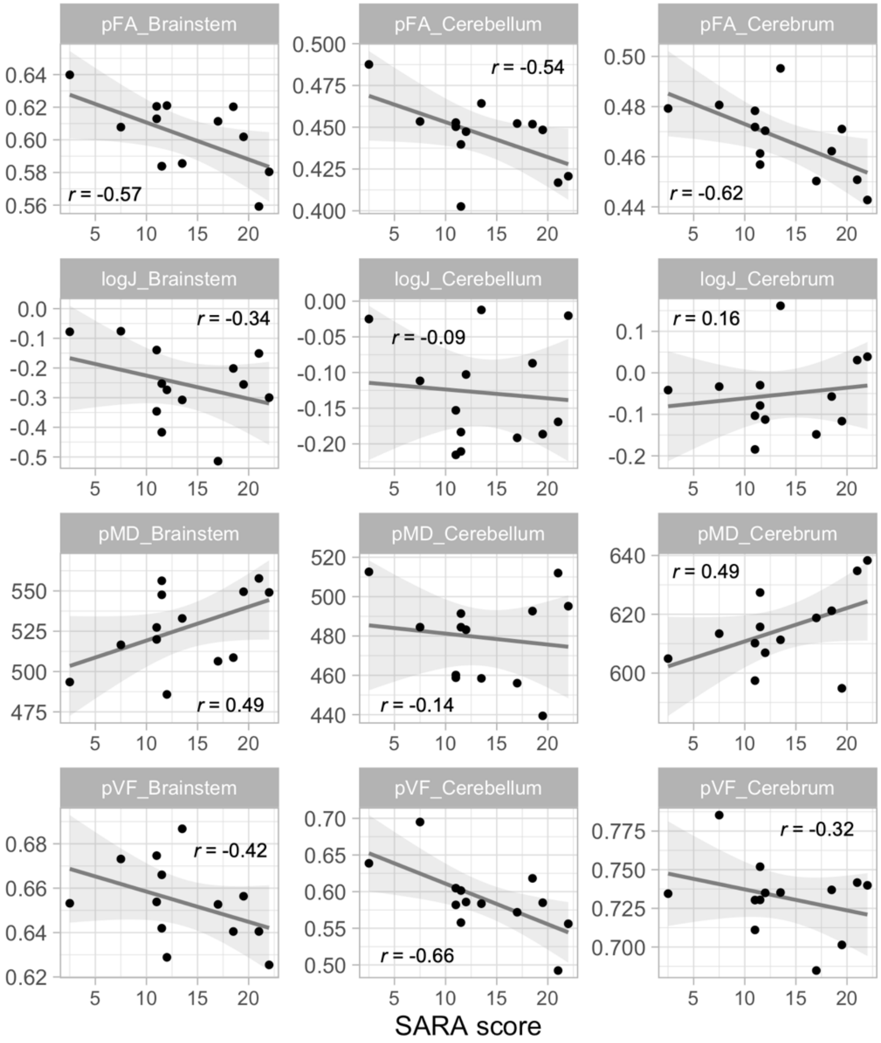


Figure S4.1: Scatter plots of the average value of every metric versus the SARA score in the brainstem, cerebellum, and cerebrum. *r* values shown are Pearson's correlation coefficient. Line and shaded area represent line of best fit and 95% confidence interval respectively.

**Figure S4.2: Correlations of single compartment diffusivity metrics (MD and FA) and dual compartment diffusivity metrics (pMD and pFA) with the parenchymal VF**

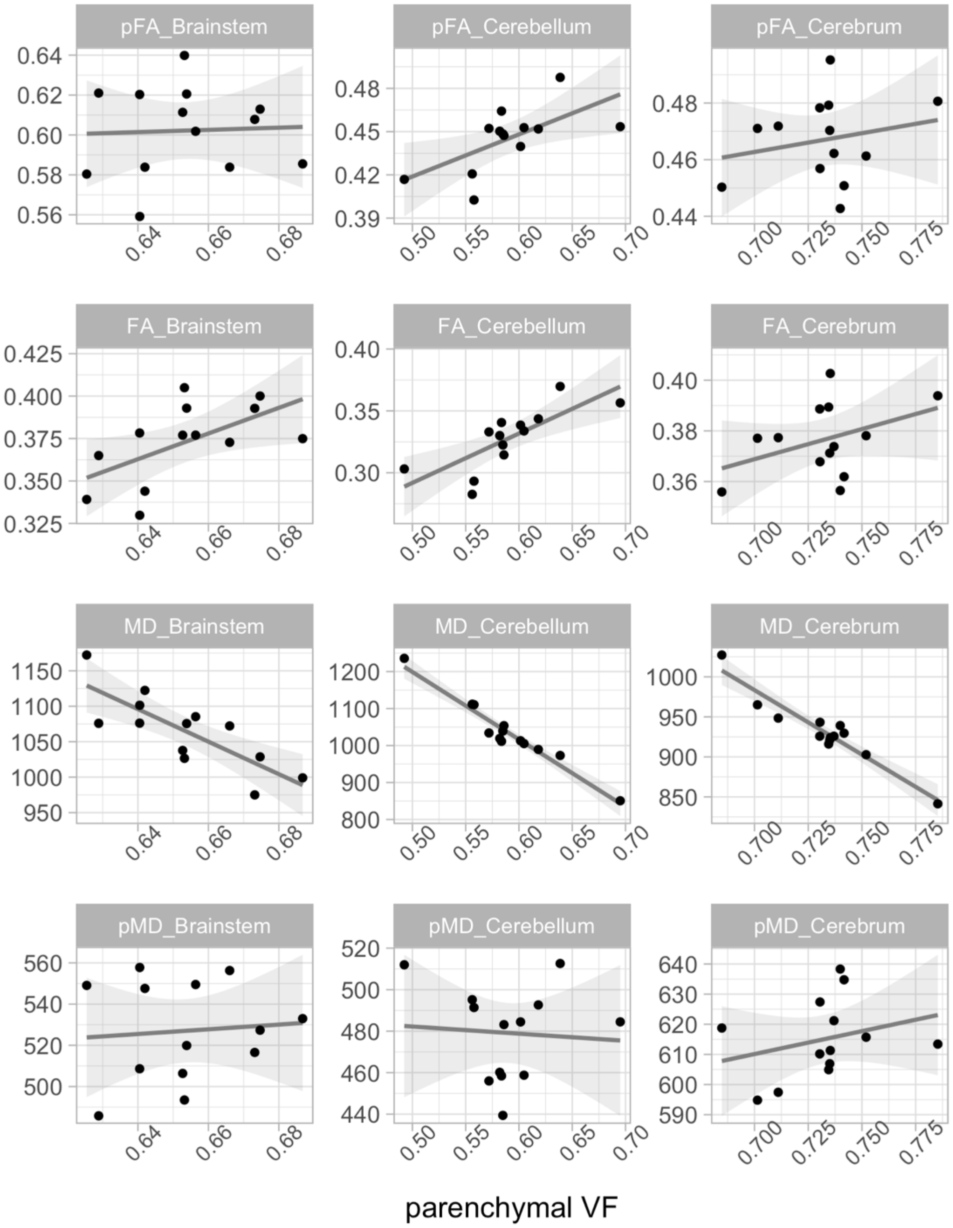


Figure S4.2: Scatter plots of the average value of single and dual compartment diffusivity metrics versus the parenchymal VF in the brainstem, cerebellum, and cerebrum. As labeled, pMD = parenchymal (dual compartment) MD, pFA = parenchymal (dual compartment) FA, MD = single compartment MD, and FA = single compartment FA. Line and shaded area represent line of best fit and 95% confidence interval respectively. Compared to the dual compartment diffusivity metrics, the single compartment diffusivity metrics are more strongly correlated with the parenchymal VF. This demonstrates that abnormalities in the single compartment MD and FA may strongly reflect differences in parenchymal tissue volume rather than purely tissue microstructural abnormalities. Given that we observe significant changes in parenchymal VF around in brainstem and cerebellum in patients, we believe the dual-compartment diffusivity metrics are more appropriate for assessing tissue microstructure.

**Figure S4.3: Correlations of whole brain pFA and cerebellar pVF with the SARA score with asymptomatic patient included**

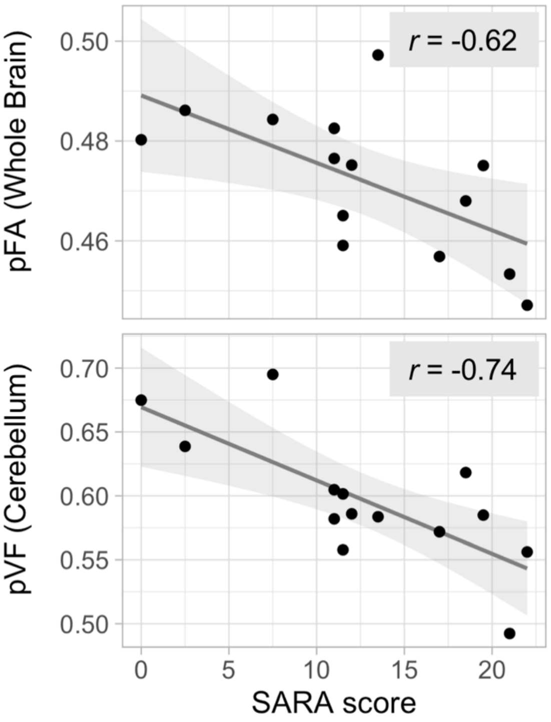


Figure S4.3: Scatter plots of whole brain pFA versus the SARA score ($r$ = -0.62, $P$ = 0.018, uncorrected) and cerebellar pVF versus the SARA score ($r$ = -0.74, $P$ = 0.002, uncorrected). Line and shaded area represent line of best fit and 95% confidence interval respectively.

**Figure S4.4: Correlations between imaging metrics and SARA score with asymptomatic patient included**

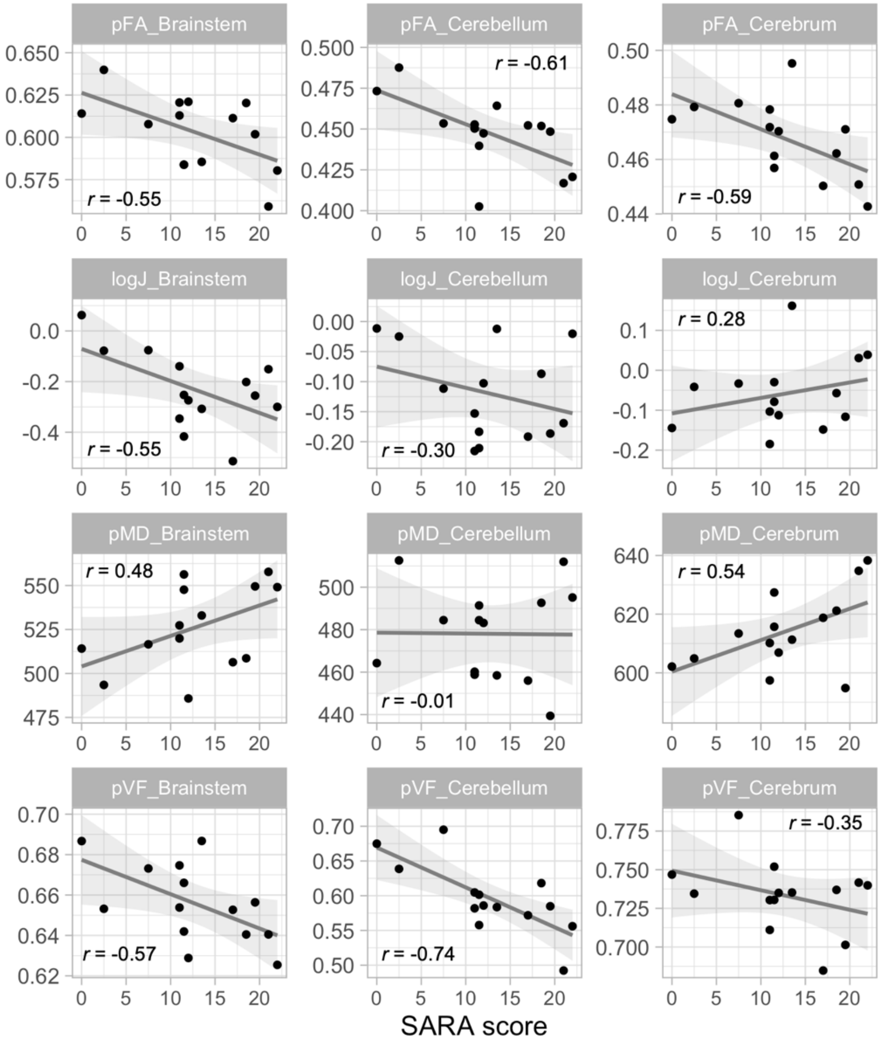


Figure S4.4: Scatter plots of the average value of every metric versus the SARA score in the brainstem, cerebellum, and cerebrum. *r* values shown are Pearson's correlation coefficient. Line and shaded area represent line of best fit and 95% confidence interval respectively.
