## Supplementary Material 2 of 2 for "*In vivo* assessment of neurodegeneration in Spinocerebellar Ataxia type 7"

#### Supplementary Materials 2 of 2

### Contents


**S5: Group comparison all metrics full atlas tables**
- **Table S5.1:** All metrics full ICBM-DTI-81 WM atlas
- **Table S5.2:** All metrics full FreeSurfer GM atlas

**S6: Group comparison full atlas tables**
- **S6.1:** DTBM (logJ) full tables
  - **Table S6.1.1:** DTBM (logJ) full ICBM-DTI-81 WM atlas
  - **Table S6.1.2:** DTBM (logJ) full FreeSurfer GM atlas
- **S6.2:** pVF full tables
  - **Table S6.2.1:** pVF full ICBM-DTI-81 WM atlas
  - **Table S6.2.2:** pVF full FreeSurfer GM atlas
- **S6.3:** pMD full tables
  - **Table S6.3.1:** pMD full ICBM-DTI-81 WM atlas
  - **Table S6.3.2:** pMD full FreeSurfer GM atlas
- **S6.4:** pFA full tables
  - **Table S6.4.1:** pFA full ICBM-DTI-81 WM atlas
  - **Table S6.4.2:** pFA full FreeSurfer GM atlas
- **S6.5:** VBM full table
  - **Table S6.5.1:** VBM full FreeSurfer GM atlas
- **S6.6:** MD full tables
  - **Table S6.6.1:** MD full ICBM-DTI-81 WM atlas
  - **Table S6.6.2:** MD full FreeSurfer GM atlas
- **S6.7:** FA full tables
  - **Table S6.7.1:** FA full ICBM-DTI-81 WM atlas
  - **Table S6.7.2:** FA full FreeSurfer GM atlas

### S5: Group comparison all metrics full atlas tables

**Table S5.1:** All metrics full ICBM-DTI-81 WM atlas table

| ROI | DTBM | pVF | pMD | MD | pFA | FA |
|---|---|---|---|---|---|---|
| Middle cerebellar peduncle | 0.826 | 0.304 | 0.608 | 0.922 | 0.550 | 0.740 |
| Pontine crossing tract (a part of MCP) | 0.997 | 0.000 | 0.812 | 0.960 | 0.806 | 0.910 |
| Genu of corpus callosum | 0.000 | 0.000 | 0.123 | 0.074 | 0.005 | 0.058 |
| Body of corpus callosum | 0.040 | 0.008 | 0.192 | 0.196 | 0.124 | 0.145 |
| Splenium of corpus callosum | 0.019 | 0.010 | 0.137 | 0.079 | 0.203 | 0.163 |
| Fornix (column and body of fornix) | 0.004 | 0.000 | 0.000 | 0.023 | 0.000 | 0.010 |
| Corticospinal tract R | 0.884 | 0.063 | 0.619 | 0.786 | 0.579 | 0.675 |
| Corticospinal tract L | 0.911 | 0.035 | 0.641 | 0.812 | 0.510 | 0.626 |
| Medial lemniscus R | 0.974 | 0.300 | 0.423 | 0.773 | 0.525 | 0.884 |
| Medial lemniscus L | 0.895 | 0.277 | 0.328 | 0.781 | 0.438 | 0.819 |
| Inferior cerebellar peduncle R | 0.843 | 0.573 | 0.297 | 0.903 | 0.311 | 0.671 |
| Inferior cerebellar peduncle L | 0.822 | 0.439 | 0.313 | 0.813 | 0.203 | 0.515 |
| Superior cerebellar peduncle R | 0.548 | 0.705 | 0.162 | 0.896 | 0.143 | 0.929 |
| Superior cerebellar peduncle L | 0.514 | 0.635 | 0.222 | 0.838 | 0.242 | 0.892 |
| Cerebral peduncle R | 0.606 | 0.141 | 0.321 | 0.538 | 0.349 | 0.456 |
| Cerebral peduncle L | 0.679 | 0.175 | 0.312 | 0.538 | 0.413 | 0.603 |
| Anterior limb of internal capsule R | 0.051 | 0.000 | 0.044 | 0.010 | 0.027 | 0.004 |
| Anterior limb of internal capsule L | 0.296 | 0.000 | 0.151 | 0.342 | 0.384 | 0.443 |
| Posterior limb of internal capsule R | 0.449 | 0.000 | 0.318 | 0.647 | 0.266 | 0.268 |
| Posterior limb of internal capsule L | 0.387 | 0.000 | 0.577 | 0.772 | 0.315 | 0.297 |
| Retrolenticular part of internal capsule R | 0.189 | 0.000 | 0.290 | 0.369 | 0.326 | 0.411 |
| Retrolenticular part of internal capsule L | 0.112 | 0.000 | 0.363 | 0.330 | 0.267 | 0.196 |
| Anterior corona radiata R | 0.049 | 0.000 | 0.135 | 0.025 | 0.021 | 0.004 |
| Anterior corona radiata L | 0.062 | 0.000 | 0.181 | 0.110 | 0.121 | 0.134 |
| Superior corona radiata R | 0.504 | 0.000 | 0.505 | 0.356 | 0.279 | 0.103 |
| Superior corona radiata L | 0.426 | 0.000 | 0.550 | 0.735 | 0.346 | 0.282 |
| Posterior corona radiata R | 0.164 | 0.000 | 0.338 | 0.280 | 0.064 | 0.037 |
| Posterior corona radiata L | 0.162 | 0.000 | 0.468 | 0.424 | 0.204 | 0.089 |
| Posterior thalamic radiation R | 0.000 | 0.000 | 0.658 | 0.211 | 0.537 | 0.432 |
| Posterior thalamic radiation L | 0.000 | 0.000 | 0.515 | 0.290 | 0.483 | 0.378 |
| Sagittal stratum R | 0.000 | 0.000 | 0.292 | 0.239 | 0.248 | 0.193 |
| Sagittal stratum L | 0.006 | 0.000 | 0.430 | 0.449 | 0.329 | 0.276 |
| External capsule R | 0.035 | 0.000 | 0.031 | 0.004 | 0.020 | 0.014 |
| External capsule L | 0.010 | 0.000 | 0.131 | 0.181 | 0.129 | 0.087 |
| Cingulum (cingulate gyrus) R | 0.000 | 0.000 | 0.048 | 0.148 | 0.023 | 0.007 |
| Cingulum (cingulate gyrus) L | 0.000 | 0.007 | 0.060 | 0.299 | 0.106 | 0.097 |
| Cingulum (hippocampus) R | 0.000 | 0.021 | 0.005 | 0.199 | 0.013 | 0.030 |
| Cingulum (hippocampus) L | 0.000 | 0.019 | 0.033 | 0.042 | 0.000 | 0.000 |
| Fornix (cres) / Stria terminalis R | 0.446 | 0.131 | 0.300 | 0.696 | 0.212 | 0.553 |
| Fornix (cres) / Stria terminalis L | 0.548 | 0.095 | 0.366 | 0.669 | 0.359 | 0.574 |
| Superior longitudinal fasciculus R | 0.000 | 0.000 | 0.306 | 0.115 | 0.161 | 0.010 |
| Superior longitudinal fasciculus L | 0.026 | 0.000 | 0.302 | 0.331 | 0.091 | 0.038 |
| Superior fronto-occipital fasciculus R | 0.240 | 0.000 | 0.163 | 0.110 | 0.107 | 0.000 |
| Superior fronto-occipital fasciculus L | 0.324 | 0.000 | 0.161 | 0.268 | 0.085 | 0.044 |
| Uncinate fasciculus R | 0.000 | 0.000 | 0.000 | 0.000 | 0.000 | 0.000 |
| Uncinate fasciculus L | 0.000 | 0.000 | 0.112 | 0.378 | 0.000 | 0.000 |
| Tapetum R | 0.157 | 0.000 | 0.236 | 0.090 | 0.249 | 0.288 |
| Tapetum L | 0.082 | 0.001 | 0.057 | 0.020 | 0.182 | 0.282 |

Legend:

0.5 ≤ Qvoxels

0.1 < Qvoxels ≤ 0.5

0 < Qvoxels < 0.1

Table S5.1: The fraction of ROI voxels significantly different between HVs and SCA7 patients (Qvoxels) for the group comparison of each imaging metric within each ROI of the ICBM-DTI-81 WM atlas.

**Table S5.2:** All metrics FreeSurfer GM atlas table

| ROI | DTBM | VBM | pVF | pMD | MD | pFA | FA |
|---|---|---|---|---|---|---|---|
| Left-Cerebellum-Cortex | 0.411 | 0.282 | 0.611 | 0.098 | 0.807 | 0.548 | 0.829 |
| Left-Thalamus-Proper | 0.550 | 0.199 | 0.016 | 0.256 | 0.454 | 0.168 | 0.276 |
| Left-Caudate | 0.007 | 0.018 | 0.000 | 0.026 | 0.066 | 0.064 | 0.136 |
| Left-Putamen | 0.011 | 0.015 | 0.000 | 0.075 | 0.127 | 0.163 | 0.127 |
| Left-Pallidum | 0.035 | 0.030 | 0.000 | 0.185 | 0.103 | 0.180 | 0.184 |
| Brain-Stem | 0.731 | 0.794 | 0.219 | 0.408 | 0.742 | 0.370 | 0.639 |
| Left-Hippocampus | 0.101 | 0.007 | 0.142 | 0.065 | 0.192 | 0.009 | 0.350 |
| Left-Amygdala | 0.018 | 0.000 | 0.034 | 0.112 | 0.115 | 0.026 | 0.118 |
| Left-Accumbens-area | 0.000 | 0.000 | 0.000 | 0.000 | 0.000 | 0.000 | 0.000 |
| Left-VentralDC | 0.554 | 0.177 | 0.154 | 0.192 | 0.442 | 0.302 | 0.524 |
| Right-Cerebellum-Cortex | 0.412 | 0.243 | 0.588 | 0.151 | 0.815 | 0.608 | 0.824 |
| Right-Thalamus-Proper | 0.565 | 0.259 | 0.025 | 0.215 | 0.463 | 0.176 | 0.261 |
| Right-Caudate | 0.033 | 0.079 | 0.000 | 0.007 | 0.027 | 0.002 | 0.002 |
| Right-Putamen | 0.014 | 0.078 | 0.000 | 0.016 | 0.014 | 0.030 | 0.029 |
| Right-Pallidum | 0.079 | 0.080 | 0.000 | 0.177 | 0.123 | 0.183 | 0.180 |
| Right-Hippocampus | 0.050 | 0.020 | 0.101 | 0.006 | 0.154 | 0.000 | 0.287 |
| Right-Amygdala | 0.000 | 0.000 | 0.019 | 0.002 | 0.004 | 0.000 | 0.051 |
| Right-Accumbens-area | 0.000 | 0.000 | 0.000 | 0.000 | 0.000 | 0.000 | 0.000 |
| Right-VentralDC | 0.445 | 0.234 | 0.145 | 0.162 | 0.417 | 0.186 | 0.435 |
| ctx-lh-bankssts | 0.000 | 0.000 | 0.000 | 0.000 | 0.002 | NA | NA |
| ctx-lh-caudalanteriorcingulate | 0.000 | 0.000 | 0.000 | 0.040 | 0.262 | NA | NA |
| ctx-lh-caudalmiddlefrontal | 0.000 | 0.000 | 0.000 | 0.113 | 0.065 | NA | NA |
| ctx-lh-cuneus | 0.078 | 0.000 | 0.030 | 0.102 | 0.161 | NA | NA |
| ctx-lh-entorhinal | 0.013 | 0.000 | 0.072 | 0.032 | 0.210 | NA | NA |
| ctx-lh-fusiform | 0.000 | 0.054 | 0.031 | 0.050 | 0.129 | NA | NA |
| ctx-lh-inferiorparietal | 0.000 | 0.000 | 0.000 | 0.089 | 0.026 | NA | NA |
| ctx-lh-inferiortemporal | 0.000 | 0.000 | 0.000 | 0.034 | 0.051 | NA | NA |
| ctx-lh-isthmuscingulate | 0.000 | 0.005 | 0.029 | 0.013 | 0.134 | NA | NA |
| ctx-lh-lateraloccipital | 0.001 | 0.000 | 0.015 | 0.094 | 0.139 | NA | NA |
| ctx-lh-lateralorbitofrontal | 0.000 | 0.000 | 0.000 | 0.062 | 0.004 | NA | NA |
| ctx-lh-lingual | 0.108 | 0.089 | 0.003 | 0.091 | 0.159 | NA | NA |
| ctx-lh-medialorbitofrontal | 0.001 | 0.000 | 0.000 | 0.017 | 0.001 | NA | NA |
| ctx-lh-middletemporal | 0.000 | 0.000 | 0.000 | 0.068 | 0.006 | NA | NA |
| ctx-lh-parahippocampal | 0.030 | 0.000 | 0.014 | 0.009 | 0.074 | NA | NA |
| ctx-lh-paracentral | 0.000 | 0.000 | 0.000 | 0.073 | 0.117 | NA | NA |
| ctx-lh-parsopercularis | 0.000 | 0.000 | 0.000 | 0.066 | 0.080 | NA | NA |

| | | | | | | | |
|---|---|---|---|---|---|---|---|
| ctx-lh-parsorbitalis | 0.000 | 0.000 | 0.000 | 0.032 | 0.000 | NA | NA |
| ctx-lh-parstriangularis | 0.000 | 0.000 | 0.000 | 0.003 | 0.003 | NA | NA |
| ctx-lh-pericalcarine | 0.028 | 0.000 | 0.020 | 0.029 | 0.358 | NA | NA |
| ctx-lh-postcentral | 0.000 | 0.001 | 0.000 | 0.136 | 0.119 | NA | NA |
| ctx-lh-posteriorcingulate | 0.002 | 0.000 | 0.020 | 0.028 | 0.194 | NA | NA |
| ctx-lh-precentral | 0.000 | 0.021 | 0.000 | 0.139 | 0.089 | NA | NA |
| ctx-lh-precuneus | 0.051 | 0.000 | 0.003 | 0.109 | 0.123 | NA | NA |
| ctx-lh-rostralanteriorcingulate | 0.000 | 0.000 | 0.000 | 0.031 | 0.051 | NA | NA |
| ctx-lh-rostralmiddlefrontal | 0.000 | 0.000 | 0.000 | 0.104 | 0.039 | NA | NA |
| ctx-lh-superiorfrontal | 0.000 | 0.000 | 0.000 | 0.160 | 0.063 | NA | NA |
| ctx-lh-superiorparietal | 0.000 | 0.001 | 0.000 | 0.131 | 0.075 | NA | NA |
| ctx-lh-superiortemporal | 0.000 | 0.000 | 0.000 | 0.035 | 0.014 | NA | NA |
| ctx-lh-supramarginal | 0.000 | 0.000 | 0.000 | 0.063 | 0.027 | NA | NA |
| ctx-lh-frontalpole | 0.000 | 0.000 | 0.000 | 0.074 | 0.000 | NA | NA |
| ctx-lh-temporalpole | 0.000 | 0.000 | 0.027 | 0.002 | 0.034 | NA | NA |
| ctx-lh-transversetemporal | 0.000 | 0.000 | 0.000 | 0.039 | 0.011 | NA | NA |
| ctx-lh-insula | 0.002 | 0.000 | 0.000 | 0.030 | 0.147 | NA | NA |
| ctx-rh-bankssts | 0.000 | 0.000 | 0.000 | 0.026 | 0.016 | NA | NA |
| ctx-rh-caudalanteriorcingulate | 0.000 | 0.000 | 0.000 | 0.035 | 0.182 | NA | NA |
| ctx-rh-caudalmiddlefrontal | 0.000 | 0.000 | 0.000 | 0.085 | 0.014 | NA | NA |
| ctx-rh-cuneus | 0.025 | 0.000 | 0.011 | 0.078 | 0.197 | NA | NA |
| ctx-rh-entorhinal | 0.007 | 0.003 | 0.071 | 0.000 | 0.065 | NA | NA |
| ctx-rh-fusiform | 0.013 | 0.069 | 0.061 | 0.086 | 0.163 | NA | NA |
| ctx-rh-inferiorparietal | 0.000 | 0.000 | 0.000 | 0.092 | 0.083 | NA | NA |
| ctx-rh-inferiortemporal | 0.000 | 0.000 | 0.000 | 0.017 | 0.024 | NA | NA |
| ctx-rh-isthmuscingulate | 0.000 | 0.000 | 0.073 | 0.018 | 0.226 | NA | NA |
| ctx-rh-lateraloccipital | 0.000 | 0.000 | 0.021 | 0.124 | 0.233 | NA | NA |
| ctx-rh-lateralorbitofrontal | 0.001 | 0.000 | 0.000 | 0.000 | 0.000 | NA | NA |
| ctx-rh-lingual | 0.093 | 0.036 | 0.016 | 0.103 | 0.137 | NA | NA |
| ctx-rh-medialorbitofrontal | 0.000 | 0.000 | 0.000 | 0.000 | 0.000 | NA | NA |
| ctx-rh-middletemporal | 0.000 | 0.000 | 0.000 | 0.004 | 0.037 | NA | NA |
| ctx-rh-parahippocampal | 0.030 | 0.000 | 0.049 | 0.001 | 0.185 | NA | NA |
| ctx-rh-paracentral | 0.000 | 0.000 | 0.000 | 0.158 | 0.217 | NA | NA |
| ctx-rh-parsopercularis | 0.000 | 0.000 | 0.000 | 0.029 | 0.002 | NA | NA |
| ctx-rh-parsorbitalis | 0.000 | 0.000 | 0.000 | 0.000 | 0.000 | NA | NA |
| ctx-rh-parstriangularis | 0.000 | 0.000 | 0.000 | 0.095 | 0.001 | NA | NA |
| ctx-rh-pericalcarine | 0.001 | 0.000 | 0.022 | 0.083 | 0.347 | NA | NA |
| ctx-rh-postcentral | 0.000 | 0.053 | 0.000 | 0.047 | 0.064 | NA | NA |
| ctx-rh-posteriorcingulate | 0.027 | 0.004 | 0.000 | 0.028 | 0.228 | NA | NA |
| ctx-rh-precentral | 0.000 | 0.107 | 0.000 | 0.068 | 0.062 | NA | NA |
| ctx-rh-precuneus | 0.056 | 0.000 | 0.000 | 0.097 | 0.087 | NA | NA |
| ctx-rh-rostralanteriorcingulate | 0.000 | 0.000 | 0.000 | 0.008 | 0.005 | NA | NA |
| ctx-rh-rostralmiddlefrontal | 0.000 | 0.000 | 0.000 | 0.047 | 0.000 | NA | NA |
| ctx-rh-superiorfrontal | 0.000 | 0.000 | 0.000 | 0.056 | 0.057 | NA | NA |
| ctx-rh-superiorparietal | 0.007 | 0.020 | 0.000 | 0.109 | 0.079 | NA | NA |
| ctx-rh-superiortemporal | 0.000 | 0.000 | 0.000 | 0.001 | 0.011 | NA | NA |
| ctx-rh-supramarginal | 0.000 | 0.011 | 0.000 | 0.075 | 0.033 | NA | NA |
| ctx-rh-frontalpole | 0.000 | 0.000 | 0.000 | 0.000 | 0.000 | NA | NA |
| ctx-rh-temporalpole | 0.000 | 0.000 | 0.000 | 0.000 | 0.000 | NA | NA |
| ctx-rh-transversetemporal | 0.000 | 0.000 | 0.000 | 0.013 | 0.064 | NA | NA |
| ctx-rh-insula | 0.007 | 0.000 | 0.000 | 0.001 | 0.021 | NA | NA |

Legend:

0.5 ≤ Qvoxels

0.1 < Qvoxels ≤ 0.5

0 < Qvoxels < 0.1

Table S5.2: The fraction of ROI voxels significantly different between HVs and SCA7 patients (Qvoxels) for the group comparison of each imaging metric within each ROI of the FreeSurfer GM atlas.

#### S6: Group comparison full atlas tables

##### **S6.1:** DTBM (logJ) full tables

**Table S6.1.1:** DTBM (logJ) full ICBM-DTI-81 WM atlas table

| ROI | *t* peak | *g* peak | Qvoxels |
|---|---|---|---|
| Middle cerebellar peduncle | -10.534 | -3.882 | 0.826 |
| Pontine crossing tract (a part of MCP) | -10.455 | -3.583 | 0.997 |
| Genu of corpus callosum | -2.824 | -0.604 | 0.000 |
| Body of corpus callosum | -5.726 | -2.068 | 0.040 |
| Splenium of corpus callosum | -3.813 | -1.304 | 0.019 |
| Fornix (column and body of fornix) | -2.584 | -1.044 | 0.004 |
| Corticospinal tract R | -9.272 | -3.090 | 0.884 |
| Corticospinal tract L | -9.103 | -3.061 | 0.911 |
| Medial lemniscus R | -13.412 | -4.443 | 0.974 |
| Medial lemniscus L | -11.353 | -4.439 | 0.895 |
| Inferior cerebellar peduncle R | -13.367 | -5.412 | 0.843 |
| Inferior cerebellar peduncle L | -12.846 | -5.309 | 0.822 |
| Superior cerebellar peduncle R | -13.362 | -4.735 | 0.548 |
| Superior cerebellar peduncle L | -8.964 | -3.532 | 0.514 |
| Cerebral peduncle R | -10.466 | -2.840 | 0.606 |
| Cerebral peduncle L | -11.885 | -2.682 | 0.679 |
| Anterior limb of internal capsule R | -4.255 | -1.823 | 0.051 |
| Anterior limb of internal capsule L | -6.626 | -2.382 | 0.296 |
| Posterior limb of internal capsule R | -9.659 | -2.100 | 0.449 |
| Posterior limb of internal capsule L | -9.206 | -2.157 | 0.387 |
| Retrolenticular part of internal capsule R | -5.109 | -1.431 | 0.189 |
| Retrolenticular part of internal capsule L | -4.132 | -1.205 | 0.112 |
| Anterior corona radiata R | -5.114 | -1.628 | 0.049 |
| Anterior corona radiata L | -3.940 | -1.482 | 0.062 |
| Superior corona radiata R | -5.668 | -2.420 | 0.504 |
| Superior corona radiata L | -6.781 | -2.658 | 0.426 |
| Posterior corona radiata R | -4.286 | -1.655 | 0.164 |
| Posterior corona radiata L | -4.578 | -1.990 | 0.162 |
| Posterior thalamic radiation R | 0.000 | 0.000 | 0.000 |
| Posterior thalamic radiation L | 0.000 | 0.000 | 0.000 |

| | | | |
|---|---|---|---|
| Sagittal stratum R | 0.000 | 0.000 | 0.000 |
| Sagittal stratum L | -3.046 | -1.330 | 0.006 |
| External capsule R | -5.388 | -1.922 | 0.035 |
| External capsule L | -3.297 | -1.277 | 0.010 |
| Cingulum (cingulate gyrus) R | 0.000 | 0.000 | 0.000 |
| Cingulum (cingulate gyrus) L | 0.000 | 0.000 | 0.000 |
| Cingulum (hippocampus) R | 0.000 | 0.000 | 0.000 |
| Cingulum (hippocampus) L | 0.000 | 0.000 | 0.000 |
| Fornix (cres) / Stria terminalis R | -5.680 | -2.361 | 0.446 |
| Fornix (cres) / Stria terminalis L | -7.330 | -2.288 | 0.548 |
| Superior longitudinal fasciculus R | 0.000 | 0.000 | 0.000 |
| Superior longitudinal fasciculus L | -3.339 | -1.143 | 0.026 |
| Superior fronto-occipital fasciculus R | -4.605 | -1.991 | 0.240 |
| Superior fronto-occipital fasciculus L | -5.707 | -2.039 | 0.324 |
| Uncinate fasciculus R | 0.000 | 0.000 | 0.000 |
| Uncinate fasciculus L | 0.000 | 0.000 | 0.000 |
| Tapetum R | -3.356 | -1.500 | 0.157 |
| Tapetum L | -3.609 | -1.605 | 0.082 |

Legend:

0.5 ≤ Qvoxels

0.1 < Qvoxels ≤ 0.5

0 < Qvoxels < 0.1

Table S6.1.1: Peak *t* statistic value, peak Hedge's *g* value, and the fraction of ROI voxels significantly different between HVs and SCA7 patients (Qvoxels) for the DTBM (logJ) group comparison within each ROI of the ICBM-DTI-81 WM atlas.

**Table S6.1.2:** DTBM (logJ) full FreeSurfer GM atlas table

| ROI | t_peak | g_peak | Qvoxels |
|---|---|---|---|
| Left-Cerebellum-Cortex | -9.428 | -3.628 | 0.411 |
| Left-Thalamus-Proper | -8.969 | -2.358 | 0.550 |
| Left-Caudate | -4.296 | -1.585 | 0.007 |
| Left-Putamen | -5.281 | -1.939 | 0.011 |
| Left-Pallidum | -5.877 | -1.660 | 0.035 |
| Brain-Stem | -13.412 | -5.412 | 0.731 |
| Left-Hippocampus | -5.870 | -1.858 | 0.101 |
| Left-Amygdala | -3.917 | -1.318 | 0.018 |
| Left-Accumbens-area | 0.000 | 0.000 | 0.000 |
| Left-VentralDC | -11.885 | -2.834 | 0.554 |
| Right-Cerebellum-Cortex | -10.807 | -4.785 | 0.412 |
| Right-Thalamus-Proper | -8.155 | -2.661 | 0.565 |
| Right-Caudate | -5.393 | -1.841 | 0.033 |
| Right-Putamen | -3.348 | -1.065 | 0.014 |
| Right-Pallidum | -8.617 | -2.100 | 0.079 |
| Right-Hippocampus | -7.364 | -1.959 | 0.050 |
| Right-Amygdala | 0.000 | 0.000 | 0.000 |
| Right-Accumbens-area | 0.000 | 0.000 | 0.000 |

| | | | |
|---|---|---|---|
| Right-VentralDC | -8.824 | -2.901 | 0.445 |
| ctx-lh-bankssts | 0.000 | 0.000 | 0.000 |
| ctx-lh-caudalanteriorcingulate | 0.000 | 0.000 | 0.000 |
| ctx-lh-caudalmiddlefrontal | -2.632 | -0.990 | 0.000 |
| ctx-lh-cuneus | 5.493 | 1.398 | 0.078 |
| ctx-lh-entorhinal | -4.968 | -1.884 | 0.013 |
| ctx-lh-fusiform | 6.459 | 1.139 | 0.000 |
| ctx-lh-inferiorparietal | 0.000 | 0.000 | 0.000 |
| ctx-lh-inferiortemporal | 0.000 | 0.000 | 0.000 |
| ctx-lh-isthmuscingulate | 0.000 | 0.000 | 0.000 |
| ctx-lh-lateraloccipital | -3.025 | -1.177 | 0.001 |
| ctx-lh-lateralorbitofrontal | 0.000 | 0.000 | 0.000 |
| ctx-lh-lingual | 6.286 | 1.575 | 0.108 |
| ctx-lh-medialorbitofrontal | -3.175 | -1.394 | 0.001 |
| ctx-lh-middletemporal | 0.000 | 0.000 | 0.000 |
| ctx-lh-parahippocampal | 5.352 | 2.022 | 0.030 |
| ctx-lh-paracentral | 0.000 | 0.000 | 0.000 |
| ctx-lh-parsopercularis | 0.000 | 0.000 | 0.000 |
| ctx-lh-parsorbitalis | 0.000 | 0.000 | 0.000 |
| ctx-lh-parstriangularis | 0.000 | 0.000 | 0.000 |
| ctx-lh-pericalcarine | 4.329 | 0.707 | 0.028 |
| ctx-lh-postcentral | 0.000 | 0.000 | 0.000 |
| ctx-lh-posteriorcingulate | -3.874 | -1.271 | 0.002 |
| ctx-lh-precentral | -3.876 | -1.414 | 0.000 |
| ctx-lh-precuneus | 5.687 | 1.254 | 0.051 |
| ctx-lh-rostralanteriorcingulate | 0.000 | 0.000 | 0.000 |
| ctx-lh-rostralmiddlefrontal | 0.000 | 0.000 | 0.000 |
| ctx-lh-superiorfrontal | -3.400 | -1.255 | 0.000 |
| ctx-lh-superiorparietal | 0.000 | 0.000 | 0.000 |
| ctx-lh-superiortemporal | 0.000 | 0.000 | 0.000 |
| ctx-lh-supramarginal | 0.000 | 0.000 | 0.000 |
| ctx-lh-frontalpole | 0.000 | 0.000 | 0.000 |
| ctx-lh-temporalpole | 0.000 | 0.000 | 0.000 |
| ctx-lh-transversetemporal | 0.000 | 0.000 | 0.000 |
| ctx-lh-insula | -3.390 | -0.960 | 0.002 |
| ctx-rh-bankssts | 0.000 | 0.000 | 0.000 |
| ctx-rh-caudalanteriorcingulate | 0.000 | 0.000 | 0.000 |
| ctx-rh-caudalmiddlefrontal | 0.000 | 0.000 | 0.000 |
| ctx-rh-cuneus | 5.573 | 1.305 | 0.025 |
| ctx-rh-entorhinal | 5.704 | 1.817 | 0.007 |
| ctx-rh-fusiform | 6.081 | 1.582 | 0.013 |
| ctx-rh-inferiorparietal | 0.000 | 0.000 | 0.000 |
| ctx-rh-inferiortemporal | 0.000 | 0.000 | 0.000 |
| ctx-rh-isthmuscingulate | 0.000 | 0.000 | 0.000 |
| ctx-rh-lateraloccipital | 0.000 | 0.000 | 0.000 |
| ctx-rh-lateralorbitofrontal | -3.364 | -0.907 | 0.001 |
| ctx-rh-lingual | 6.531 | 1.630 | 0.093 |
| ctx-rh-medialorbitofrontal | 0.000 | 0.000 | 0.000 |
| ctx-rh-middletemporal | 0.000 | 0.000 | 0.000 |
| ctx-rh-parahippocampal | 4.978 | 2.126 | 0.030 |
| ctx-rh-paracentral | 0.000 | 0.000 | 0.000 |
| ctx-rh-parsopercularis | 0.000 | 0.000 | 0.000 |
| ctx-rh-parsorbitalis | 0.000 | 0.000 | 0.000 |
| ctx-rh-parstriangularis | 0.000 | 0.000 | 0.000 |
| ctx-rh-pericalcarine | 3.449 | 0.552 | 0.001 |
| ctx-rh-postcentral | 0.000 | 0.000 | 0.000 |

| | | | |
|---|---|---|---|
| ctx-rh-posteriorcingulate | -3.495 | -1.550 | 0.027 |
| ctx-rh-precentral | 0.000 | 0.000 | 0.000 |
| ctx-rh-precuneus | 6.719 | 1.354 | 0.056 |
| ctx-rh-rostralanteriorcingulate | 0.000 | 0.000 | 0.000 |
| ctx-rh-rostralmiddlefrontal | 0.000 | 0.000 | 0.000 |
| ctx-rh-superiorfrontal | 0.000 | 0.000 | 0.000 |
| ctx-rh-superiorparietal | 5.254 | 1.433 | 0.007 |
| ctx-rh-superiortemporal | 0.000 | 0.000 | 0.000 |
| ctx-rh-supramarginal | 0.000 | 0.000 | 0.000 |
| ctx-rh-frontalpole | 0.000 | 0.000 | 0.000 |
| ctx-rh-temporalpole | 0.000 | 0.000 | 0.000 |
| ctx-rh-transversetemporal | 0.000 | 0.000 | 0.000 |
| ctx-rh-insula | -2.975 | -0.500 | 0.007 |

Legend:

0.5 ≤ Qvoxels

0.1 < Qvoxels ≤ 0.5

0 < Qvoxels < 0.1

Table S6.1.2: Peak *t* statistic value, peak Hedge's *g* value, and the fraction of ROI voxels significantly different between HVs and SCA7 patients (Qvoxels) for the DTBM (logJ) group comparison within each ROI of the FreeSurfer GM atlas.

**S6.2:** pVF full tables

**Table S6.2.1:** pVF full ICBM-DTI-81 WM atlas table

| ROI | *t* peak | *g* peak | Qvoxels |
|---|---|---|---|
| Middle cerebellar peduncle | -11.173 | -4.468 | 0.304 |
| Pontine crossing tract (a part of MCP) | 0.000 | 0.000 | 0.000 |
| Genu of corpus callosum | 0.000 | 0.000 | 0.000 |
| Body of corpus callosum | -3.718 | -1.368 | 0.008 |
| Splenium of corpus callosum | -3.618 | -1.278 | 0.010 |
| Fornix (column and body of fornix) | 0.000 | 0.000 | 0.000 |
| Corticospinal tract R | -5.102 | -1.705 | 0.063 |
| Corticospinal tract L | -4.207 | -1.597 | 0.035 |
| Medial lemniscus R | -10.121 | -4.063 | 0.300 |
| Medial lemniscus L | -9.266 | -3.720 | 0.277 |
| Inferior cerebellar peduncle R | -11.150 | -4.358 | 0.573 |
| Inferior cerebellar peduncle L | -9.301 | -3.636 | 0.439 |
| Superior cerebellar peduncle R | -12.941 | -5.176 | 0.705 |
| Superior cerebellar peduncle L | -16.208 | -6.184 | 0.635 |
| Cerebral peduncle R | -7.332 | -2.863 | 0.141 |
| Cerebral peduncle L | -7.189 | -2.451 | 0.175 |
| Anterior limb of internal capsule R | 0.000 | 0.000 | 0.000 |
| Anterior limb of internal capsule L | 0.000 | 0.000 | 0.000 |
| Posterior limb of internal capsule R | 0.000 | 0.000 | 0.000 |
| Posterior limb of internal capsule L | 0.000 | 0.000 | 0.000 |
| Retrolenticular part of internal capsule R | 0.000 | 0.000 | 0.000 |

| | | | |
|---|---|---|---|
| Retrolenticular part of internal capsule L | 0.000 | 0.000 | 0.000 |
| Anterior corona radiata R | 0.000 | 0.000 | 0.000 |
| Anterior corona radiata L | 0.000 | 0.000 | 0.000 |
| Superior corona radiata R | 0.000 | 0.000 | 0.000 |
| Superior corona radiata L | 0.000 | 0.000 | 0.000 |
| Posterior corona radiata R | 0.000 | 0.000 | 0.000 |
| Posterior corona radiata L | 0.000 | 0.000 | 0.000 |
| Posterior thalamic radiation R | 0.000 | 0.000 | 0.000 |
| Posterior thalamic radiation L | 0.000 | 0.000 | 0.000 |
| Sagittal stratum R | 0.000 | 0.000 | 0.000 |
| Sagittal stratum L | 0.000 | 0.000 | 0.000 |
| External capsule R | 0.000 | 0.000 | 0.000 |
| External capsule L | 0.000 | 0.000 | 0.000 |
| Cingulum (cingulate gyrus) R | 0.000 | 0.000 | 0.000 |
| Cingulum (cingulate gyrus) L | -3.973 | -0.998 | 0.007 |
| Cingulum (hippocampus) R | -2.973 | -1.166 | 0.021 |
| Cingulum (hippocampus) L | -3.228 | -1.053 | 0.019 |
| Fornix (cres) / Stria terminalis R | -4.362 | -1.820 | 0.131 |
| Fornix (cres) / Stria terminalis L | -4.017 | -1.431 | 0.095 |
| Superior longitudinal fasciculus R | 0.000 | 0.000 | 0.000 |
| Superior longitudinal fasciculus L | 0.000 | 0.000 | 0.000 |
| Superior fronto-occipital fasciculus R | 0.000 | 0.000 | 0.000 |
| Superior fronto-occipital fasciculus L | 0.000 | 0.000 | 0.000 |
| Uncinate fasciculus R | 0.000 | 0.000 | 0.000 |
| Uncinate fasciculus L | 0.000 | 0.000 | 0.000 |
| Tapetum R | 0.000 | 0.000 | 0.000 |
| Tapetum L | -2.946 | -0.993 | 0.001 |

Legend:

0.5 ≤ Qvoxels

0.1 < Qvoxels ≤ 0.5

0 < Qvoxels < 0.1

Table S6.2.1: Peak *t* statistic value, peak Hedge's *g* value, and the fraction of ROI voxels significantly different between HVs and SCA7 patients (Qvoxels) for the pVF group comparison within each ROI of the ICBM-DTI-81 WM atlas.

**Table S6.2.2:** pVF full FreeSurfer GM atlas table

| ROI | t_peak | g_peak | Qvoxels |
|---|---|---|---|
| Left-Cerebellum-Cortex | -15.178 | -5.707 | 0.611 |
| Left-Thalamus-Proper | -3.581 | -1.310 | 0.016 |
| Left-Caudate | 0.000 | 0.000 | 0.000 |
| Left-Putamen | 0.000 | 0.000 | 0.000 |
| Left-Pallidum | 0.000 | 0.000 | 0.000 |
| Brain-Stem | -16.208 | -6.184 | 0.219 |
| Left-Hippocampus | -4.354 | -1.692 | 0.142 |
| Left-Amygdala | -3.868 | -1.304 | 0.034 |

| | | | |
|---|---|---|---|
| Left-Accumbens-area | 0.000 | 0.000 | 0.000 |
| Left-VentralDC | -7.189 | -2.451 | 0.154 |
| Right-Cerebellum-Cortex | -10.328 | -4.036 | 0.588 |
| Right-Thalamus-Proper | -4.199 | -1.684 | 0.025 |
| Right-Caudate | 0.000 | 0.000 | 0.000 |
| Right-Putamen | 0.000 | 0.000 | 0.000 |
| Right-Pallidum | 0.000 | 0.000 | 0.000 |
| Right-Hippocampus | -4.153 | -1.646 | 0.101 |
| Right-Amygdala | -4.957 | -1.922 | 0.019 |
| Right-Accumbens-area | 0.000 | 0.000 | 0.000 |
| Right-VentralDC | -7.814 | -2.921 | 0.145 |
| ctx-lh-bankssts | 0.000 | 0.000 | 0.000 |
| ctx-lh-caudalanteriorcingulate | 0.000 | 0.000 | 0.000 |
| ctx-lh-caudalmiddlefrontal | 0.000 | 0.000 | 0.000 |
| ctx-lh-cuneus | -5.056 | -1.937 | 0.030 |
| ctx-lh-entorhinal | -4.387 | -1.572 | 0.072 |
| ctx-lh-fusiform | -5.134 | -1.603 | 0.031 |
| ctx-lh-inferiorparietal | 0.000 | 0.000 | 0.000 |
| ctx-lh-inferiortemporal | 0.000 | 0.000 | 0.000 |
| ctx-lh-isthmuscingulate | -3.973 | -1.092 | 0.029 |
| ctx-lh-lateraloccipital | -4.034 | -1.664 | 0.015 |
| ctx-lh-lateralorbitofrontal | 0.000 | 0.000 | 0.000 |
| ctx-lh-lingual | -4.056 | -1.411 | 0.003 |
| ctx-lh-medialorbitofrontal | 0.000 | 0.000 | 0.000 |
| ctx-lh-middletemporal | 0.000 | 0.000 | 0.000 |
| ctx-lh-parahippocampal | -3.321 | -1.144 | 0.014 |
| ctx-lh-paracentral | 0.000 | 0.000 | 0.000 |
| ctx-lh-parsopercularis | 0.000 | 0.000 | 0.000 |
| ctx-lh-parsorbitalis | 0.000 | 0.000 | 0.000 |
| ctx-lh-parstriangularis | 0.000 | 0.000 | 0.000 |
| ctx-lh-pericalcarine | -5.181 | -1.795 | 0.020 |
| ctx-lh-postcentral | 0.000 | 0.000 | 0.000 |
| ctx-lh-posteriorcingulate | -4.414 | -1.246 | 0.020 |
| ctx-lh-precentral | 0.000 | 0.000 | 0.000 |
| ctx-lh-precuneus | -6.004 | -1.719 | 0.003 |
| ctx-lh-rostralanteriorcingulate | 0.000 | 0.000 | 0.000 |
| ctx-lh-rostralmiddlefrontal | 0.000 | 0.000 | 0.000 |
| ctx-lh-superiorfrontal | 0.000 | 0.000 | 0.000 |
| ctx-lh-superiorparietal | 0.000 | 0.000 | 0.000 |
| ctx-lh-superiortemporal | 0.000 | 0.000 | 0.000 |
| ctx-lh-supramarginal | 0.000 | 0.000 | 0.000 |
| ctx-lh-frontalpole | 0.000 | 0.000 | 0.000 |
| ctx-lh-temporalpole | -3.951 | -1.195 | 0.027 |
| ctx-lh-transversetemporal | 0.000 | 0.000 | 0.000 |
| ctx-lh-insula | 0.000 | 0.000 | 0.000 |
| ctx-rh-bankssts | 0.000 | 0.000 | 0.000 |
| ctx-rh-caudalanteriorcingulate | 0.000 | 0.000 | 0.000 |
| ctx-rh-caudalmiddlefrontal | 0.000 | 0.000 | 0.000 |
| ctx-rh-cuneus | -6.507 | -2.545 | 0.011 |
| ctx-rh-entorhinal | -3.628 | -1.475 | 0.071 |
| ctx-rh-fusiform | -7.256 | -2.244 | 0.061 |
| ctx-rh-inferiorparietal | 0.000 | 0.000 | 0.000 |
| ctx-rh-inferiortemporal | 0.000 | 0.000 | 0.000 |
| ctx-rh-isthmuscingulate | -4.086 | -1.402 | 0.073 |
| ctx-rh-lateraloccipital | -5.636 | -2.286 | 0.021 |
| ctx-rh-lateralorbitofrontal | 0.000 | 0.000 | 0.000 |

| | | | |
|---|---|---|---|
| ctx-rh-lingual | -3.929 | -1.462 | 0.016 |
| ctx-rh-medialorbitofrontal | 0.000 | 0.000 | 0.000 |
| ctx-rh-middletemporal | 0.000 | 0.000 | 0.000 |
| ctx-rh-parahippocampal | -4.278 | -1.537 | 0.049 |
| ctx-rh-paracentral | 0.000 | 0.000 | 0.000 |
| ctx-rh-parsopercularis | 0.000 | 0.000 | 0.000 |
| ctx-rh-parsorbitalis | 0.000 | 0.000 | 0.000 |
| ctx-rh-parstriangularis | 0.000 | 0.000 | 0.000 |
| ctx-rh-pericalcarine | -4.660 | -1.797 | 0.022 |
| ctx-rh-postcentral | 0.000 | 0.000 | 0.000 |
| ctx-rh-posteriorcingulate | 0.000 | 0.000 | 0.000 |
| ctx-rh-precentral | 0.000 | 0.000 | 0.000 |
| ctx-rh-precuneus | 0.000 | 0.000 | 0.000 |
| ctx-rh-rostralanteriorcingulate | 0.000 | 0.000 | 0.000 |
| ctx-rh-rostralmiddlefrontal | 0.000 | 0.000 | 0.000 |
| ctx-rh-superiorfrontal | 0.000 | 0.000 | 0.000 |
| ctx-rh-superiorparietal | 0.000 | 0.000 | 0.000 |
| ctx-rh-superiortemporal | 0.000 | 0.000 | 0.000 |
| ctx-rh-supramarginal | 0.000 | 0.000 | 0.000 |
| ctx-rh-frontalpole | 0.000 | 0.000 | 0.000 |
| ctx-rh-temporalpole | 0.000 | 0.000 | 0.000 |
| ctx-rh-transversetemporal | 0.000 | 0.000 | 0.000 |
| ctx-rh-insula | 0.000 | 0.000 | 0.000 |

Legend:

0.5 ≤ Qvoxels

0.1 < Qvoxels ≤ 0.5

0 < Qvoxels < 0.1

Table S6.2.2: Peak *t* statistic value, peak Hedge's *g* value, and the fraction of ROI voxels significantly different between HVs and SCA7 patients (Qvoxels) for the pVF group comparison within each ROI of the FreeSurfer GM atlas.

**S6.3:** pMD full tables

**Table S6.3.1:** pMD full ICBM-DTI-81 WM atlas table

| ROI | *t* peak | *g* peak | Qvoxels |
|---|---|---|---|
| Middle cerebellar peduncle | 10.373 | 4.162 | 0.608 |
| Pontine crossing tract (a part of MCP) | 5.913 | 2.404 | 0.812 |
| Genu of corpus callosum | 4.262 | 1.695 | 0.123 |
| Body of corpus callosum | 5.362 | 2.172 | 0.192 |
| Splenium of corpus callosum | 5.834 | 1.881 | 0.137 |
| Fornix (column and body of fornix) | 0.000 | 0.000 | 0.000 |
| Corticospinal tract R | 5.920 | 2.260 | 0.619 |
| Corticospinal tract L | 7.029 | 2.764 | 0.641 |
| Medial lemniscus R | 7.612 | 2.977 | 0.423 |
| Medial lemniscus L | 5.974 | 2.326 | 0.328 |
| Inferior cerebellar peduncle R | 10.308 | 4.033 | 0.297 |

| | | | |
|---|---|---|---|
| Inferior cerebellar peduncle L | 12.316 | 4.921 | 0.313 |
| Superior cerebellar peduncle R | 6.349 | 2.557 | 0.162 |
| Superior cerebellar peduncle L | 5.106 | 1.967 | 0.222 |
| Cerebral peduncle R | 5.351 | 2.161 | 0.321 |
| Cerebral peduncle L | 6.197 | 2.420 | 0.312 |
| Anterior limb of internal capsule R | 4.522 | 1.690 | 0.044 |
| Anterior limb of internal capsule L | 4.916 | 1.892 | 0.151 |
| Posterior limb of internal capsule R | 5.782 | 2.360 | 0.318 |
| Posterior limb of internal capsule L | 6.352 | 2.566 | 0.577 |
| Retrolenticular part of internal capsule R | 4.894 | 2.022 | 0.290 |
| Retrolenticular part of internal capsule L | 5.412 | 2.222 | 0.363 |
| Anterior corona radiata R | 4.970 | 1.377 | 0.135 |
| Anterior corona radiata L | 5.491 | 2.239 | 0.181 |
| Superior corona radiata R | 6.310 | 2.559 | 0.505 |
| Superior corona radiata L | 6.439 | 2.606 | 0.550 |
| Posterior corona radiata R | 4.903 | 2.014 | 0.338 |
| Posterior corona radiata L | 6.521 | 2.526 | 0.468 |
| Posterior thalamic radiation R | 6.861 | 2.763 | 0.658 |
| Posterior thalamic radiation L | 4.747 | 1.884 | 0.515 |
| Sagittal stratum R | 5.369 | 2.078 | 0.292 |
| Sagittal stratum L | 4.059 | 1.614 | 0.430 |
| External capsule R | 3.707 | 1.440 | 0.031 |
| External capsule L | 6.396 | 1.953 | 0.131 |
| Cingulum (cingulate gyrus) R | 3.164 | 1.264 | 0.048 |
| Cingulum (cingulate gyrus) L | 4.474 | 1.675 | 0.060 |
| Cingulum (hippocampus) R | 3.127 | 1.293 | 0.005 |
| Cingulum (hippocampus) L | 2.528 | 1.029 | 0.033 |
| Fornix (cres) / Stria terminalis R | 5.071 | 2.090 | 0.300 |
| Fornix (cres) / Stria terminalis L | 5.220 | 2.085 | 0.366 |
| Superior longitudinal fasciculus R | 4.969 | 1.934 | 0.306 |
| Superior longitudinal fasciculus L | 6.649 | 2.636 | 0.302 |
| Superior fronto-occipital fasciculus R | 3.682 | 1.225 | 0.163 |
| Superior fronto-occipital fasciculus L | 4.533 | 1.742 | 0.161 |
| Uncinate fasciculus R | 0.000 | 0.000 | 0.000 |
| Uncinate fasciculus L | 3.531 | 1.463 | 0.112 |
| Tapetum R | 4.788 | 1.898 | 0.236 |
| Tapetum L | 3.186 | 1.350 | 0.057 |

Legend:

0.5 ≤ Qvoxels

0.1 < Qvoxels ≤ 0.5

0 < Qvoxels < 0.1

Table S6.3.1: Peak *t* statistic value, peak Hedge's *g* value, and the fraction of ROI voxels significantly different between HVs and SCA7 patients (Qvoxels) for the pMD group comparison within each ROI of the ICBM-DTI-81 WM atlas.

**Table S6.3.2:** pMD FreeSurfer full GM atlas table

| ROI | t_peak | g_peak | Qvoxels |
|---|---|---|---|
| Left-Cerebellum-Cortex | 6.725 | 2.534 | 0.098 |
| Left-Thalamus-Proper | 7.045 | 2.828 | 0.256 |
| Left-Caudate | 3.484 | 1.355 | 0.026 |
| Left-Putamen | 4.080 | 1.561 | 0.075 |
| Left-Pallidum | 5.079 | 1.996 | 0.185 |
| Brain-Stem | 10.308 | 4.033 | 0.408 |
| Left-Hippocampus | 3.920 | 1.512 | 0.065 |
| Left-Amygdala | 3.786 | 1.580 | 0.112 |
| Left-Accumbens-area | 0.000 | 0.000 | 0.000 |
| Left-VentralDC | 6.815 | 2.420 | 0.192 |
| Right-Cerebellum-Cortex | 9.328 | 3.629 | 0.151 |
| Right-Thalamus-Proper | 6.758 | 2.686 | 0.215 |
| Right-Caudate | 2.977 | 1.277 | 0.007 |
| Right-Putamen | 3.731 | 1.395 | 0.016 |
| Right-Pallidum | 4.541 | 1.740 | 0.177 |
| Right-Hippocampus | 2.758 | 0.990 | 0.006 |
| Right-Amygdala | 2.222 | 0.838 | 0.002 |
| Right-Accumbens-area | 0.000 | 0.000 | 0.000 |
| Right-VentralDC | 5.351 | 2.164 | 0.162 |
| ctx-lh-bankssts | 0.000 | 0.000 | 0.000 |
| ctx-lh-caudalanteriorcingulate | 3.214 | 1.185 | 0.040 |
| ctx-lh-caudalmiddlefrontal | 4.351 | 1.730 | 0.113 |
| ctx-lh-cuneus | 4.344 | 1.716 | 0.102 |
| ctx-lh-entorhinal | 3.674 | 1.556 | 0.032 |
| ctx-lh-fusiform | 4.972 | 1.862 | 0.050 |
| ctx-lh-inferiorparietal | 5.441 | 2.127 | 0.089 |
| ctx-lh-inferiortemporal | 3.796 | 1.606 | 0.034 |
| ctx-lh-isthmuscingulate | 4.183 | 1.576 | 0.013 |
| ctx-lh-lateraloccipital | 5.182 | 2.133 | 0.094 |
| ctx-lh-lateralorbitofrontal | 5.058 | 1.772 | 0.062 |
| ctx-lh-lingual | 4.556 | 1.851 | 0.091 |
| ctx-lh-medialorbitofrontal | 4.230 | 1.603 | 0.017 |
| ctx-lh-middletemporal | 4.052 | 1.591 | 0.068 |
| ctx-lh-parahippocampal | 2.930 | 1.230 | 0.009 |
| ctx-lh-paracentral | 5.010 | 1.952 | 0.073 |
| ctx-lh-parsopercularis | 4.255 | 1.488 | 0.066 |
| ctx-lh-parsorbitalis | 2.906 | 1.110 | 0.032 |
| ctx-lh-parstriangularis | 2.280 | 1.012 | 0.003 |
| ctx-lh-pericalcarine | 3.327 | 1.249 | 0.029 |
| ctx-lh-postcentral | 6.315 | 2.033 | 0.136 |
| ctx-lh-posteriorcingulate | 3.772 | 1.534 | 0.028 |
| ctx-lh-precentral | 5.175 | 1.940 | 0.139 |
| ctx-lh-precuneus | 4.548 | 1.775 | 0.109 |
| ctx-lh-rostralanteriorcingulate | 3.158 | 1.105 | 0.031 |
| ctx-lh-rostralmiddlefrontal | 4.807 | 1.789 | 0.104 |
| ctx-lh-superiorfrontal | 5.469 | 2.236 | 0.160 |
| ctx-lh-superiorparietal | 4.730 | 1.912 | 0.131 |
| ctx-lh-superiortemporal | 3.890 | 1.413 | 0.035 |
| ctx-lh-supramarginal | 4.808 | 1.990 | 0.063 |
| ctx-lh-frontalpole | 3.000 | 1.163 | 0.074 |
| ctx-lh-temporalpole | 2.446 | 1.008 | 0.002 |
| ctx-lh-transversetemporal | 3.804 | 1.423 | 0.039 |
| ctx-lh-insula | 4.353 | 1.753 | 0.030 |
| ctx-rh-bankssts | 3.171 | 1.308 | 0.026 |
| ctx-rh-caudalanteriorcingulate | 3.372 | 1.297 | 0.035 |

| | | | |
|---|---|---|---|
| ctx-rh-caudalmiddlefrontal | 4.783 | 1.755 | 0.085 |
| ctx-rh-cuneus | 3.917 | 1.508 | 0.078 |
| ctx-rh-entorhinal | 0.000 | 0.000 | 0.000 |
| ctx-rh-fusiform | 4.638 | 1.897 | 0.086 |
| ctx-rh-inferiorparietal | 5.873 | 2.124 | 0.092 |
| ctx-rh-inferiortemporal | 5.056 | 2.068 | 0.017 |
| ctx-rh-isthmuscingulate | 4.370 | 1.820 | 0.018 |
| ctx-rh-lateraloccipital | 5.069 | 1.999 | 0.124 |
| ctx-rh-lateralorbitofrontal | 0.000 | 0.000 | 0.000 |
| ctx-rh-lingual | 5.316 | 1.733 | 0.103 |
| ctx-rh-medialorbitofrontal | 0.000 | 0.000 | 0.000 |
| ctx-rh-middletemporal | 4.424 | 1.842 | 0.004 |
| ctx-rh-parahippocampal | 2.344 | 0.982 | 0.001 |
| ctx-rh-paracentral | 5.025 | 2.031 | 0.158 |
| ctx-rh-parsopercularis | 3.554 | 1.327 | 0.029 |
| ctx-rh-parsorbitalis | 0.000 | 0.000 | 0.000 |
| ctx-rh-parstriangularis | 4.156 | 1.683 | 0.095 |
| ctx-rh-pericalcarine | 5.323 | 2.163 | 0.083 |
| ctx-rh-postcentral | 5.224 | 1.881 | 0.047 |
| ctx-rh-posteriorcingulate | 4.177 | 1.669 | 0.028 |
| ctx-rh-precentral | 4.871 | 1.995 | 0.068 |
| ctx-rh-precuneus | 5.246 | 1.903 | 0.097 |
| ctx-rh-rostralanteriorcingulate | 3.333 | 1.143 | 0.008 |
| ctx-rh-rostralmiddlefrontal | 4.870 | 1.710 | 0.047 |
| ctx-rh-superiorfrontal | 4.921 | 1.980 | 0.056 |
| ctx-rh-superiorparietal | 5.431 | 2.224 | 0.109 |
| ctx-rh-superiortemporal | 3.418 | 1.430 | 0.001 |
| ctx-rh-supramarginal | 4.293 | 1.585 | 0.075 |
| ctx-rh-frontalpole | 0.000 | 0.000 | 0.000 |
| ctx-rh-temporalpole | 0.000 | 0.000 | 0.000 |
| ctx-rh-transversetemporal | 3.344 | 1.416 | 0.013 |
| ctx-rh-insula | 2.685 | 1.153 | 0.001 |

Legend:

0.5 ≤ Qvoxels

0.1 < Qvoxels ≤ 0.5

0 < Qvoxels < 0.1

Table S6.3.2: Peak *t* statistic value, peak Hedge's *g* value, and the fraction of ROI voxels significantly different between HVs and SCA7 patients (Qvoxels) for the pMD group comparison within each ROI of the FreeSurfer GM atlas.

**S6.4:** pFA full tables

**Table S6.4.1:** pFA full ICBM-DTI-81 WM atlas table

| ROI | *t* peak | *g* peak | Qvoxels |
|---|---|---|---|
| Middle cerebellar peduncle | -11.768 | -4.686 | 0.550 |
| Pontine crossing tract (a part of MCP) | -7.721 | -3.116 | 0.806 |

| | | | |
|---|---|---|---|
| Genu of corpus callosum | -4.459 | -1.304 | 0.005 |
| Body of corpus callosum | -4.279 | -1.752 | 0.124 |
| Splenium of corpus callosum | -4.602 | -1.909 | 0.203 |
| Fornix (column and body of fornix) | 0.000 | 0.000 | 0.000 |
| Corticospinal tract R | -6.511 | -2.609 | 0.579 |
| Corticospinal tract L | -7.584 | -3.066 | 0.510 |
| Medial lemniscus R | -10.778 | -4.266 | 0.525 |
| Medial lemniscus L | -8.643 | -3.334 | 0.438 |
| Inferior cerebellar peduncle R | -10.071 | -3.917 | 0.311 |
| Inferior cerebellar peduncle L | -8.061 | -3.225 | 0.203 |
| Superior cerebellar peduncle R | -7.301 | -2.800 | 0.143 |
| Superior cerebellar peduncle L | -6.754 | -2.308 | 0.242 |
| Cerebral peduncle R | -6.664 | -2.698 | 0.349 |
| Cerebral peduncle L | -6.609 | -2.652 | 0.413 |
| Anterior limb of internal capsule R | -3.825 | -1.608 | 0.027 |
| Anterior limb of internal capsule L | -4.673 | -1.816 | 0.384 |
| Posterior limb of internal capsule R | -6.302 | -2.178 | 0.266 |
| Posterior limb of internal capsule L | -7.149 | -2.773 | 0.315 |
| Retrolenticular part of internal capsule R | -6.232 | -2.320 | 0.326 |
| Retrolenticular part of internal capsule L | -5.730 | -2.127 | 0.267 |
| Anterior corona radiata R | -4.064 | -1.294 | 0.021 |
| Anterior corona radiata L | -4.807 | -1.971 | 0.121 |
| Superior corona radiata R | -5.850 | -2.097 | 0.279 |
| Superior corona radiata L | -6.521 | -2.592 | 0.346 |
| Posterior corona radiata R | -5.081 | -1.926 | 0.064 |
| Posterior corona radiata L | -5.049 | -2.066 | 0.204 |
| Posterior thalamic radiation R | -7.608 | -3.032 | 0.537 |
| Posterior thalamic radiation L | -5.455 | -2.224 | 0.483 |
| Sagittal stratum R | -5.543 | -2.263 | 0.248 |
| Sagittal stratum L | -5.704 | -2.303 | 0.329 |
| External capsule R | -3.557 | -1.373 | 0.020 |
| External capsule L | -5.235 | -1.955 | 0.129 |
| Cingulum (cingulate gyrus) R | -3.634 | -1.379 | 0.023 |
| Cingulum (cingulate gyrus) L | -5.321 | -1.861 | 0.106 |
| Cingulum (hippocampus) R | -2.647 | -1.111 | 0.013 |
| Cingulum (hippocampus) L | 0.000 | 0.000 | 0.000 |
| Fornix (cres) / Stria terminalis R | -3.917 | -1.649 | 0.212 |
| Fornix (cres) / Stria terminalis L | -6.761 | -2.744 | 0.359 |
| Superior longitudinal fasciculus R | -6.822 | -1.682 | 0.161 |
| Superior longitudinal fasciculus L | -4.146 | -1.670 | 0.091 |
| Superior fronto-occipital fasciculus R | -3.102 | -0.991 | 0.107 |
| Superior fronto-occipital fasciculus L | -4.162 | -1.684 | 0.085 |
| Uncinate fasciculus R | 0.000 | 0.000 | 0.000 |
| Uncinate fasciculus L | 0.000 | 0.000 | 0.000 |
| Tapetum R | -3.938 | -1.589 | 0.249 |
| Tapetum L | -4.678 | -1.936 | 0.182 |

Legend:

0.5 ≤ Qvoxels

0.1 < Qvoxels ≤ 0.5

0 < Qvoxels < 0.1

Table S6.4.1: Peak *t* statistic value, peak Hedge's *g* value, and the fraction of ROI voxels significantly different between HVs and SCA7 patients (Qvoxels) for the pFA group comparison within each ROI of the ICBM-DTI-81 WM atlas.

**Table S6.4.2:** pFA full FreeSurfer GM atlas table

| ROI | t_peak | g_peak | Qvoxels |
|---|---|---|---|
| Left-Cerebellum-Cortex | -9.739 | -3.789 | 0.548 |
| Left-Thalamus-Proper | -8.241 | -3.322 | 0.168 |
| Left-Caudate | -3.410 | -1.418 | 0.064 |
| Left-Putamen | -3.937 | -1.536 | 0.163 |
| Left-Pallidum | -6.079 | -2.372 | 0.180 |
| Brain-Stem | -10.778 | -4.266 | 0.370 |
| Left-Hippocampus | -3.060 | -1.211 | 0.009 |
| Left-Amygdala | -2.543 | -1.130 | 0.026 |
| Left-Accumbens-area | 0.000 | 0.000 | 0.000 |
| Left-VentralDC | -6.330 | -2.553 | 0.302 |
| Right-Cerebellum-Cortex | -10.831 | -4.254 | 0.608 |
| Right-Thalamus-Proper | -5.730 | -2.320 | 0.176 |
| Right-Caudate | -2.474 | -0.929 | 0.002 |
| Right-Putamen | -4.428 | -1.701 | 0.030 |
| Right-Pallidum | -5.333 | -2.144 | 0.183 |
| Right-Hippocampus | 0.000 | 0.000 | 0.000 |
| Right-Amygdala | 0.000 | 0.000 | 0.000 |
| Right-Accumbens-area | 0.000 | 0.000 | 0.000 |
| Right-VentralDC | -5.189 | -2.136 | 0.186 |

Legend:

0.5 ≤ Qvoxels

0.1 < Qvoxels ≤ 0.5

0 < Qvoxels < 0.1

Table S6.4.2: Peak *t* statistic value, peak Hedge's *g* value, and the fraction of ROI voxels significantly different between HVs and SCA7 patients (Qvoxels) for the pFA group comparison within each ROI of the FreeSurfer GM atlas.

**S6.5:** VBM full table

**Table S6.5.1:** VBM full FreeSurfer GM atlas table

| ROI | t_peak | g_peak | Qvoxels |
|---|---|---|---|
| Left-Cerebellum-Cortex | -11.242 | -4.199 | 0.282 |
| Left-Thalamus-Proper | -8.238 | -2.122 | 0.199 |
| Left-Caudate | -4.067 | -1.721 | 0.018 |
| Left-Putamen | -4.310 | -1.790 | 0.015 |
| Left-Pallidum | -4.453 | -1.647 | 0.030 |

| | | | |
|---|---|---|---|
| Brain-Stem | -12.066 | -4.570 | 0.794 |
| Left-Hippocampus | -3.084 | -0.988 | 0.007 |
| Left-Amygdala | 0.000 | 0.000 | 0.000 |
| Left-Accumbens-area | 0.000 | 0.000 | 0.000 |
| Left-VentralDC | -6.790 | -2.338 | 0.177 |
| Right-Cerebellum-Cortex | -10.527 | -2.847 | 0.243 |
| Right-Thalamus-Proper | -7.765 | -3.067 | 0.259 |
| Right-Caudate | -5.035 | -2.323 | 0.079 |
| Right-Putamen | -5.790 | -2.092 | 0.078 |
| Right-Pallidum | -5.851 | -2.170 | 0.080 |
| Right-Hippocampus | -3.486 | -1.171 | 0.020 |
| Right-Amygdala | 0.000 | 0.000 | 0.000 |
| Right-Accumbens-area | 0.000 | 0.000 | 0.000 |
| Right-VentralDC | -6.782 | -2.270 | 0.234 |
| ctx-lh-bankssts | 0.000 | 0.000 | 0.000 |
| ctx-lh-caudalanteriorcingulate | 0.000 | 0.000 | 0.000 |
| ctx-lh-caudalmiddlefrontal | 0.000 | 0.000 | 0.000 |
| ctx-lh-cuneus | 0.000 | 0.000 | 0.000 |
| ctx-lh-entorhinal | 0.000 | 0.000 | 0.000 |
| ctx-lh-fusiform | -5.154 | -2.174 | 0.054 |
| ctx-lh-inferiorparietal | 0.000 | 0.000 | 0.000 |
| ctx-lh-inferiortemporal | 0.000 | 0.000 | 0.000 |
| ctx-lh-isthmuscingulate | -2.966 | -0.794 | 0.005 |
| ctx-lh-lateraloccipital | 0.000 | 0.000 | 0.000 |
| ctx-lh-lateralorbitofrontal | 0.000 | 0.000 | 0.000 |
| ctx-lh-lingual | -4.064 | -1.559 | 0.089 |
| ctx-lh-medialorbitofrontal | 0.000 | 0.000 | 0.000 |
| ctx-lh-middletemporal | 0.000 | 0.000 | 0.000 |
| ctx-lh-parahippocampal | 0.000 | 0.000 | 0.000 |
| ctx-lh-paracentral | 0.000 | 0.000 | 0.000 |
| ctx-lh-parsopercularis | 0.000 | 0.000 | 0.000 |
| ctx-lh-parsorbitalis | 0.000 | 0.000 | 0.000 |
| ctx-lh-parstriangularis | 0.000 | 0.000 | 0.000 |
| ctx-lh-pericalcarine | 0.000 | 0.000 | 0.000 |
| ctx-lh-postcentral | -3.670 | -0.961 | 0.001 |
| ctx-lh-posteriorcingulate | 0.000 | 0.000 | 0.000 |
| ctx-lh-precentral | -4.331 | -0.936 | 0.021 |
| ctx-lh-precuneus | 0.000 | 0.000 | 0.000 |
| ctx-lh-rostralanteriorcingulate | 0.000 | 0.000 | 0.000 |
| ctx-lh-rostralmiddlefrontal | 0.000 | 0.000 | 0.000 |
| ctx-lh-superiorfrontal | 0.000 | 0.000 | 0.000 |
| ctx-lh-superiorparietal | -3.756 | -1.009 | 0.001 |
| ctx-lh-superiortemporal | 0.000 | 0.000 | 0.000 |
| ctx-lh-supramarginal | 0.000 | 0.000 | 0.000 |
| ctx-lh-frontalpole | 0.000 | 0.000 | 0.000 |
| ctx-lh-temporalpole | 0.000 | 0.000 | 0.000 |
| ctx-lh-transversetemporal | 0.000 | 0.000 | 0.000 |
| ctx-lh-insula | 0.000 | 0.000 | 0.000 |
| ctx-rh-bankssts | 0.000 | 0.000 | 0.000 |
| ctx-rh-caudalanteriorcingulate | 0.000 | 0.000 | 0.000 |
| ctx-rh-caudalmiddlefrontal | 0.000 | 0.000 | 0.000 |
| ctx-rh-cuneus | 0.000 | 0.000 | 0.000 |
| ctx-rh-entorhinal | -2.821 | -1.026 | 0.003 |
| ctx-rh-fusiform | -6.412 | -2.493 | 0.069 |
| ctx-rh-inferiorparietal | 0.000 | 0.000 | 0.000 |
| ctx-rh-inferiortemporal | 0.000 | 0.000 | 0.000 |

| | | | |
|---|---|---|---|
| ctx-rh-isthmuscingulate | 0.000 | 0.000 | 0.000 |
| ctx-rh-lateraloccipital | 0.000 | 0.000 | 0.000 |
| ctx-rh-lateralorbitofrontal | 0.000 | 0.000 | 0.000 |
| ctx-rh-lingual | -4.574 | -1.618 | 0.036 |
| ctx-rh-medialorbitofrontal | 0.000 | 0.000 | 0.000 |
| ctx-rh-middletemporal | 0.000 | 0.000 | 0.000 |
| ctx-rh-parahippocampal | 0.000 | 0.000 | 0.000 |
| ctx-rh-paracentral | 0.000 | 0.000 | 0.000 |
| ctx-rh-parsopercularis | 0.000 | 0.000 | 0.000 |
| ctx-rh-parsorbitalis | 0.000 | 0.000 | 0.000 |
| ctx-rh-parstriangularis | 0.000 | 0.000 | 0.000 |
| ctx-rh-pericalcarine | 0.000 | 0.000 | 0.000 |
| ctx-rh-postcentral | -3.905 | -1.344 | 0.053 |
| ctx-rh-posteriorcingulate | -2.881 | -0.778 | 0.004 |
| ctx-rh-precentral | -6.153 | -1.760 | 0.107 |
| ctx-rh-precuneus | 0.000 | 0.000 | 0.000 |
| ctx-rh-rostralanteriorcingulate | 0.000 | 0.000 | 0.000 |
| ctx-rh-rostralmiddlefrontal | 0.000 | 0.000 | 0.000 |
| ctx-rh-superiorfrontal | 0.000 | 0.000 | 0.000 |
| ctx-rh-superiorparietal | -4.216 | -1.341 | 0.020 |
| ctx-rh-superiortemporal | 0.000 | 0.000 | 0.000 |
| ctx-rh-supramarginal | -3.550 | -1.208 | 0.011 |
| ctx-rh-frontalpole | 0.000 | 0.000 | 0.000 |
| ctx-rh-temporalpole | 0.000 | 0.000 | 0.000 |
| ctx-rh-transversetemporal | 0.000 | 0.000 | 0.000 |
| ctx-rh-insula | 0.000 | 0.000 | 0.000 |

Legend:

0.5 ≤ Qvoxels

0.1 < Qvoxels ≤ 0.5

0 < Qvoxels < 0.1

Table S6.5.1: Peak *t* statistic value, peak Hedge's *g* value, and the fraction of ROI voxels significantly different between HVs and SCA7 patients (Qvoxels) for the VBM group comparison within each ROI of the FreeSurfer GM atlas.

#### **S6.6:** MD full tables

##### **Table S6.6.1:** MD full ICBM-DTI-81 WM atlas table

| ROI | *t* peak | *g* peak | Qvoxels |
|---|---|---|---|
| Middle cerebellar peduncle | 11.879 | 4.706 | 0.922 |
| Pontine crossing tract (a part of MCP) | 11.588 | 4.443 | 0.960 |
| Genu of corpus callosum | 5.617 | 1.825 | 0.074 |
| Body of corpus callosum | 4.942 | 1.703 | 0.196 |
| Splenium of corpus callosum | 4.803 | 1.687 | 0.079 |
| Fornix (column and body of fornix) | 3.251 | 1.088 | 0.023 |
| Corticospinal tract R | 7.170 | 2.889 | 0.786 |
| Corticospinal tract L | 7.453 | 2.918 | 0.812 |

| | | | |
|---|---|---|---|
| Medial lemniscus R | 11.524 | 4.396 | 0.773 |
| Medial lemniscus L | 10.868 | 3.923 | 0.781 |
| Inferior cerebellar peduncle R | 11.249 | 4.508 | 0.903 |
| Inferior cerebellar peduncle L | 10.490 | 4.042 | 0.813 |
| Superior cerebellar peduncle R | 14.568 | 5.362 | 0.896 |
| Superior cerebellar peduncle L | 11.237 | 4.472 | 0.838 |
| Cerebral peduncle R | 7.082 | 2.862 | 0.538 |
| Cerebral peduncle L | 8.495 | 2.989 | 0.538 |
| Anterior limb of internal capsule R | 6.560 | 2.608 | 0.010 |
| Anterior limb of internal capsule L | 6.849 | 2.725 | 0.342 |
| Posterior limb of internal capsule R | 6.621 | 2.688 | 0.647 |
| Posterior limb of internal capsule L | 8.300 | 3.254 | 0.772 |
| Retrolenticular part of internal capsule R | 5.996 | 2.429 | 0.369 |
| Retrolenticular part of internal capsule L | 6.082 | 2.457 | 0.330 |
| Anterior corona radiata R | 3.724 | 1.319 | 0.025 |
| Anterior corona radiata L | 4.343 | 1.607 | 0.110 |
| Superior corona radiata R | 6.012 | 2.440 | 0.356 |
| Superior corona radiata L | 6.240 | 2.372 | 0.735 |
| Posterior corona radiata R | 4.441 | 1.833 | 0.280 |
| Posterior corona radiata L | 4.918 | 1.982 | 0.424 |
| Posterior thalamic radiation R | 4.221 | 1.759 | 0.211 |
| Posterior thalamic radiation L | 4.808 | 1.854 | 0.290 |
| Sagittal stratum R | 3.957 | 1.587 | 0.239 |
| Sagittal stratum L | 4.260 | 1.773 | 0.449 |
| External capsule R | 2.847 | 1.228 | 0.004 |
| External capsule L | 5.381 | 2.086 | 0.181 |
| Cingulum (cingulate gyrus) R | 4.087 | 1.627 | 0.148 |
| Cingulum (cingulate gyrus) L | 5.324 | 1.946 | 0.299 |
| Cingulum (hippocampus) R | 4.182 | 1.697 | 0.199 |
| Cingulum (hippocampus) L | 3.332 | 1.004 | 0.042 |
| Fornix (cres) / Stria terminalis R | 5.883 | 2.396 | 0.696 |
| Fornix (cres) / Stria terminalis L | 7.055 | 2.816 | 0.669 |
| Superior longitudinal fasciculus R | 3.995 | 1.620 | 0.115 |
| Superior longitudinal fasciculus L | 4.862 | 1.990 | 0.331 |
| Superior fronto-occipital fasciculus R | 3.528 | 1.497 | 0.110 |
| Superior fronto-occipital fasciculus L | 3.707 | 1.509 | 0.268 |
| Uncinate fasciculus R | 0.000 | 0.000 | 0.000 |
| Uncinate fasciculus L | 5.660 | 2.171 | 0.378 |
| Tapetum R | 4.221 | 1.605 | 0.090 |
| Tapetum L | 3.335 | 1.282 | 0.020 |

Legend:

0.5 ≤ Qvoxels

0.1 < Qvoxels ≤ 0.5

0 < Qvoxels < 0.1

Table S6.6.1: Peak *t* statistic value, peak Hedge's *g* value, and the fraction of ROI voxels significantly different between HVs and SCA7 patients (Qvoxels) for the MD group comparison within each ROI of the ICBM-DTI-81 WM atlas.

**Table S6.6.2:** MD full FreeSurfer GM atlas table

| ROI | t_peak | g_peak | Qvoxels |
|---|---|---|---|
| Left-Cerebellum-Cortex | 11.504 | 4.558 | 0.807 |
| Left-Thalamus-Proper | 8.300 | 3.254 | 0.454 |
| Left-Caudate | 5.007 | 1.820 | 0.066 |
| Left-Putamen | 5.534 | 2.100 | 0.127 |
| Left-Pallidum | 4.688 | 1.939 | 0.103 |
| Brain-Stem | 14.568 | 5.362 | 0.742 |
| Left-Hippocampus | 5.023 | 1.876 | 0.192 |
| Left-Amygdala | 4.554 | 1.641 | 0.115 |
| Left-Accumbens-area | 0.000 | 0.000 | 0.000 |
| Left-VentralDC | 7.482 | 2.965 | 0.442 |
| Right-Cerebellum-Cortex | 11.628 | 4.418 | 0.815 |
| Right-Thalamus-Proper | 9.102 | 3.659 | 0.463 |
| Right-Caudate | 4.366 | 1.414 | 0.027 |
| Right-Putamen | 4.538 | 1.879 | 0.014 |
| Right-Pallidum | 4.457 | 1.778 | 0.123 |
| Right-Hippocampus | 4.651 | 1.847 | 0.154 |
| Right-Amygdala | 2.422 | 1.014 | 0.004 |
| Right-Accumbens-area | 0.000 | 0.000 | 0.000 |
| Right-VentralDC | 7.944 | 3.033 | 0.417 |
| ctx-lh-bankssts | 3.092 | 1.285 | 0.002 |
| ctx-lh-caudalanteriorcingulate | 4.777 | 1.654 | 0.262 |
| ctx-lh-caudalmiddlefrontal | 4.258 | 1.762 | 0.065 |
| ctx-lh-cuneus | 5.405 | 2.072 | 0.161 |
| ctx-lh-entorhinal | 4.981 | 1.967 | 0.210 |
| ctx-lh-fusiform | 5.619 | 2.188 | 0.129 |
| ctx-lh-inferiorparietal | 4.305 | 1.351 | 0.026 |
| ctx-lh-inferiortemporal | 4.134 | 1.597 | 0.051 |
| ctx-lh-isthmuscingulate | 4.514 | 1.517 | 0.134 |
| ctx-lh-lateraloccipital | 5.871 | 2.003 | 0.139 |
| ctx-lh-lateralorbitofrontal | 5.454 | 1.898 | 0.004 |
| ctx-lh-lingual | 5.089 | 2.079 | 0.159 |
| ctx-lh-medialorbitofrontal | 2.272 | 0.903 | 0.001 |
| ctx-lh-middletemporal | 3.388 | 1.419 | 0.006 |
| ctx-lh-parahippocampal | 5.180 | 1.988 | 0.074 |
| ctx-lh-paracentral | 4.604 | 1.594 | 0.117 |
| ctx-lh-parsopercularis | 4.748 | 1.785 | 0.080 |
| ctx-lh-parsorbitalis | 0.000 | 0.000 | 0.000 |
| ctx-lh-parstriangularis | 3.133 | 1.236 | 0.003 |
| ctx-lh-pericalcarine | 4.846 | 1.801 | 0.358 |
| ctx-lh-postcentral | 5.091 | 1.735 | 0.119 |
| ctx-lh-posteriorcingulate | 4.942 | 1.611 | 0.194 |
| ctx-lh-precentral | 5.123 | 1.975 | 0.089 |
| ctx-lh-precuneus | 6.319 | 1.701 | 0.123 |
| ctx-lh-rostralanteriorcingulate | 4.319 | 1.595 | 0.051 |
| ctx-lh-rostralmiddlefrontal | 5.234 | 1.827 | 0.039 |
| ctx-lh-superiorfrontal | 4.986 | 1.805 | 0.063 |
| ctx-lh-superiorparietal | 6.344 | 2.553 | 0.075 |
| ctx-lh-superiortemporal | 4.285 | 1.410 | 0.014 |
| ctx-lh-supramarginal | 4.123 | 1.603 | 0.027 |
| ctx-lh-frontalpole | 0.000 | 0.000 | 0.000 |
| ctx-lh-temporalpole | 3.692 | 1.317 | 0.034 |
| ctx-lh-transversetemporal | 3.464 | 1.318 | 0.011 |

| | | | |
|---|---|---|---|
| ctx-lh-insula | 5.865 | 2.171 | 0.147 |
| ctx-rh-bankssts | 3.669 | 1.096 | 0.016 |
| ctx-rh-caudalanteriorcingulate | 4.283 | 1.631 | 0.182 |
| ctx-rh-caudalmiddlefrontal | 3.539 | 1.100 | 0.014 |
| ctx-rh-cuneus | 5.312 | 2.094 | 0.197 |
| ctx-rh-entorhinal | 5.138 | 2.027 | 0.065 |
| ctx-rh-fusiform | 7.262 | 2.575 | 0.163 |
| ctx-rh-inferiorparietal | 4.354 | 1.478 | 0.083 |
| ctx-rh-inferiortemporal | 3.859 | 1.614 | 0.024 |
| ctx-rh-isthmuscingulate | 4.400 | 1.595 | 0.226 |
| ctx-rh-lateraloccipital | 5.761 | 2.188 | 0.233 |
| ctx-rh-lateralorbitofrontal | 0.000 | 0.000 | 0.000 |
| ctx-rh-lingual | 6.828 | 1.905 | 0.137 |
| ctx-rh-medialorbitofrontal | 0.000 | 0.000 | 0.000 |
| ctx-rh-middletemporal | 4.813 | 1.992 | 0.037 |
| ctx-rh-parahippocampal | 5.229 | 2.113 | 0.185 |
| ctx-rh-paracentral | 7.162 | 2.837 | 0.217 |
| ctx-rh-parsopercularis | 4.350 | 1.579 | 0.002 |
| ctx-rh-parsorbitalis | 0.000 | 0.000 | 0.000 |
| ctx-rh-parstriangularis | 4.350 | 1.615 | 0.001 |
| ctx-rh-pericalcarine | 6.305 | 2.230 | 0.347 |
| ctx-rh-postcentral | 4.085 | 1.450 | 0.064 |
| ctx-rh-posteriorcingulate | 5.867 | 1.645 | 0.228 |
| ctx-rh-precentral | 4.619 | 1.631 | 0.062 |
| ctx-rh-precuneus | 5.443 | 1.898 | 0.087 |
| ctx-rh-rostralanteriorcingulate | 2.911 | 1.070 | 0.005 |
| ctx-rh-rostralmiddlefrontal | 2.215 | 0.902 | 0.000 |
| ctx-rh-superiorfrontal | 4.933 | 1.831 | 0.057 |
| ctx-rh-superiorparietal | 5.419 | 1.970 | 0.079 |
| ctx-rh-superiortemporal | 3.487 | 1.213 | 0.011 |
| ctx-rh-supramarginal | 5.342 | 1.890 | 0.033 |
| ctx-rh-frontalpole | 0.000 | 0.000 | 0.000 |
| ctx-rh-temporalpole | 0.000 | 0.000 | 0.000 |
| ctx-rh-transversetemporal | 4.025 | 1.551 | 0.064 |
| ctx-rh-insula | 5.401 | 1.783 | 0.021 |

Legend:

0.5 ≤ Qvoxels

0.1 < Qvoxels ≤ 0.5

0 < Qvoxels < 0.1

Table S6.6.2: Peak *t* statistic value, peak Hedge's *g* value, and the fraction of ROI voxels significantly different between HVs and SCA7 patients (Qvoxels) for the MD group comparison within each ROI of the FreeSurfer GM atlas.

**S6.7:** FA full tables

**Table S6.7.1:** FA full ICBM-DTI-81 WM atlas table

| ROI | *t* peak | *g* peak | Qvoxels |
|---|---|---|---|
| Middle cerebellar peduncle | -14.007 | -4.536 | 0.740 |
| Pontine crossing tract (a part of MCP) | -7.921 | -2.995 | 0.910 |
| Genu of corpus callosum | -4.197 | -1.570 | 0.058 |
| Body of corpus callosum | -4.918 | -1.702 | 0.145 |
| Splenium of corpus callosum | -4.652 | -1.872 | 0.163 |
| Fornix (column and body of fornix) | -2.682 | -0.900 | 0.010 |
| Corticospinal tract R | -6.023 | -2.445 | 0.675 |
| Corticospinal tract L | -7.951 | -2.969 | 0.626 |
| Medial lemniscus R | -10.257 | -3.596 | 0.884 |
| Medial lemniscus L | -10.361 | -3.718 | 0.819 |
| Inferior cerebellar peduncle R | -13.507 | -4.956 | 0.671 |
| Inferior cerebellar peduncle L | -12.553 | -5.002 | 0.515 |
| Superior cerebellar peduncle R | -16.047 | -5.455 | 0.929 |
| Superior cerebellar peduncle L | -16.335 | -5.591 | 0.892 |
| Cerebral peduncle R | -7.620 | -2.961 | 0.456 |
| Cerebral peduncle L | -8.831 | -3.276 | 0.603 |
| Anterior limb of internal capsule R | -3.985 | -1.652 | 0.004 |
| Anterior limb of internal capsule L | -6.877 | -2.755 | 0.443 |
| Posterior limb of internal capsule R | -6.265 | -2.436 | 0.268 |
| Posterior limb of internal capsule L | -6.649 | -2.342 | 0.297 |
| Retrolenticular part of internal capsule R | -5.508 | -1.831 | 0.411 |
| Retrolenticular part of internal capsule L | -5.227 | -1.914 | 0.196 |
| Anterior corona radiata R | -5.664 | -1.745 | 0.004 |
| Anterior corona radiata L | -5.115 | -1.628 | 0.134 |
| Superior corona radiata R | -4.913 | -1.915 | 0.103 |
| Superior corona radiata L | -5.299 | -2.115 | 0.282 |
| Posterior corona radiata R | -3.843 | -1.598 | 0.037 |
| Posterior corona radiata L | -3.834 | -1.617 | 0.089 |
| Posterior thalamic radiation R | -4.702 | -1.945 | 0.432 |
| Posterior thalamic radiation L | -5.519 | -2.250 | 0.378 |
| Sagittal stratum R | -4.356 | -1.790 | 0.193 |
| Sagittal stratum L | -4.704 | -1.945 | 0.276 |
| External capsule R | -3.326 | -1.270 | 0.014 |
| External capsule L | -5.727 | -2.021 | 0.087 |
| Cingulum (cingulate gyrus) R | -3.267 | -1.312 | 0.007 |
| Cingulum (cingulate gyrus) L | -4.081 | -1.652 | 0.097 |
| Cingulum (hippocampus) R | -3.700 | -1.470 | 0.030 |
| Cingulum (hippocampus) L | 0.000 | 0.000 | 0.000 |
| Fornix (cres) / Stria terminalis R | -7.278 | -2.946 | 0.553 |
| Fornix (cres) / Stria terminalis L | -7.380 | -2.972 | 0.574 |
| Superior longitudinal fasciculus R | -3.665 | -1.127 | 0.010 |
| Superior longitudinal fasciculus L | -4.187 | -1.755 | 0.038 |
| Superior fronto-occipital fasciculus R | 0.000 | 0.000 | 0.000 |
| Superior fronto-occipital fasciculus L | -2.846 | -1.053 | 0.044 |
| Uncinate fasciculus R | 0.000 | 0.000 | 0.000 |
| Uncinate fasciculus L | 0.000 | 0.000 | 0.000 |
| Tapetum R | -3.923 | -1.621 | 0.288 |
| Tapetum L | -4.273 | -1.713 | 0.282 |

Legend:

0.5 ≤ Qvoxels

0.1 < Qvoxels ≤ 0.5

0 < Qvoxels < 0.1

Table S6.7.1: Peak *t* statistic value, peak Hedge's *g* value, and the fraction of ROI voxels significantly different between HVs and SCA7 patients (Qvoxels) for the FA group comparison within each ROI of the ICBM-DTI-81 WM atlas.

**Table S6.7.2:** FA full FreeSurfer GM atlas table

| ROI | t_peak | g_peak | Qvoxels |
|---|---|---|---|
| Left-Cerebellum-Cortex | -15.155 | -5.471 | 0.829 |
| Left-Thalamus-Proper | -6.382 | -2.560 | 0.276 |
| Left-Caudate | -4.602 | -1.833 | 0.136 |
| Left-Putamen | -4.722 | -1.881 | 0.127 |
| Left-Pallidum | -5.069 | -1.991 | 0.184 |
| Brain-Stem | -16.335 | -5.591 | 0.639 |
| Left-Hippocampus | -7.866 | -3.114 | 0.350 |
| Left-Amygdala | -6.477 | -2.105 | 0.118 |
| Left-Accumbens-area | 0.000 | 0.000 | 0.000 |
| Left-VentralDC | -8.732 | -3.276 | 0.524 |
| Right-Cerebellum-Cortex | -15.866 | -5.266 | 0.824 |
| Right-Thalamus-Proper | -6.969 | -2.816 | 0.261 |
| Right-Caudate | -2.263 | -1.039 | 0.002 |
| Right-Putamen | -4.328 | -1.788 | 0.029 |
| Right-Pallidum | -6.381 | -2.464 | 0.180 |
| Right-Hippocampus | -5.964 | -2.356 | 0.287 |
| Right-Amygdala | -3.476 | -1.345 | 0.051 |
| Right-Accumbens-area | 0.000 | 0.000 | 0.000 |
| Right-VentralDC | -7.614 | -2.925 | 0.435 |

Legend:

0.5 ≤ Qvoxels

0.1 < Qvoxels ≤ 0.5

0 < Qvoxels < 0.1

Table S6.7.2: Peak *t* statistic value, peak Hedge's *g* value, and the fraction of ROI voxels significantly different between HVs and SCA7 patients (Qvoxels) for the FA group comparison within each ROI of the FreeSurfer GM atlas.
